# Rapid review of the characteristics and outcomes of children involved in private family law proceedings due to parental separation

**DOI:** 10.1101/2024.12.04.24318480

**Authors:** Meg Kiseleva, Juliet Hounsome, Judit Csontos, Deborah Edwards, Elizabeth Gillen, Mala Mann, Rhiannon Tudor Edwards, Jacob Davies, Adrian Edwards, Alison Cooper, Ruth Lewis

## Abstract

Private law childrens proceedings typically involve court disputes between parents who have separated and disagree about child arrangements, and are asking the court to make orders that determine where a child should live and with whom they should spend time. Children involved in private law, who potentially represent a vulnerable group, commonly receive less attention in policy than those in public law cases. The aim of this review was to shine a light on the wellbeing and other important characteristics or outcomes of children who are currently, or have been, involved in family law proceedings due to parental separation, to identify the support needs of these children who are often overlooked in policy. This rapid review is intended for policymakers who are responsible for policy concerning children and families as well as for family law professionals and families in private law childrens proceedings.

The literature searches were conducted between June and August 2024. The included literature was published between 2001 and 2022. 22 studies reported in 25 documents were identified (8 published in academic journals and 17 in reports produced by organisations). Originated in England and Wales (n=13), Australia (n=7), Canada (n=1), New Zealand (n=1). Most studies aimed to describe the characteristics of children who are or have been involved in private family law proceedings, whilst only one compared the outcomes of such children to those in the general population.

Almost all of the studies addressed mental health and emotional wellbeing. Written accounts of children, parents, and professionals described children as having anxiety, depression, anger, post-traumatic stress disorder symptoms, and eating disorders, and experiencing self-harm and suicide attempts. In Wales, children with a history of involvement in private law proceedings had higher incidence of depression and anxiety than children in the general population. From the evidence, it was unclear whether the poor mental health was associated with parental separation, the court proceedings, court orders, or some other factors, but some participants attributed difficulties to unwanted court orders. Other key areas of evidence included engagement with mental health services, behaviour, development, social relationships, learning and education, and physical health.

Cardiff Evidence Synthesis Collaborative were funded for this work by the Health and Care Research Wales Evidence Centre, itself funded by Health and Care Research Wales on behalf of Welsh Government.

## GLOSSARY

- **Private law** cases involve court disputes between two or more private individuals, typically parents who have separated and disagree about child arrangement orders that determine where a child should live, and who they should spend time with (Ministry of Justice 2015).
- **Public law** involves state interventions to protect children at risk of harm, which may lead to care or supervision orders (Ministry of Justice 2015).
- **Cafcass** is a non-departmental public body in England sponsored by the Ministry of Justice that independently advises family courts about the best interests of children and young people.
- **Cafcass Cymru** operates in the same capacity in Wales. It is part of the Welsh Government.
- **Academic literature** is literature published in peer-reviewed academic journals.
- **Grey literature** is literature published outside of academic journals, for example, reports by government organisations, charities, research institutes etc.
- **Publication bias** is the trend for studies that report positive/statistically significant findings or findings that are perceived to be important to be more likely to be published or published quickly. Can be minimised by searching grey literature.
- **Qualitative research** is research that uses non-numeric data such as people’s views, for example, findings from interviews or focus groups.
- **Quantitative research** is research that uses numbers or statistical data.

## 1. BACKGROUND

### 1.1 Who is this review for?

This rapid review was conducted as part of the Health and Care Research Wales Evidence Centre Work Programme. The question was suggested by stakeholders from Cafcass Cymru. This review is intended primarily for policymakers who are responsible for policy concerning children and families but also for family law professionals and families in private law children’s proceedings.

### 1.2 Background and purpose of this review

#### 1.2.1 Private law landscape in England and Wales

Private law children’s proceedings typically involve court disputes between parents who have separated and disagree about child arrangements, and apply for orders that determine where a child should live and who they should spend time with (Ministry of Justice 2015). These proceedings are often initiated by a separated parent, usually the father (Cusworth et al. 2020). In contrast, public law involves state interventions to protect children at risk of harm, which may lead to care or supervision orders (Ministry of Justice 2015).

The number of private law applications in England and Wales has risen over the past few years, indicating the growing needs in this area (Cusworth et al. 2020), and is twice that of public law cases annually (Nuffield Family Justice Observatory 2021). In 2019/20 England had approximately 46,500 private law applications, increasing from about 35,000 in 2007/08 (Cusworth et al. 2021). Similarly, Wales also showed an increase in the same timeframe, with 3,390 private law applications in 2018 and 2,440 in 2007 (Cusworth et al. 2020). The rate of private family law applications per 10,000 family households has been growing slightly in both England and Wales (Cusworth et al. 2021, Cusworth et al. 2020). However, only a minority of separated families resort to court: this figure is estimated to be 10% or less (Cusworth et al. 2020).

Parental separation can be highly stressful for children. Research shows that children from separated families are more likely to be negatively affected in terms of their social, emotional, and physical wellbeing as well as education, and that adults who experienced parental divorce as children are at higher risk of poor mental health than other people (Symonds et al. 2022). Court involvement may further exacerbate this stress (Jones 2023). While only around 10% of separating parents go to court to settle disputes over their children, these tend to be complex cases of parental separation, marked by the inability to agree on arrangements concerning children. This may be due to a high degree of parental conflict or, frequently, alleged or confirmed domestic abuse. Indeed, the prevalence of alleged or confirmed domestic abuse in private law cases is estimated to be 49% to 62% (Hunter et al. 2020). For children, growing up in a situation of domestic abuse can be highly traumatic and is linked to poorer outcomes in later life (Symonds et al. 2022). Even witnessing abuse happening to a parent, without being a direct subject to it, can be damaging to children (Hunter et al. 2020).

At the same time, children involved in private law commonly receive less attention in policy than those in public law cases. The evidence base to inform private law policymaking is less developed than for public law (Cusworth et al. 2021). There are also concerns that child protection workers are more familiar with public law procedures and do not always have the necessary training to assist children in private law (Hunter et al. 2020). All this points towards the need for more attention towards this vulnerable group of children.

The aim of this review is to shine a light on the wellbeing and other important characteristics or outcomes of children who are currently, or have been, involved in family law proceedings due to parental separation, to paint a realistic picture of the support needs of these children who are often overlooked in policy. This work focuses on literature from England and Wales, but also includes literature from countries with comparable legal systems, as identified by the stakeholders from Cafcass Cymru: Australia, Canada, and New Zealand.

#### 1.2.2 Review question

The question guiding the focus of this review, developed with the stakeholders from Cafcass Cymru and a Health and Care Research Wales (HCRW) Evidence Centre public involvement member, is as follows: *What are the characteristics and outcomes of children involved in private family law proceedings due to parental separation?* The review is intended to inform readers about the mental health and wellbeing, education, and physical health needs of these children, and therefore broadly focuses on these factors.

## 2. RESULTS

Extensive literature searches were conducted, which included searching bibliographic databases, identifying literature included in existing reviews on similar topics, finding studies cited by or citing those already identified, and searching a large number of websites of relevant government, third sector, and research organisations. Details of the criteria used for selecting studies for inclusion in the review and review methods are provided in Section 5 of this report. The review included both descriptive studies, which describe the characteristics of a specific population, and analytic studies, which assess the impact of a specific exposure or observe the outcomes in a specific population. It included studies reporting quantitative data (numbers or statistics) and qualitative data (words or meaning).

A total of 22 studies reported in 25 documents were identified, originating from England (n=9), Wales (n=2), England and Wales (n=2), Australia (n=7), Canada (n=1), and New Zealand (n=1). The identified literature is a mix of articles published in academic journals (n=8) and grey literature reports (n=17). Ten studies used quantitative methods, eight qualitative methods, and four mixed methods.

The following seven broad themes for the characteristics and outcomes of the children were identified: mental health and emotional wellbeing, engagement with mental health services, behaviour, development, social relationships, learning and education, and physical health. The findings below are summarised according to these seven themes, with the findings from studies conducted in England and Wales presented first. Table 1 provides a summary of which outcomes were described in each study. A more detailed summary of the included studies and the full record of the relevant data extracted from these studies can be found in Section 6.2.

**Table 1:** Study characteristics.

| Reference<br>(First author, year) | Population characteristics | Study design* | Children's characteristics or outcomes |  |  |  |  |  |  |
| --- | --- | --- | --- | --- | --- | --- | --- | --- | --- |
|  |  |  | Mental health and emotional wellbeing | Engagement with mental health services | Behavioural | Developmental | Social relationships | Learning and educational | Physical health |
| England and/or Wales |  |  |  |  |  |  |  |  |  |
| Bailey 2011†/Timms 2007‡<sup>1</sup> | Welfare report/party to proceedings | Qualitative (survey) | ✓ | × | × | × | × | × | × |
| Bream 2003† | Welfare report | Quantitative (interviews with survey) | ✓ | × | × | × | × | × | × |
| Cafcass 2015‡ | Cafcass report (unspecified) | Quantitative (case review) | ✓ | ✓ | ✓ | × | × | × | × |
| Douglas 2006‡ | Rule 9.5 (separate representation) | Qualitative (interviews) | ✓ | ✓ | × | × | × | × | × |
| Green 2014‡ | Subject to Serious Case Review | Quantitative (case review) | ✓ | × | × | × | × | × | ✓ |
| Green 2016‡ | Subject to Serious Case Review | Quantitative (case review) | ✓ | × | × | × | × | × | ✓ |
| Green 2017‡ | Subject to Serious Case Review | Quantitative (case review) | ✓ | × | × | × | × | × | ✓ |
| Griffiths 2022a‡/2022b†<sup>2</sup> | Any | Quantitative comparative (cohort) | ✓ | × | × | × | × | × | × |
| Harold 2013‡ | Rule 16.4 (party to proceedings) | Mixed (survey) | ✓ | × | ✓ | × | ✓ | × | × |
| Hunter 2020‡ | Any but mostly domestic abuse | Qualitative (various methods) | ✓ | ✓ | ✓ | ✓ | ✓ | ✓ | ✓ |
| Trinder 2006‡ | Any | Mixed (interviews with survey) | ✓ | × | ✓ | × | ✓ | × | × |
| Trinder 2007‡ | Any | Quantitative (interviews with survey) | ✓ | × | ✓ | × | ✓ | × | × |
| Women's Aid 2022‡ | Domestic abuse | Qualitative (call for evidence) | ✓ | ✓ | × | ✓ | ✓ | × | ✓ |
| Australia, Canada, or New Zealand |  |  |  |  |  |  |  |  |  |
| Black 2021† | Any | Quantitative (cross-sectional) | ✓ | × | × | × | × | × | ✓ |
| Brown 2001‡ / 2002† (1)<sup>^3</sup> | Child abuse allegations | Quantitative (cross-sectional) | ✓ | ✓ | × | × | × | × | × |
| Brown 2001‡ / 2002† (2)<sup>^^3</sup> | Serious allegations of abuse | Quantitative (cross-sectional) | ✓ | ✓ | × | × | × | ✓ | × |
| Carson 2018‡ | Any | Mixed (interviews) | ✓ | ✓ | ✓ | × | ✓ | × | ✓ |
| Carson 2022‡ | Any | Mixed (survey, case review) | ✓ | × | ✓ | × | ✓ | ✓ | ✓ |
| Darlington 2001† | Any | Qualitative (interviews) | × | × | × | × | ✓ | × | × |
| Gollop 2020‡ | Any | Qualitative (interviews) | ✓ | ✓ | ✓ | × | ✓ | ✓ | × |
| Nelson 2022‡ | Domestic abuse | Qualitative (interviews) | ✓ | × | ✓ | × | ✓ | × | × |
| Shea Hart 2010†/2011†<sup>4</sup> | Domestic abuse | Qualitative (case review) | ✓ | × | ✓ | ✓ | ✓ | ✓ | ✓ |

\*If only the qualitative or quantitative part of a mixed-methods study was extracted, it is reported here as qualitative or quantitative accordingly; more detail in Section 6.2.
†Academic publication, ‡grey literature publication.
<sup>1</sup> Referred to as "Bailey et al. 2011", <sup>2</sup> as "Griffiths et al. 2022b", <sup>3</sup> as "Brown 2002", <sup>4</sup> as "Shea Hart 2011" in this report.
<sup>^</sup>"The First Study", <sup>^^</sup>"The Magellan Study".

Most of the studies from England and Wales limited inclusion to complex private law cases, for example, involving allegations or confirmed instances of domestic abuse, children for whom a welfare report was prepared or who were made party to the proceedings, or cases that were subject to a Serious Case Review (SCR), which happens when either a child dies and abuse or neglect is suspected to contribute to the death, or when a child has been seriously harmed and there are concerns about how well multi-agency working was performed. Only three of the studies from England and Wales did not limit their samples to particularly complex private law cases, and two of these studies included the same sample of participants. Of the studies from the comparable countries, four were focused on cases that included domestic or child abuse and the other five had no such sample restrictions. More information is available in Table 1.

### 2.1 Mental health and emotional wellbeing

#### 2.1.1 Overview of the evidence base

Children’s mental health and wellbeing outcomes were described in the majority of the identified studies, including 13 from England and Wales (Bailey et al. 2011, Women’s Aid 2022, Bream & Buchanan 2003, Cafcass Policy Team and National Improvement Service 2015, Douglas et al. 2006, Green & Halliday 2017, Green et al. 2014, Green et al. 2016, Griffiths et al. 2022b, Harold 2013, Hunter et al. 2020, Trinder et al. 2006, Trinder & Kellett 2007) and eight from the comparable countries (Black et al. 2021, Brown 2002 (two studies), Carson et al. 2018, Carson et al. 2022, Gollop et al. 2020, Nelson 2022, Shea Hart 2011). In terms of the study designs, seven studies from England and Wales were quantitative, four qualitative, and two mixed methods. The designs of the studies from the comparable countries were as follows: three quantitative, three qualitative, and two mixed methods.

#### 2.1.2 Findings

##### Evidence from England and Wales

First, we describe the evidence from England and Wales. The thirteen studies from England and Wales reporting mental health and emotional wellbeing included reports of children who were or had been involved in court proceedings due to parental separation experiencing: anxiety (Bailey et al. 2011, Cafcass Policy Team and National Improvement Service 2015, Griffiths et al. 2022b, Hunter et al. 2020, Women’s Aid 2022), unhappiness, sadness, and depression (Bailey et al. 2011, Douglas et al. 2006, Harold 2013, Hunter et al. 2020, Griffiths et al. 2022b), anger (Bailey et al. 2011, Hunter et al. 2020), trauma and post-traumatic stress disorder (PTSD) symptoms, such as nightmares and panic attacks (Women’s Aid 2022, Harold 2013, Hunter et al. 2020), eating disorders (Hunter et al. 2020), or self-harm and suicide attempts (Hunter et al. 2020).

Of note is the report of the findings of the Ministry of Justice’s (MoJ) expert panel, known as the Harm Panel, assessing the risk of harm to children and parents involved in private law children cases (Hunter et al. 2020). The report includes findings from a call for evidence that attracted more than 1,200 responses from individuals and organisations as well as roundtables and focus groups, much of which focused on cases involving allegations or confirmed instances of domestic abuse.

The responses received by the panel spoke of children experiencing a wide range of mental health problems, such as anxiety, depression, PTSD, complex PTSD, eating disorders, self-harm, and suicide attempts. It also notes a few cases of children’s suicide. Many of these were attributed to the, often protracted, private law proceedings. One example includes a childhood victim of domestic abuse responding to the call for evidence recalled becoming “very distressed” when she had to attend the court-ordered meetings with her father and “standing in the doorway of the [contact] centre screaming and refusing to go in” (Hunter et al. 2020, p.142). Another respondent sharing their experiences as a child involved in private law proceedings talked about suffering from anxiety and having problems with eating, sleeping, and toileting as a child and having a “profound fear of authority figures” as a teenager and adult that stemmed from seeing such figures as someone who could force them to do things against their will and punish them (Hunter et al. 2020, p.162). Yet another childhood victim talked about developing PTSD due to the stress they went through.

Reports from parents and professionals published in Hunter et al. (2020) highlight the same issues. One mother talked about her daughter self-harming and her sons being “deeply upset” and having “severe reactions” after receiving letters from their father in jail, who was allowed to contact the children once a month as an interim measure before the final court hearing. Another mother spoke of her son becoming suicidal after the court ordered contact with the father despite Cafcass’s recommendation for a prolonged period of therapy before contact took place. Other parents described their children as suffering from PTSD, “crying and shaking” when forced to attend a contact session with the father, as well as having sleeping problems, feeling “scared, isolated, and depressed”, bedwetting, suffering from nightmares, and having chronic stomach aches and vomiting in the week before contact. A divorce and domestic abuse professional responding to the call for evidence recalled examples of children hiding under their beds, locking themselves in rooms, running into a road, and hurting themselves to avoid unwanted contact with a parent, as well as becoming mute and catatonic when asked about the parents that they were scared of. In response to the Harm Panel’s findings, Women’s Aid published a report in which survivors of domestic abuse who had been involved in private law proceedings since the Harm Report’s publication (n=21), as well as professionals from specialist support services (n=10), shared their experiences with the private law system (Women’s Aid 2022). In the report, both survivors’ and professionals’ accounts showed that “two years on, private law children proceedings continue to be a source of re-traumatisation” (Women’s Aid 2022, p.11). Parents talked about their children having anxiety, panic attacks, stress symptoms, and nightmares, waking up multiple times in the night due to trauma, bedwetting, and having stomach aches, mirroring the accounts published in Hunter et al. (2020).

These qualitative findings are supported by quantitative data from nine studies from England and Wales. Bream & Buchanan (2003) interviewed parents (n=100) involved in private law proceedings whose children were subject to a welfare report for the court shortly after the end of the proceedings and one year later. Parents were asked to provide an assessment of their children using two standardised questionnaires: the Strengths and Difficulties Questionnaire (SDQ), which measures conduct, hyperactivity, peer relations, emotional problems, and prosocial behaviour, and the General Health Questionnaire (GHQ), which measures short-term changes in mental health and functioning. In this study, at the first interview, according to parental reports (n=56), 52% of boys and 48% of girls had borderline or abnormal scores on the SDQ, and at the second interview (n=47) those figures were 62% and 32% respectively. Children over eight years old (n=28) were also asked to complete the SDQ at the second time point and the percentage of children with borderline or abnormal scores was 36%. Data related to the subscales of the SDQ were not provided in this study, so it is not possible to report them here, even though some of the subscales would better fit other sections of this report. On the GHQ, there was a strong relationship between distress in children and parents, who also provided an assessment of their own wellbeing. The children aged 7–9 years old had the highest scores, and among the children under 7 years of age around half were found to be distressed at the first interview and 80% at the second.

Trinder et al. (2006) and Trinder & Kellett (2007) also used the SDQ. They interviewed parents sampled from court cases that had children named on the application. In the Trinder et al. (2006) study, the interviews happened within a few days of the conciliation appointment (n=250) and then 6–9 months later (n=175). The authors reported that the number of children with borderline or abnormal scores at baseline was double the national average at 42.9%, according to 156 parent-reports. When looking at the numbers reported by resident vs non-resident parent, 51% of the children had borderline or abnormal scores according to the resident parent (n=100) and 28.6% to the non-resident parent (n=56). This difference was statistically significant (*p=*0.011), suggesting a discrepancy in how resident and non-resident parents understood their children’s experiences. The authors suggested that resident parents were more likely to provide a more accurate assessment of their children’s wellbeing due to having more contact. The difference in the SDQ scores in resident vs non-resident parent-reports may reflect differences in how the children’s wellbeing and behaviour are experienced by each parent: for example, a child may be more likely to show distress at home.

At the follow-up in the Trinder et al. (2006) study, when only data from parents that were available at both time points were compared (n=106), the average SDQ scores decreased statistically significantly from 13.14 (SD=7.02) at baseline to 11.48 (SD=6.94) at follow-up (*p*=0.002). However, but there was not a statistically significant difference in the number of children with normal SDQ scores, with 57.5% being in the normal band at baseline and 66.0% at follow-up (*p*=0.078). The same parents were invited to participate in another follow-up on average 27 months after the first baseline interview and 117 parents took part (Trinder & Kellett 2007). In this follow-up study, 43% of the children had borderline or abnormal scores according to the resident parent and 35% according to all parent reports, again exceeding the UK average reported by the authors to be at 20%. Trinder & Kellett (2007) also ran a logistic regression analysis the likelihood of children scoring within the normal range on the SDQ (n=73), which showed that the best predictor of normal SDQ scores at the follow-up was a normal score at baseline (*p*=0.000) and that having contact, further litigation, and current adult wellbeing were not statistically significant predictors.

A more recent study examined the incidence of depression and anxiety in children involved in private law proceedings in Wales (Griffiths et al. 2022a, Griffiths et al. 2022b). It used linked population-level data across Wales from Cafcass Cymru and the Welsh Longitudinal General Practice (WLGP) data, resulting 17,041 records related to children in private law. The incidence of depression per 1,000 person-years at risk was 3.5 (95% CI 3.4–3.7), which was 60% higher in the private law group (IRR=1.6, 95% CI 1.4–1.7, adjusted for calendar year, gender, age, and deprivation) than in the general population comparison group.

Children in private law proceedings were also more likely to develop depression than those in the comparison group (HR=1.9, 95% CI 1.7–2.1, adjusted for previous history of depression and deprivation). Similar results were observed in the data on anxiety: the incidence per 1,000 person-years at risk was 4.3 (95% CI 4.2–4.5), which was 30% higher in the private law group (IRR=1.3, 95% CI 1.2–1.1.4, adjusted for calendar year, gender, age and deprivation) than in the comparison group. Children in the private law group were also significantly more likely to have anxiety than the control group (HR=1.4, 95% CI 1.2–1.6, adjusted for previous history of anxiety and deprivation).

Finally, we identified five studies from Cafcass or Cafcass Cymru that reported quantitative data on children involved in private law proceedings. A case review conducted by the Cafcass Policy Team and National Improvement Service (2015) examined the prevalence and nature of mental health concerns raised in cases in England in which Cafcass was involved. They selected a random sample of reports (n=20) filed to a family court from across all Cafcass local service areas, of which half (n=10) featured mental health concerns. Of these, five were in private law and reported anxiety (n=3) and low resilience and high vulnerability (n=1).

Three Cafcass reports reviewing submissions to SCRs were identified (Green & Halliday 2017, Green et al. 2014, Green et al. 2016). SCRs are conducted in cases when a child dies and abuse or neglect is suspected or confirmed to be a factor, or in cases of serious harm to a child where there are concerns about multi-agency working. The Cafcass reports assigned risk ratings to reviewed cases against 13 risk factors, such as emotional abuse. The ratings were based on what had been known to Cafcass about the recency, frequency, and severity of each factor at the time of their involvement in the case. In the Green et al. (2014) report, emotional abuse was found to be a risk in 53.8% of the 13 cases concerning private law, of which 12 were private law-only and one case involved both private and public law. Of these 13 cases, 23.1% were high risk, 15.4% medium risk, and 15.4% low risk. Similarly, of the seven private law cases in Green et al. (2016), emotional abuse was a risk in 57.1% (14.3% high risk, 28.6% medium risk, 14.3% low risk). The same document also reported an instance of a child in a both private and public case having many suicide attempts. Green & Halliday (2017) reported aggregated risk for domestic and emotional abuse and of the three private law cases in that report, 66.7% were at high risk.

An internal Cafcass Cymru study provided for the purposes of this review featuring 112 children from 81 families involved in private law proceedings found that 32% at risk were at risk of emotional problems, as per the SDQ (Harold 2013). Regarding the overall SDQ score measuring multiple problems (emotional, conduct, and hyperactivity), a total of 48% of the children were considered at risk. The qualitative component of this study reported some of these children to “feel so hurt” and have “many bad dreams” as well as “feel sick” or be “really really unhappy” when they had to be with one of the parents. One of the children described their life as “a living hell”. These were complex private law cases where Rule 16.4 of the Family Procedure Rules 2010 was applied to make the children party to the proceedings.

##### Evidence from Australia, Canada, and New Zealand

Findings from the comparable countries show more variability. In an Australian study of cases of a residence or contact dispute involving child abuse allegations (n=200), 28% of children showed high levels of distress (Brown 2002, “The First Study”). However, in another study by the same authors, of cases with serious allegations of abuse (n=175), it was 4% (Brown 2002 “The Magellan study”). A study of cases involving child custody or access disputes (estimated n=15,582) from Canada found that emotional harm was evident in 22.8% of the cases (Black et al. 2021).

Some court file data from Australia is also available. In a study that reported a review of 300 court file samples and 147 published and unpublished judgements (Carson et al. 2022), court data showed that 41.8% of the children had psychological needs, 36.1% mental health special needs, and 15.5% trauma relating to the requirement for having to spend time with their non-primary parent or carer. Another Australian study of judgements made in family courts, which concerned 33 children aged 2–16 represented in 20 judgements, reported that the judgements described the children as anxious, depressed, frightened, stressed, upset, unsettled, traumatised and having suicidal ideation, problems with sleep, difficulties concentrating, and “deep psychological scars” (Shea Hart 2011).

Nelson (2022) spoke with seven adult survivors of domestic abuse from Australia whose parents were involved in court proceedings when they were children. One of the participants, a woman in her 30s, described the emotional harm caused by the court as “more traumatic” than the family violence she had experienced. Another participant, a man in his 20s, said that the court-ordered telephone calls to his father were traumatising: “even to this day, it’s still…it still lingers. I would almost classify the whole scenario as like a type of PTSD, and looking back now, is…constant anger, sadness, a lot of frustration” (Nelson 2022, p.7).

Similarly, in a study reporting a survey parents and carers, 65% of those who identified issues relating to children (n=111) thought that the family law system harmed their children or failed to protect them from harm (Carson et al. 2022). Some parents and carers from New Zealand also spoke of the protracted proceedings and obstructive actions of the other parent causing stress to their children and making them go through “years of trauma” (Gollop et al. 2020). The parents and carers talked about children having anxiety attacks and self-harming. Some positive outcomes were also shared: some of the parents said that the arrangements provided their children with a routine, which reduced uncertainty and made them happier and less anxious.

Australian children (n=61) aged 10–17 years old interviewed by Carson et al. (2018) spoke about the court proceedings “detrimentally” impacting their sense of wellbeing and them having anxiety, trouble sleeping, and traumatic recurring dreams about the court. However, looking at the quantitative data from the same study, 16.4% reported being happy with life “all of the time”, 67.2% “most of the time”, 14.8% “sometimes”, and 1.6% “rarely”. Similarly, when asked about their overall mental and physical health, 26.7% of the participants said it was “excellent”, 28.3% “very good”, 31.7% “good”, 10.0% “fair”, and 3.3% “poor”.

#### 2.1.3 Bottom line results for mental health and emotional wellbeing

Together, these findings show that children involved in private law proceedings suffer from a wide range of mental health and emotional wellbeing problems, with some evidence suggesting that it is more so than children in the general population. Accounts from both England and Wales and from Australia, Canada, and New Zealand showed many children subject to family law proceedings due to parental separation to have high levels of anxiety and depression and suffer from trauma and emotional distress. There is a mix of qualitative and quantitative evidence in the identified literature, with the former containing rich first-hand accounts by children, parents, and professionals and the latter providing numeric data regarding children’s mental health and emotional wellbeing characteristics and outcomes.

The qualitative accounts are broadly similar both in the literature from England and Wales and from the comparable countries. They describe a wide range of mental health difficulties experienced by the children, such as anxiety, depression, anger, trauma and PTSD symptoms, eating disorders, self-harm, and suicide attempts. A few accounts by parents and carers from New Zealand described the positive effects of agreeing on child arrangements but such examples were not numerous. Due to the nature of the included studies, some of which included self-selecting samples and many focused on complex and particularly stressful cases, the available qualitative evidence is likely to be skewed towards accounts of negative experiences of contact with the family law system. This is not to dismiss such evidence: even without information on the prevalence of negative experiences, it illustrates the lived reality of children going through family court systems due to parental separation.

With regard to the quantitative evidence, most of the data from England and Wales was based on the SDQ and the majority of the included studies that used it did not report subscale-level data. However, across all the studies that used it and reported the overall SDQ score (Bream & Buchanan 2003, Trinder et al. 2006, Trinder & Kellett 2007, Harold 2013), around half the children had borderline or abnormal scores, so the evidence across the UK studies appears consistent. There is more variability in the results of the quantitative studies from the comparable countries, with some evidence showing much more positive outcomes. Fewer of these studies than those from England and Wales were limited to particularly complex cases and a wider range of instruments to measure children’s outcomes was used.

It was not possible to ascertain if the poor mental health outcomes were attributable to parental separation, the court proceedings, court orders, or other factors (for example, domestic abuse prevalent in private law cases) – or a combination of these. Due to the designs of the included studies, making causal inferences from the data also was not possible. However, particularly in the cases of domestic violence, many participants spoke of the stress and anxiety caused or exacerbated by family law proceedings and by unwanted court orders. Such orders were often described as putting children in highly stressful and dangerous situations, such as mandating that they spend time with an abusive parent. Many mental health difficulties were attributed to that in the participants’ accounts. This was the case in the qualitative evidence both from England and Wales and from the comparable countries. It appears that it is not the court proceedings per se but rather unwanted court orders that may underlie children’s mental health and emotional wellbeing struggles. The stress that parents’ experience from court proceedings may also be a factor in children’s mental health and emotional wellbeing.

Within the limitations of this rapid review, only academic studies were formally critically appraised in terms of their quality. More information about how the quality of the studies was assessed is provided in Section 5.6. The Welsh cohort study by Griffiths et al. (2022b) had some methodological limitations in terms of how the loss to follow-up was handled and the lack of clarity regarding the timelines for the development of outcomes (see Section 6.3 for detail), but overall it was performed well. The quality of most of the quantitative cross-sectional studies (Black et al. 2021, Bream & Buchanan 2003, Brown 2002 “The First Study”, “The Magellan Study”) was considered quite low. Participant inclusion criteria were not always clear, none of the studies provided detailed enough descriptions of the subjects and setting, and most did not identify or address confounding factors—that is, factors that may affect the relationship between the variables of interest. However, in all of these studies it was judged that children’s involvement in family law proceedings was measured in a valid and reliable way, and so were their outcomes. The Canadian study by Black et al. (2021) was considered to be the most reliably performed. With regard to the qualitative academic studies that informed this section of the report, one (Shea Hart 2011) was considered to be very well performed, with no serious methodological issues detected, and the other (Bailey et al. 2011) had some serious limitations, particularly with regard to how thoroughly it reported the participants’ views. It should be noted that the outcomes of interest in this review were not the main focus of the Bailey et al. (2011) study and our critical appraisal focused on what was relevant to the question of the review.

Regarding the certainty of the overall body of evidence, no formal assessment was performed within this rapid review, however, here we summarise some of the main factors. Section 5.8 provides more information on how these were identified. The studies included in this synthesis, both quantitative and qualitative, provided evidence that directly addressed the review question as they all include children involved in family law proceedings due to parental separation. The only exception is the Gollop et al. (2020) report from New Zealand which, in addition to parents and carers who had used family justice services, included those who had had limited or no service use.

The confidence in the findings from the overall body of quantitative evidence was assessed, where possible, in terms of methodological quality, consistency across study findings, precision, directness of the evidence, and the possibility of publication bias. As described above, the included academic studies had a number of methodological limitations, introducing a risk of bias; the methodological quality of the grey literature was not formally assessed. The findings of the studies from England and Wales related to mental health and emotional wellbeing were consistent with each other, but there was more variability in the evidence from the comparable countries, broadly showing less negative trends. In some but not all cases this may be because the samples were not limited to particularly complex and stressful family law cases. Most of the quantitative studies were descriptive and only provided the number or percentage of children in the sample experiencing an outcome. Only in the Griffiths et al. (2022b) and Trinder et al. (2006) studies were statistical analyses performed. In this type of review, a formal assessment of publication bias was not possible, but we included extensive grey literature searches to maximise the retrieval of evidence.

The confidence in the findings from the overall body of qualitative evidence was assessed in terms of methodological limitations, coherence, adequacy of data, and relevance to the research question. No formal quality assessment of the grey literature was conducted and the quality of the academic studies varied, as described earlier in this section. However, the included reports, together with the two academic studies that contributed to this section, were numerous and provided rich qualitative accounts by a large number of children, parents, and professionals. The evidence consistently showed the struggles that children face, with the same themes appearing across different studies.

### 2.2 Engagement with mental health services

#### 2.2.1 Overview of the evidence base

Eight studies, four from England and Wales (Cafcass Policy Team and National Improvement Service 2015, Douglas et al. 2006, Hunter et al. 2020, Women’s Aid 2022) and four from the comparable countries (Brown 2002 (two studies), Carson et al. 2018, Gollop et al. 2020) covered children’s engagement with mental health services.

#### 2.2.2 Findings

##### Evidence from England and Wales

Many of the submissions to the Harm Panel spoke of children being either referred to or in need of a referral to CAMHS, including a childhood victim of domestic abuse who described themselves as “lucky” for coming under CAMHS for their PTSD because it “saved [them] from further unwanted contact [with a parent]” (Hunter et al. 2020). A participant in the Douglas et al. (2006) study, 14 years old at the time, talked about having to see a “mind doctor” because of his father’s actions. He recalled: “I think mum maybe knew that I was getting to the point where I would do something silly with the stress because I just couldn’t cope and I think the court saw that as well” (Douglas et al. 2006, p.51).

All of the children in private law cases with identified mental health concerns (n=5) in the Cafcass Policy Team and National Improvement Service (2015) report were receiving some level of support, such as school-based support (n=2), professional services (n=1), or specialist multi-disciplinary team support (n=2). The specifics of the support were as follows: school “team around the child” (n=1), unspecified school support (n=1), school providing an external counsellor (n=1), CAMHS referral (n=1), receiving CAMHS support (n=1).

The Women’s Aid (2022) report warned of children in private law proceedings being prevented from accessing mental health support. A representative of a support service for survivors of sexual violence and child sexual abuse responding to the call for evidence spoke of the lack of clear guidance on whether children in private law cases could access therapy for sexual abuse and that mothers bringing children to play therapy might be viewed as “corroborating a false narrative”. Two children described by the respondent had been denied therapy which they would have had a right to had they been involved in a criminal investigation instead.

##### Evidence from Australia, Canada, and New Zealand

Australian children in at least 68% of the cases in the “First Study” (n=200) and in 63% of cases in the “Magellan Study” (n=175) were reported to use counselling services (Brown 2002). Similarly, another, much later, Australian study (n=61) reported that 62.3% of the children had engagement with mental health services (Carson et al. 2018). Some of the parents and carers from New Zealand interviewed by Gollop et al. (2020) raised the issue of the lack of support for post-separation issues from mental health or counselling services available to children. One mother recalled being turned away by a counselling agency and psychiatric services because her son’s issues were caused by the parents’ actions. She said: “Psych services assessed him and basically said, ‘He’s fine, there’s nothing mentally wrong with him. It’s you guys. You need to sort yourselves out.’ Which, obviously I knew that, that’s really helpful!” (Gollop et al. 2020, p.159). Other parents in the study spoke of their children attending counselling, including school counselling services.

#### 2.2.3 Bottom line results for engagement with mental health services

Little information on the use of mental health services by children in private law proceedings in England and Wales has been identified. The available evidence shows variable levels of support accessed by the children, from school counselling to engagement with CAMHS, however, the prevalence of service use is not known, and neither is how many children in private law proceedings are in need of mental health services and how many of those are able to access them. More research on this subject would be helpful. Looking at the evidence from Australia, studies conducted almost two decades apart showed that over half of the children involved in them were accessing mental health services. Only descriptive statistics were provided. No comparison with the general population was available.

### 2.3 Behaviour

#### 2.3.1 Overview of the evidence base

Behavioural outcomes were described in five studies from England and Wales (Cafcass Policy Team and National Improvement Service 2015, Harold 2013, Hunter et al. 2020, Trinder et al. 2006, Trinder & Kellett 2007) and five studies from the comparable countries (Carson et al. 2018, Carson et al. 2022, Gollop et al. 2020, Nelson 2022, Shea Hart 2011).

#### 2.3.2 Findings

##### Evidence from England and Wales

In a Cafcass Cymru study of 112 children from 81 families subject to Rule 16.4 of the Family Procedure Rules 2010, 33% were at risk of conduct problems according to the SDQ (Harold 2013). Behaviour problems were reported in one of the five private law cases in the Cafcass Policy Team and National Improvement Service (2015) study. Trinder et al. (2006) and Trinder & Kellett (2007) reported children’s SDQ scores. The SDQ includes a conduct problems subscale. However, no subscale-level data was provided in either report, so it is not possible to untangle conduct problems from the overall SDQ score. The overall SDQ data from these two studies is provided in Section 2.1.2 of this report. Some qualitative data pertaining to children’s behaviour was reported in Hunter et al. (2020). Respondents spoke of children becoming “frustrated” and “lashing out” at the mother, turning “violent” after an order for unsupervised contact with a parent, and copying the father’s behaviour and becoming physically and verbally abusive to the mother and younger siblings.

##### Evidence from Australia, Canada, and New Zealand

The Australian children interviewed by Carson et al. (2018) were asked about losing their temper. None of them reported that it happened “all of the time”, 4.9% said they did “most of the time”, 39.3% “sometimes”, 49.2% “rarely”, and 6.6% never. A review of Australian court files in the Carson et al. (2022) showed that 11.9% of the children had behavioural problems. One of the Australian participants in the Nelson (2022) study whose parents went to family court when he was younger recalled “acting out, and throwing tantrums, as a child” because he did not know how to verbalise his struggles due to his age at the time. A study that reviewed Australian court judgements (n=20) found that many judgements recorded children’s behavioural problems (Shea Hart 2011). The children were described as having “bad behaviour at school and home”, having regular detentions at school, being “boisterous and disruptive” towards other children, showing “uncontrollable”, “aggressive”, and “antisocial” behaviour, having been “violent” towards a sister and other girls at school, showing “anger and hostility”, acting out and “kicking, hitting, teasing and showing no remorse”, “defecating in the house and smearing faeces over herself, the walls and the furniture in the house”, and talking about the desire to “shoot” the father. In a study from New Zealand, a mother spoke of her son hiding under the bed, lashing out, beating an older sibling, and letting himself out at night (Gollop et al. 2020).

#### 2.3.3 Bottom line results for behaviour

The evidence that addresses this question was scarce. Some reports suggest that a substantial number of children involved in family law proceedings experience conduct problems, but it is unclear whether there is causal relationship between them and parental separation, court proceedings, or court orders. Some accounts attributed behavioural problems to frustration with and hurt from the situation the children find themselves in and difficulty in verbalising their feelings.

### 2.4 Development

#### 2.4.1 Overview of the evidence base

Developmental characteristics were only described in two studies from the UK (Hunter et al. 2020, Women’s Aid 2022) and one from Australia (Shea Hart 2011).

#### 2.4.2 Findings

##### Evidence from England and Wales

A divorce and domestic abuse professional responding to a call for evidence spoke of children in private law proceedings subjected to unwanted contact with a parent regressing “to a much younger age in behaviour” (Hunter et al. 2020). Similarly, a survivor of domestic abuse that responded to the call for evidence by Women’s Aid (2022) recalled that her daughter that was forced to go to contact sessions had “gone back years” in her development and regressed in toileting and wanted to wear nappies and use a pushchair.

##### Evidence from Australia, Canada, and New Zealand

One of the Australian court judgements (n=20) reviewed by Shea Hart (2011) described children experiencing speech and developmental delay problems and another reported that a child needed ongoing therapy for speech and verbal reasoning.

#### 2.4.3 Bottom line results for development

Little evidence is available on children’s developmental characteristics, but the few accounts that included it spoke of children experiencing developmental delays and regressing to an earlier developmental stage. However, it is not clear how widespread this issue is.

### 2.5 Social relationships

#### 2.5.1 Overview of the evidence base

Social relationships were covered in five studies from England and Wales (Harold 2013, Hunter et al. 2020, Trinder et al. 2006, Trinder & Kellett 2007, Women’s Aid 2022) and six studies from the comparable countries (Carson et al. 2018, Carson et al. 2022, Darlington 2001, Gollop et al. 2020, Nelson 2022, Shea Hart 2011).

#### 2.5.2 Findings

##### Evidence from England and Wales

Submissions to the Harm Panel described children’s relationships being “distorted by the ongoing abuse they experienced through contact”, children experiencing problems with friends or stopping socialising with peers altogether, having a damaged relationship with a parent due to blaming them for the situation they were put in, and becoming “hysterical” when in the same room with adult men (Hunter et al. 2020). The report also spoke of children growing up without appropriate role models and not being able to learn the differences between healthy and unhealthy relationships, which for some resulted in forming relationships with abusive partners in adulthood. Women’s Aid (2022) also reported an account of parent whose daughter forced to have unwanted contact with the other parent started falling out with her friends.

The Cafcass Cymru study measured children’s perceptions of parenting experiences (Harold 2013). Of the 112 children subject to Rule 16.4 of the Family Procedure Rules 2010, 26% were at risk of low acceptance by the mother and 31% by the father, 23% at risk of rejection by the mother and 25% by the father, and 38% at risk of hostile detachment by the mother and 39% by the father, as perceived by the children themselves. The SDQ used in Trinder et al. (2006) Trinder & Kellett (2007) contains a subscale measuring peer problems but no subscale data was provided in these studies. The overall SDQ scores are provided in Section 2.1.2 of this report.

##### Evidence from Australia, Canada, and New Zealand

Of the Australian children (n=61) interviewed by Carson et al. (2018), 26.7% said they got along with peers “all of the time”, 60.0% “most of the time”, 10.0% “sometimes”, 3.3% “rarely”, and 0% “never”. In terms of closeness to their parents, 79.7% said they were “very close”, 15.3% “quite close”, 3.4% “not very close”, and 1.7% “not close at all” to their mother and 22.4% “very close”, 34.5% “quite close”, 19.0% “not very close”, and 24.1% “not close at all” to their father. In the Carson et al. (2022) study, Australian parents and carers (n=470) reported how well their children got along with other peers compared with children of the same age: 4.8% said their children did “much better”, 7.4% “somewhat better”, 41.1% “about the same”, 21.4% “somewhat worse”, 12.9% “much worse”, 0.9% preferred not to say, and 7.4% did not know or could not say; 4.1% chose “other” as their response.

Similarly to the account in Hunter et al. (2020), one of the participants from Australia in the Nelson (2022) study, a woman in her 30s who had been a domestic abuse victim as a child, said she and her younger sibling were “frightened of men” when they were younger. A stepmother from New Zealand shared that the length of time the court proceedings took was damaging for the children’s relationship with their father and with herself (Gollop et al. 2020). Two of the Australian court judgements reviewed in the Shea Hart (2010, 2011) study described children having difficulties in relating to peers and experiencing social isolation.

Finally, the Darlington (2001) study from Australia focused on the social relationships of adults who had been subject to family court proceedings as children (n=18). Specifically, it described children’s relationships with parents and romantic partners. Seven reported that they’d always had a good relationship with both parents, five that they came to accept the non-preferred parent more than previously, three said that the court proceedings exacerbated the difficulties they had in the relationship with the non-preferred parent, and five had little or no contact with one of the parents. In terms of romantic relationships, all 18 said they did not want to repeat the pattern of divorce. Seven said they did not want to “rush into a relationship”, three highlighted the “need for emotional independence and a strong sense of self in a relationship”, three that they needed personal financial security, five spoke of the importance of communication with partners, and four talked about how if divorce was inevitable, it should be done “cleanly”.

#### 2.5.3 Bottom line results for social relationships

Some of the studies from England and Wales spoke of children’s difficulties in relationships with parents and peers. There were qualitative accounts of children who had problems with friends or stopped socialising with them altogether. What quantitative evidence was available came from a study with children involved in particularly complex private law cases and many of the children involved perceived their relationships with their parents as strained. The data from the comparable countries are more promising. In Australian studies, both the children themselves and their parents indicated that children had few problems with socialising with peers. Some qualitative evidence, however, also showed that there were children who struggled with social relationships. No data directly comparing such outcomes to those of children who did not go through family court proceedings was available.

### 2.6 Learning and education

#### 2.6.1 Overview of the evidence base

Learning and education were described in only one study from England and Wales (Hunter et al. 2020) and four studies from the comparable countries (Brown 2002 “The Magellan Study”, Carson et al. 2022, Gollop et al. 2020, Shea Hart 2011).

#### 2.6.2 Findings

##### Evidence from England and Wales

The Harm Panel received submissions describing children having their schooling affected by the proceedings, experiencing learning difficulties, and being excluded from school (Hunter et al. 2020). One mother responding to the call for evidence whose then-12–year-old son was forced to have contact with his father said: “His schooling went downhill, he was kicking off at school” (Hunter et al. 2020, p.154).

##### Evidence from Australia, Canada, and New Zealand

In an Australian study, 25% of the children (n=175) were reported to have learning problems (Brown 2002 “The Magellan Study”). In another study, also from Australia, court files showed that 34.5% of the children had learning difficulties (Carson et al. 2022). In addition, parents and carers (n=470) interviewed in this study reported their children’s learning or schoolwork outcomes compared with children of the same age: 6.7% said it was “much better”, 12.9% “somewhat better”, 29.4% “about the same”, 22.5% “somewhat worse”, 15.6% “much worse”, 0.5% preferred not to say, 4.6% did not know or could not say; and 7.8% chose “other”. Two of the court judgements from Australia (n=20) reviewed by Shea Hart (2011) described children struggling in school academically and having learning difficulties. Finally, in a report from New Zealand, a father described how his formerly “A-plus, amazing, doing well in school” son “went to Ds, Es” and was “kicked out of school” (Gollop et al. 2020).

#### 2.6.3 Bottom line results for learning and education

Only a few qualitative accounts of children’s learning and education from England and Wales were available, talking about children’s schooling being negatively affected, supported by similar data from the comparable countries. In an Australian study, almost 40% of parents thought that their children’s learning and schoolwork were worse than their peers’ (Carson et al. 2022). No data directly comparing learning and educational outcomes of children involved and not involved in family law proceedings was available. Overall, the volume of evidence to address this question was low.

### 2.7 Physical health

#### 2.7.1 Overview of the evidence base

Five studies from England and Wales (Green & Halliday 2017, Green et al. 2014, Green et al. 2016, Hunter et al. 2020, Women’s Aid 2022) and four from the comparable countries (Black et al. 2021, Carson et al. 2018, Carson et al. 2022, Shea Hart 2011) described children’s physical health outcomes.

#### 2.7.2 Findings

##### Evidence from England and Wales

Data on fatal and non-fatal incidents involving abuse and neglect was provided in the Cafcass reports reviewing submissions to SCRs in England. Green et al. (2014) reported a total of 26 cases, of which 12 were private law cases and two more concerned both private and public law. Of the 11 cases of fatal physical abuse, seven were in private law. There were two cases of spite or revenge killing, both in private law. Additionally, the report described an instance of non-fatal physical abuse in a private law case and two cases of non-fatal neglect. Of the 12 private law cases and one case that included both private and public law, physical abuse was a risk in 38.5% (15.4% high risk, 15.4% medium risk, 7.7% low risk).

In the Green et al. (2016) report that included seven private law and three private and public law cases, there was one instance of fatal physical abuse, one instance of fatal neglect, two instances of spite or revenge killing, and one suicide in the private law cases. In addition, there was one instance of non-fatal neglect in a private law case as well an instance of intrafamilial sexual abuse in a both private and public law case. Of the seven private law cases, 42.8% were at risk of physical abuse (28.6% high risk, 14.3% medium risk). The Green & Halliday (2017) report included three private law cases. Of those, in two cases children presented in hospital: one with multiple injuries and one at high risk of death from malnutrition due to neglect. Both cases were non-fatal. It was unclear what happened in the remaining case.

The Harm Panel was also told of children in private law proceedings experiencing multiple physical injuries and being sexually abused (Hunter et al. 2020). Both Hunter et al. (2020) and Women’s Aid (2022) reported accounts of children experiencing stomach aches, likely due to anxiety.

##### Evidence from Australia, Canada, and New Zealand

A Canadian study of an estimated 15,582 children involved in custody or access disputes reported that physical harm was evident in 5.1% of the cases. Of the 61 Australian children in the Carson et al. (2018) study, 26.7% rated their overall mental and physical health as “excellent”, 28.3% as “very good”, 31.7% as “good”, 10.0% as “fair”, and 3.3% as “poor”. In Carson et al. (2022), of the 470 Australian parents and carers, 12.9% said that their child’s health was “excellent”, 20.7% “very good”, 19.8% “good”, 15.2% “fair”, 14.3% “poor”, 0.5% preferred not to say, 5.1% did not know or could not say, and 11.5% selected “other”. Finally, of the 20 Australian court judgements in the Shea Heart (2011) study, one described children having bowel distress and “difficulty with soiling”.

#### 2.7.3 Bottom line results for physical health

Regarding the evidence from England and Wales, accounts of children suffering from severe abuse and neglect, leading to death in some cases, including spite and revenge killings, were provided in the submissions to SCRs in England. These were some of the most complex and severe private law cases. No data on prevalence of such events in private law was available. Physical abuse experienced by children involved in private law was also highlighted in the Harm Panel report. A Canadian study estimated that about 5% of children involved in custody or access disputes experienced physical harm. Some more reassuring evidence came from Australian studies where the samples were not limited to complex cases: the majority of both children and of parents and carers thought that children’s overall mental and physical health was good, very good, or excellent.

## 3. DISCUSSION

### 3.1 Summary of the findings

This review identified 22 studies, including 13 from England and Wales. These studies described a wide range of children’s vulnerabilities, including those pertaining to mental health and wellbeing, engagement with mental health services, behaviour, development, social relationships, learning and education, and physical health.

Most of the available data related to children’s mental health and wellbeing and showed a wide range of issues experienced by these children, including anxiety, depression, and PTSD. Qualitative accounts from both England and Wales and from Australia, Canada, and New Zealand were in broad agreement, but while the quantitative studies from England and Wales were consistent with each other, there was more variability in the data from the comparable countries. Data comparing the incidence of anxiety and depression in children that had been involved in private law proceedings and those in the general population in Wales was available and revealed that children in private law fared worse than peers (Griffiths et al. 2022b). These findings are in line with wider research on children from separated families that does not focus on court involvement (Symonds et al. 2022). Apart from the Griffiths et al. (2022b) study, no other studies comparing children in private law to those in the general population were found.

Less data was available on other types of characteristics and outcomes than mental health and emotional wellbeing. However, the existing evidence points towards many of the children involved in family court proceedings experiencing poor outcomes in other domains. In particular, some of the children in private law proceedings suffered from severe abuse and neglect which in some cases led to death. In some cases, the deaths were classified as spite or revenge killings, demonstrating the extreme levels of violence that some children in private law experience and the need for better risk identification methods and protection mechanisms when it comes to child arrangement disputes. In addition, there were accounts of children having conduct problems, likely attributable to hurt and frustration that they could not find another outlet for. Some children’s social relationships also suffered, with children becoming withdrawn and socially isolated and having difficulty in relating to peers. Little data was available for these types of outcomes.

Overall, the findings point towards the need for interventions to better support children in private law and ensure their safety and wellbeing. Symonds et al. (2022) suggest that at the policy level, a “safety net” needs to be provided to those families that do not have the resources to otherwise ensure that harm to their children is minimised. The authors argue that adequate support provided to children through the separation process can help them to cope better with separation and transition to a different living situation.

### 3.2 Strengths and limitations of the available evidence

The studies included in this review provided a wealth of evidence on the vulnerability and experiences of children involved in family law proceedings due to parental separation, particularly in relation to mental health and wellbeing. The identified qualitative evidence included rich accounts of children, parents, and professionals in the family law system. The available quantitative data was broadly in line with these accounts and with each other, both quantitative and qualitative research pointing towards many of the children experiencing a range of issues across the types of outcomes identified in this review.

Much of the evidence included in this report came from grey literature reports. Only eight relevant academic publications were identified. These academic papers were formally critically appraised and variable quality of the evidence was detected. When it came to quantitative evidence, in most studies, little detail about the children involved was provided, it was not clear how they were sampled, and factors that might be influencing the outcomes, such as existing mental health problems, levels of deprivation, and others, were not always identified and accounted for. Most, but not all, of the qualitative research was deemed to adequately represent participant’s voices, however, in most studies the important topic of the researcher’s own position and potential influence on the research was not addressed. The details of the critical appraisal are available in Section 6.3. Had there been time to formally appraise the grey literature included in this report, we would have likely found the same issues. The level of detail in reporting was variable in the included studies, particularly in terms of reporting the outcomes related to the subscales of questionnaires that concern distinct constructs. Some valuable data was lost because of that.

Finally, the self-selecting samples, particularly in the studies that relied on calls for evidence, mean that families with particularly difficult experiences are likely to be overrepresented. However, their accounts are valuable to draw attention to the existing problems that children involved in family law proceedings due to parents’ separation experience.

### 3.3 Strengths and limitations of this Rapid Review

The review question and the eligibility criteria for this review were developed in consultation with the stakeholders from Cafcass Cymru who requested this work, ensuring that it addresses policy needs, and with a HCRW Evidence Centre PPI member. The main strength of this review is the extensive search for both academic and grey literature: we searched seven bibliographic databases and over 80 websites, screened the studies included in nine existing reviews on similar topics, and searched for literature cited by and citing the relevant studies that we identified during the database searches. This helped us to maximise the amount of identified relevant literature. A limitation of this review is that only academic literature was critically appraised, so no formal judgment about the quality of the included grey literature reports was made. There was also no formal appraisal of the overall body of evidence.

### 3.4 Implications for policy and practice

Despite there being twice as many children in private law proceedings compared to public law, they receive little attention in policy. The findings of this review show that many children in private law cases experience issues with mental health and emotional wellbeing and that these issues are serious and wide-ranging. However, little support is available for this vulnerable group of children, especially while the proceedings are ongoing. Therefore, the implications of this review go beyond the Family Justice system. The findings indicate a need for a broad public health response to create a safety net around these children, which may include schools, GPs, and mental health services. A coordinated response can help to identify vulnerable children and provide appropriate and timely support. The availability of support during ongoing court proceedings is important because many children in private law cases are in acute distress, so having to wait until the cases is concluded, especially given the protracted nature of some cases, may mean that such distress is prolonged and exacerbated.

### 3.5 Implications for future research

While we identified a wealth of evidence that characterises children who are or have been involved in family law proceedings due to parental separation, most of it comes from grey literature, suggesting a disconnect between academic research and real-world policy needs. There is a need for more high-quality studies of the outcomes of such children, particularly long-term, in order to better inform policy decisions. Research comparing short- and long-term outcomes would also be helpful to understand how support needs may change.

Much of the identified research was informed by parents and professionals. While there was a general agreement between the accounts provided by children or adults who had been involved in family law proceedings as children and parents and professionals, it is important to make sure that children are provided with sufficient opportunities to have their views and accounts of their own experiences considered in matters that concern them.

Even within the broad themes identified in this review, there were significant gaps. The only direct comparison with children in the general population was made in data related to anxiety and depression (Griffiths et al. 2022b). While the available evidence indicates that children in private law experience a wide range of negative outcomes across different domains, it is unclear how their support needs differ from children in the general population. Therefore, there is a need for more high-quality comparative studies to better understand the specific support needs of children in private law. More robust data on the prevalence of poor outcomes in children in private law proceedings would also be helpful. Finally, research is needed on the outcomes of children in private law compared to children from separating or separated families whose parents do not go to court over child arrangement disputes to better understand whether and how private law proceedings exacerbate children’s experiences, as currently suggested by qualitative accounts identified in this review.

##### 3.6 Economic considerations*

Most private law cases in Wales concern Child Arrangement Orders (CAOs) for a single child aged between one and nine years old (Cusworth et al. 2020). A third (33%) of the mothers and 29% of the fathers making private family law applications in Wales are from the most deprived quintile (Cusworth et al. 2020). Since 2013, legal aid is not available for private law proceedings except some cases, particularly those involving domestic abuse (Hunter et al. 2020). In face of the lack of resource, children and parents in cases not entitled to legal aid are expected to accommodate themselves to contact and to bear direct and indirect costs (Hunter et al. 2020). Given the prevalence of socioeconomically deprived individuals in private family law cases, this acts as a compounding economic challenge further to the disruption of the case itself.

It is estimated that between 49% and 62% of child arrangement and contact cases involve allegations or findings of domestic abuse (Hunter et al. 2020). There is an acknowledgement that safeguarding measures to conduct risk-assessments and putting interventions in place to ensure child arrangements are safe are required, however a lack of financial resources in the private law system restricts their implementation (Hunter et al. 2020). Resource shortages affect the whole private law system, but familial domestic abuse cases may be the most at-risk given they are likely to be more resource-intensive to address than non-abuse cases (Hunter et al. 2020). The Ministry of Justice recommends an appropriate model of specialist domestic abuse advocacy and support services be evaluated in terms of its effectiveness and cost-effectiveness of supporting alleged victims and alleged perpetrators (Hunter et al. 2020). More broadly, domestic abuse cases incur economic costs of £66 billion per annum in the UK. However, these figures did not include the cost of harms to children or the costs of financial abuse or coercive and controlling behaviour (Oliver et al. 2019).

In terms of children’s long-term outcomes, children who experience parental separation before the age of 15 can experience an associated 46% reduction in lifetime net wealth compared to those who do not experience it (Lersch & Baxter 2021). However, it is unclear whether there is a difference in lifetime net wealth of those children whose parents used courts and those whose parents did not.

## Data Availability

All data produced in the present study are available upon reasonable request to the authors

## Acknowledgements

The authors would like to thank Matthew Pinnell and Anna Sinclair (Cafcass Cymru) and Olivia Gallen (HCRW Evidence Centre public involvement member) for their time, expertise, and contributions during stakeholder meetings in guiding the focus of the review and interpretation of findings. We would also like to thank Hannah Furness and Asha Mahamed for their contribution during the early stages of this review.

## 4. RAPID REVIEW METHODS

The protocol for this review was registered on the OSF website and is available through the following link: https://osf.io/7ngza/. Some deviations from the protocol were made in describing the methodology of the review to clarify the inclusion of descriptive studies in addition to analytic studies. For the same reason, the review question was also modified to include the word “characteristics”.

### 4.1 Eligibility criteria

The eligibility criteria were developed in consultation with the stakeholders from Cafcass Cymru that requested this review and a HCRW Evidence Centre public involvement member. They are available in Table 2.

**Table 2:** Eligibility criteria.

|  | **Inclusion criteria** | **Exclusion criteria** |
| --- | --- | --- |
| **Population** | Children aged <18 at the time of court proceedings | • Adult family members<br>• Professionals involved in court proceedings |
| **Exposure** | Private family law proceedings due to parental separation | • Public family law proceedings due to parental separation<br>• Other legal proceedings that are not due to parental separation<br>• Family conflict without parental separation<br>• Parental separation without court involvement |
| **Outcomes** | Description of the characteristics or short-and long-term outcomes in the following domains:<br>• Mental health and wellbeing, including but not limited to:<br>○ Mental health conditions<br>○ Engagement with CAMHS<br>○ Stress<br>○ Emotional regulation<br>○ Social relationships<br>○ Behaviour<br>• Education<br>• Physical health |  |
| **Context** | Private family law proceedings |  |
| **Study design** | Primary quantitative, qualitative, or mixed methods studies (analytic or descriptive), including but not limited to:<br>• Longitudinal studies<br>• Cohort studies<br>• Cross-sectional studies<br>• Interviews<br>• Focus groups | Secondary research |
| **Countries** | • England and Wales (countries of primary interest)<br>• Australia, Canada, New Zealand (countries with comparable legal systems) |  |
| **Language of publication** | English |  |
| **Publication date** | ≥2001 |  |
| **Publication type** | • Published academic literature<br>• Grey literature | • Letters<br>• Commentaries<br>• Editorials<br>• Conference abstracts<br>• Opinion pieces |
| **Other factors** | Literature that reports data obtained directly from children and/or from parents, guardians, or relevant professionals (e.g., teachers, social workers, family court advisors etc) is included. |  |

### 4.2 Literature search

A number of search strategies, including database searches, review unpicking, citation searching, and website searches, were used and are reported in detail in this section.

#### 4.2.1 Database searches

The following databases were searched in July 2024 for literature published since 2001: PsycINFO via Ovid, MEDLINE via Ovid, Scopus, the Web of Science, Social Science Database via ProQuest, Sociology Collection via ProQuest, ERIC via ProQuest. A range of terms related to children and family law was used in different combinations to maximise the sensitivity of the searches. The searches were limited to the English language. Where possible, country limits were applied. The full search strategies for each database are available in Appendix 1.

#### 4.2.2 Identifying literature from existing reviews

Nine reviews, published in ten documents (Allen 2014, Barnett 2020, Birnbaum & Saini 2012a, Birnbaum & Saini 2012b, Doughty et al. 2018, Giovannini 2011, Kelly & Emery 2003, Nuffield Family Justice Observatory 2021, Roe 2021, Sands et al. 2017), which had been identified during informal preliminary searches, during database and grey literature searches, and through other reviews, were unpicked. Only records warranting further investigation were exported, i.e. not those from ineligible countries, published before 2001, or excluded based on the title.

#### 4.2.3 Citation searching

Citation searching was undertaken using a combination of the Web of Science and Scopus databases. Five seed articles that we included after being identified during the database searches were used. Both backward and forward citation searching was conducted, with the references found from each seed article uploaded to Endnote. The identified references were deduplicated against each other and against the existing library of references identified during the database searches.

#### 4.2.4 Website searches

A list of websites of relevant UK-, Australia-, Canada-, and New Zealand-based government, research, and third sector organisations was identified through known literature, web searches, snowballing, and the review team’s prior knowledge. The list is available in Appendix 2. The searches were conducted between June and August 2024. Each website was searched by at least one reviewer using keywords and/or by reviewing lists of publications where those were available on the websites. After deduplication, potentially relevant documents were downloaded for further review.

#### 4.2.5 Other literature identification methods

An internal study (Harold 2013) was supplied by the stakeholders from Cafcass Cymru for the purposes of this review.

### 4.3 Study selection process

The flow of citations identified through each method through the review process is reported in the PRISMA flow diagram (Page et al. 2021) in Figure 1.

**Figure 1:**
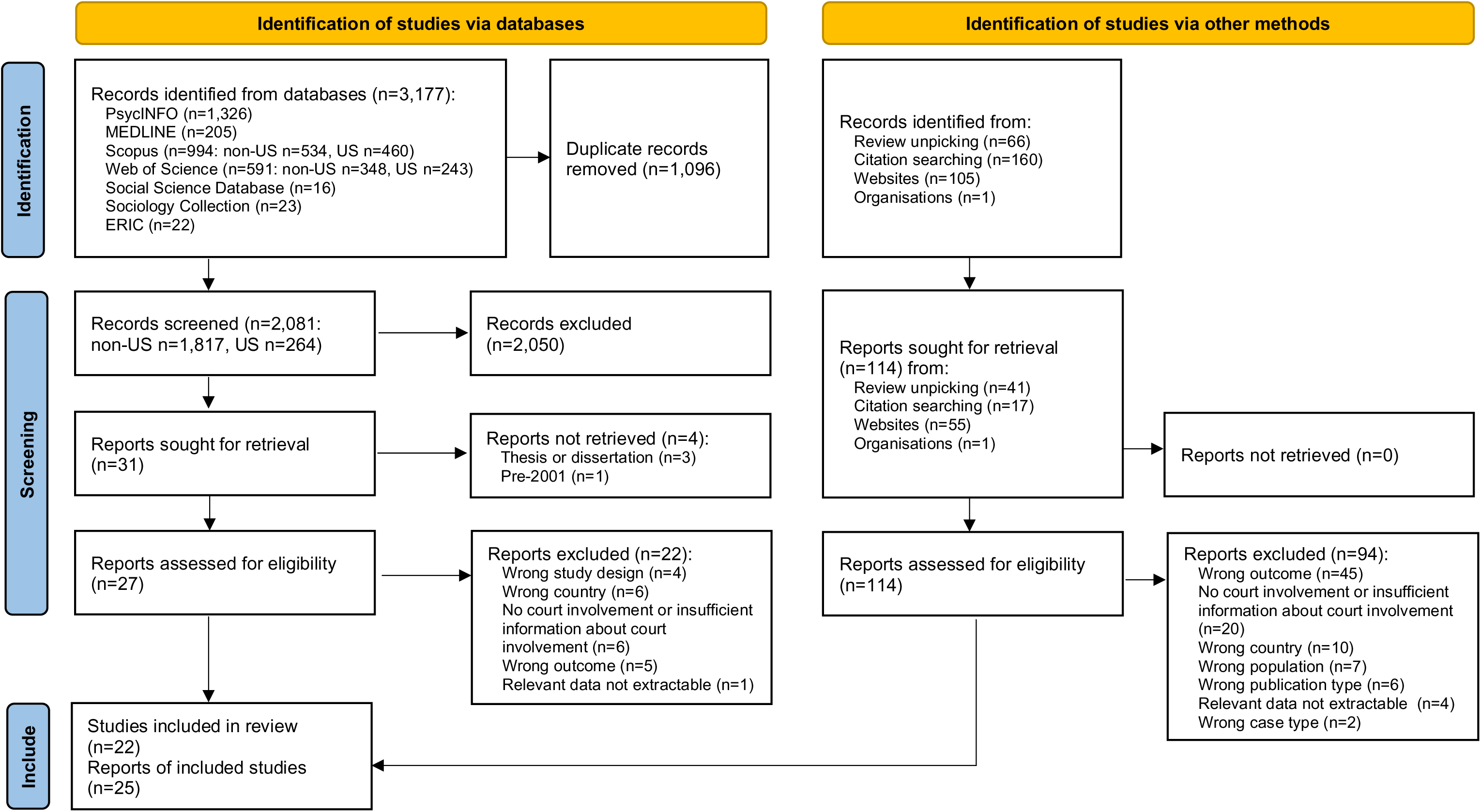
PRISMA flow diagram.

#### 4.3.1 Identified database records

The records identified in Scopus and the Web of Science as originating in the US were screened by a single reviewer. The rest of the records were independently screened by two reviewers based on the information provided in the titles and abstracts, with disagreements resolved by a third reviewer where necessary. Records that appeared to meet the eligibility criteria and those where a decision could not be made based on the information provided in the title and abstracts alone progressed to full-text screening. A decision algorithm based on the eligibility criteria was developed and piloted on two of the studies. Full-text screening was performed by two reviewers independently using the decision algorithm, with any disagreements resolved by a third reviewer. The list of studies excluded during full-text screening with exclusion reasons is available in Appendix 3.

#### 4.3.2 Literature identified from existing reviews

The identified references were deduplicated against those already found during the database searches and previous grey literature searches. The resulting additional references were a mix of academic and grey literature documents. These references were screened by a single reviewer based on the information provided in the titles and abstracts, and records that appeared to be eligible, or where a decision could not be made based on the titles and abstracts alone, progressed to full-text screening. The full texts were scanned by a single reviewer and excluded if the study was performed in an ineligible country. The rest of the full texts were screened independently by two reviewers, with a third reviewer arbitrating conflicts where necessary.

#### 4.3.3 Literature identified through citation searching

The identified references were screened based on the information provided in the titles and abstracts by a single reviewer. References that appeared to be eligible, or where a decision could not be made based on the titles and abstracts alone, were independently screened at full text by two reviewers.

#### 4.3.4 Literature identified through website searches

All of the downloaded documents were initially scanned by a single reviewer and irrelevant documents were excluded. Where a document appeared to be relevant or where a decision could not be made by brief scanning, it proceeded to the next stage of review and was independently screened by two reviewers.

### 4.4 Data extraction

Relevant data from the identified studies were extracted into a table which had been piloted on two studies of different designs first by a single reviewer in conversation with another reviewer. The following data were extracted from each study by a single reviewer and checked for accuracy and completeness by another reviewer: study aim, study design, dates of data collection, data collection methods, quality assessment tool and rating, who the informants were, sample size, participants’ characteristics within family law, participants’ demographics, outcomes of interest and outcome measures, relevant findings.

### 4.5 Study design: classification

This review included studies published in academic articles and grey literature reports, using quantitative methods, qualitative methods, or mixed methods. Where only quantitative or qualitative data were relevant to the review question and extracted from a mixed method study, the study was recoded as quantitative or qualitative accordingly. Only studies published in academic articles were classified according to study design, which was done for the purposes of selecting an appropriate critical appraisal checklist. No formal study identification algorithm was used. Instead, two reviewers classified the articles in conversation with each other.

### 4.6 Quality appraisal

Due to the time limitations of this review, only academic articles were critically appraised. Critical appraisal was performed by one reviewer and checked by another, with any disagreements resolved through discussion. Cross-sectional studies were assessed using the JBI Critical Appraisal Checklist For Analytical Cross Sectional Studies (Moola et al. 2020). For qualitative studies, the JBI Critical Appraisal Checklist For Qualitative Research (Lockwood et al. 2015) was used. Finally, the cohort study was assessed using the JBI Critical Appraisal Checklist For Cohort Studies (Moola et al. 2020). The full record of the critical appraisal is provided in Section 6.3.

#### 4.7 Synthesis

Data from the included studies were synthesised narratively using a series of thematic summaries. For readability, the data were grouped in seven sections, not three as anticipated at the protocol development stage.

### 4.8 Assessment of body of evidence

No formal assessment of the overall body of evidence was performed within the limitations of this rapid review, however, the dimensions included in the Grading of Recommendations, Assessment, Development and Evaluation (GRADE) approach (Schünemann et al. 2023) and the Confidence in the Evidence from Reviews of Qualitative research (GRADE-CERQual) approach (Lewin et al. 2018) were considered. As such, when narratively describing the overall body of quantitative evidence, where possible, we reflected on the risk of bias, imprecision, inconsistency, and indirectness of the evidence as well as possible publication bias; methodological limitations, coherence, adequacy of data, and relevance to the research question were considered in relation to the qualitative evidence.

## 5. EVIDENCE

### 5.1 Search results and study selection

A visual representation of the flow of study selection throughout the review can be found in Figure 1. A total of 22 studies (reported in 25 publications) were included in the review. If only a quantitative or qualitative part of a mixed methods study was relevant to the review question and therefore extracted, the study was classified as quantitative or qualitative accordingly. As a result, of the 22 included studies, ten were classified as quantitative methods, eight as qualitative, and four as mixed method. Out of the eight academic studies, four were classified as cross-sectional (Black et al. 2021, Bream & Buchanan 2003, Brown 2002 “The First Study”, “The Magellan Study”), three as qualitative (Bailey et al. 2011, Darlington 2001, Shea Hart 2011), and one as cohort (Griffiths et al. 2022b).

### 5.2 Data extraction

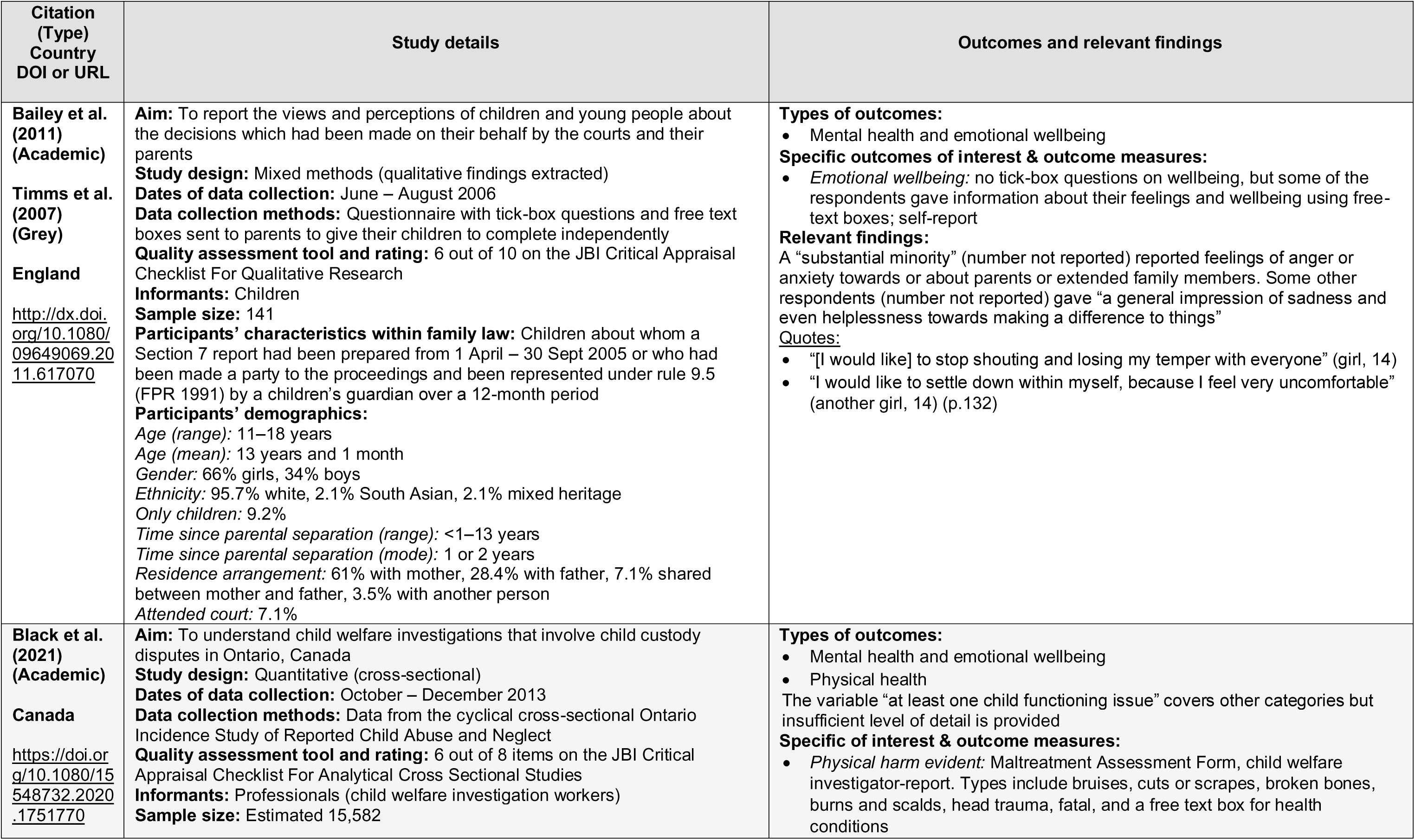
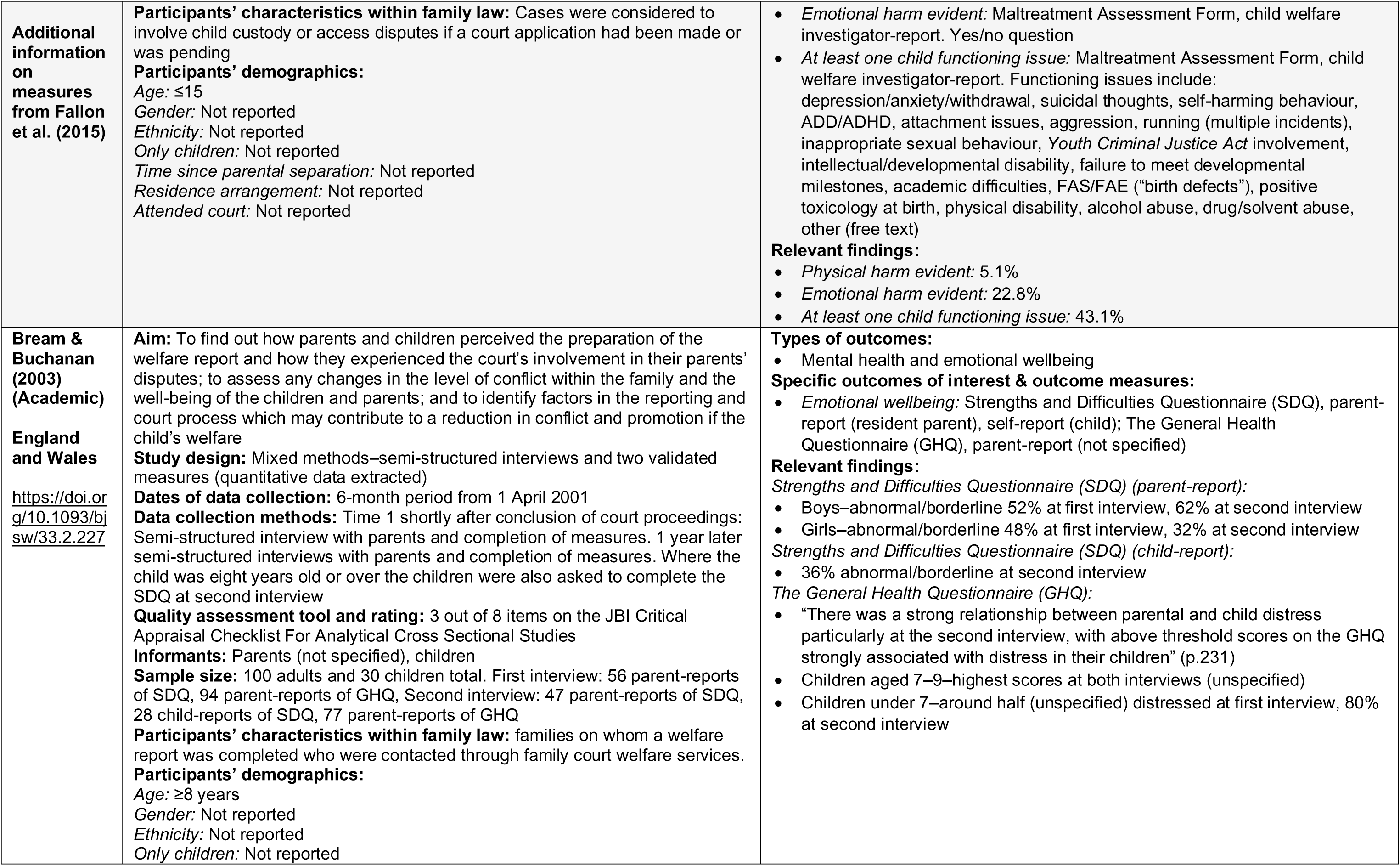
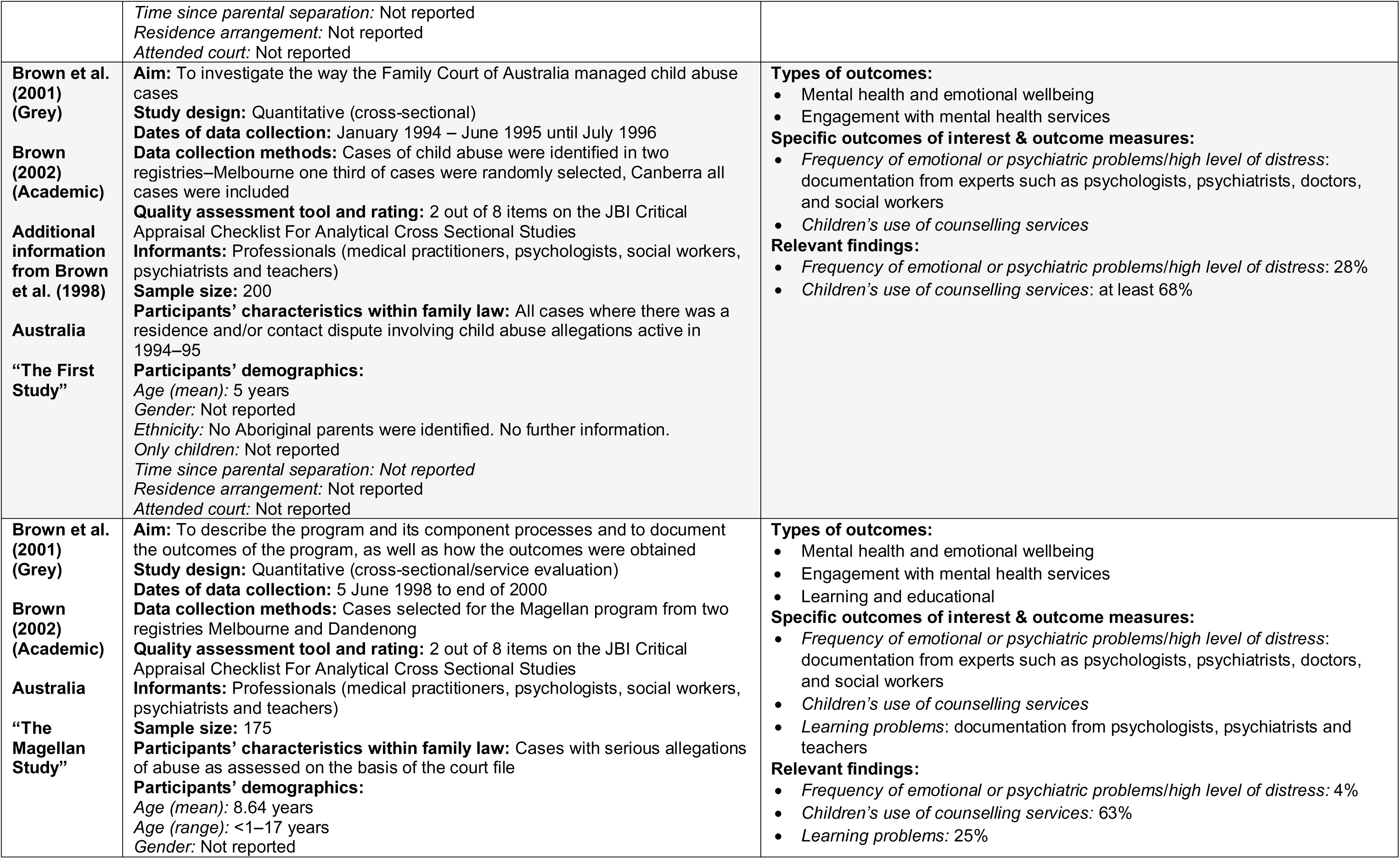
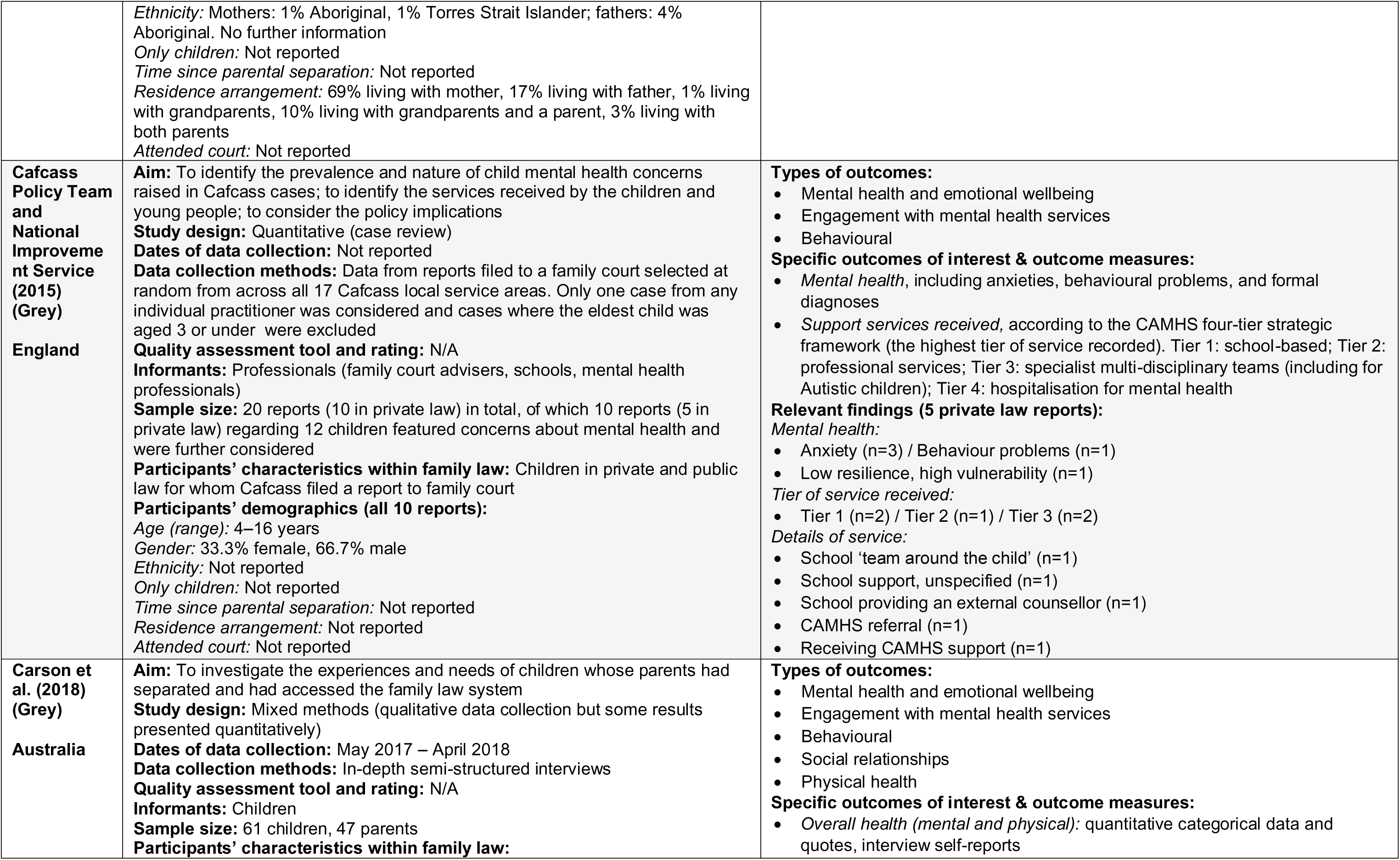
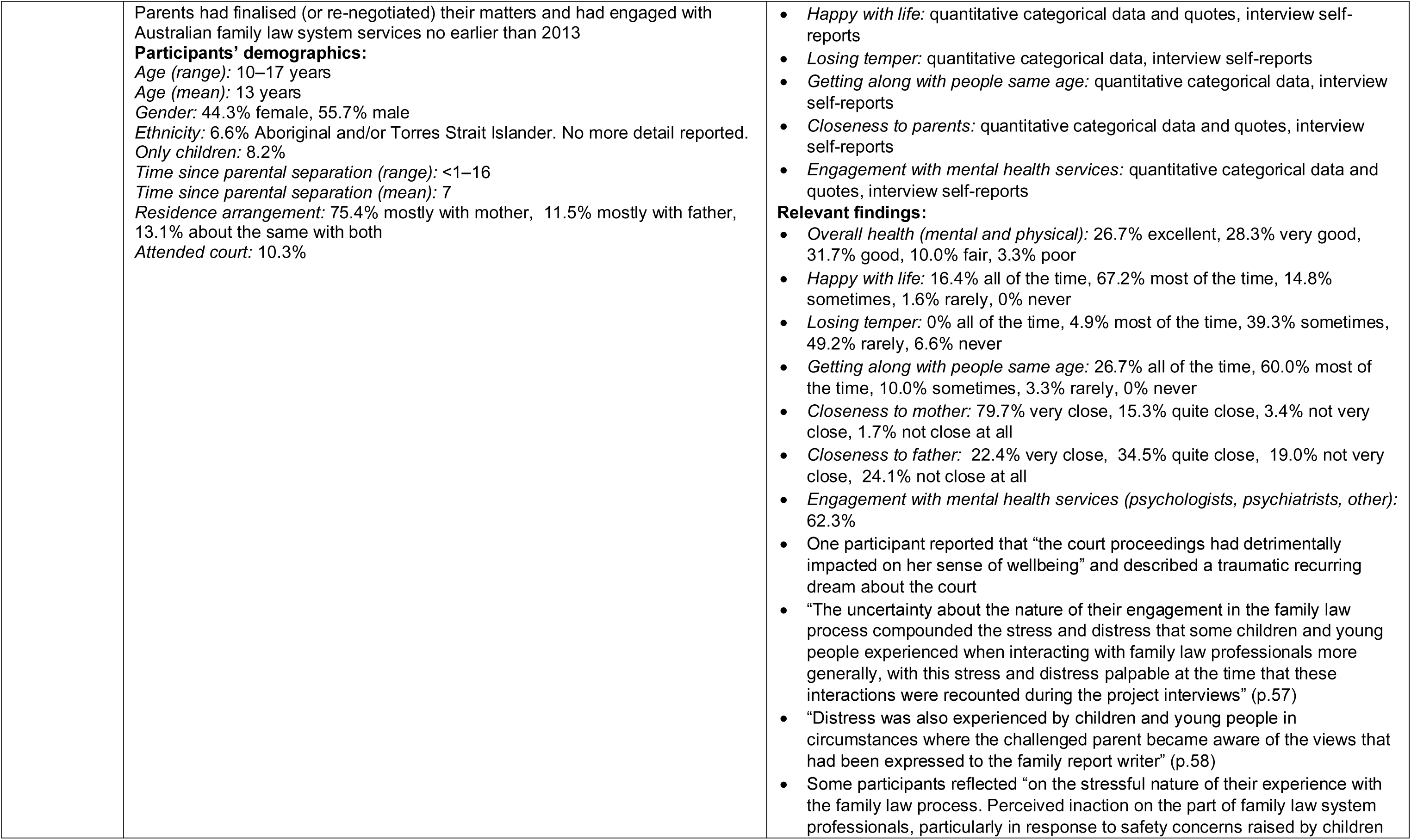
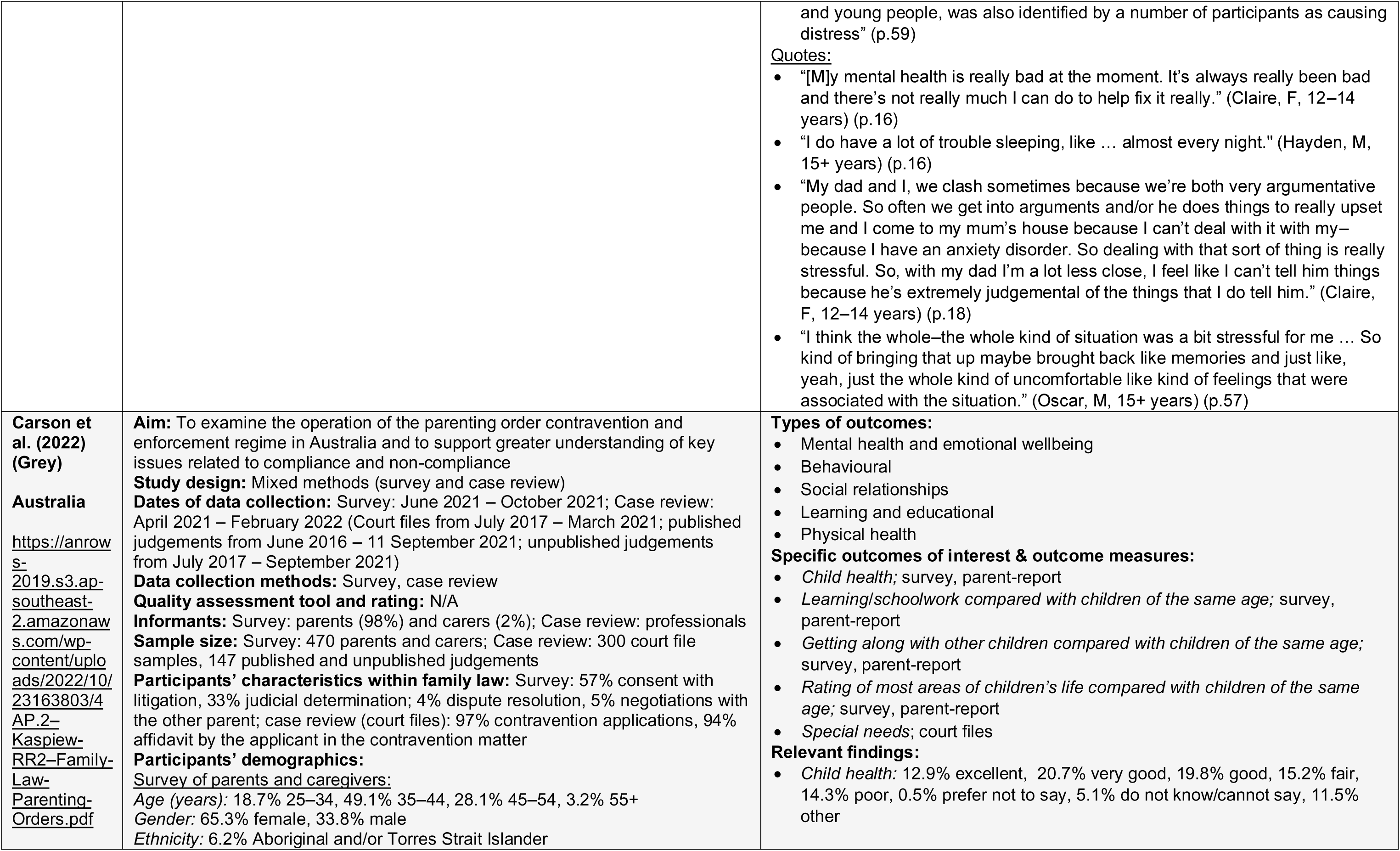
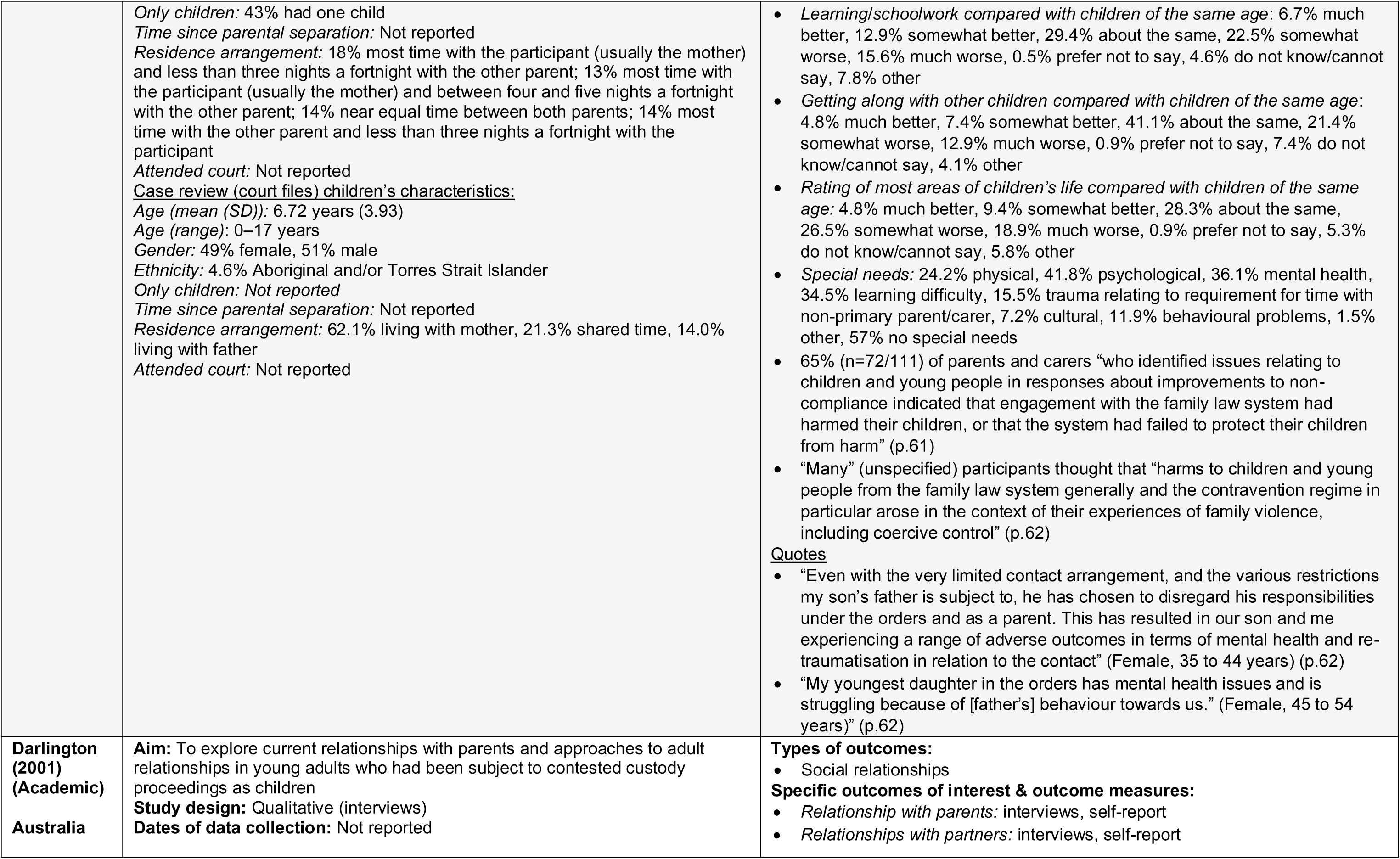
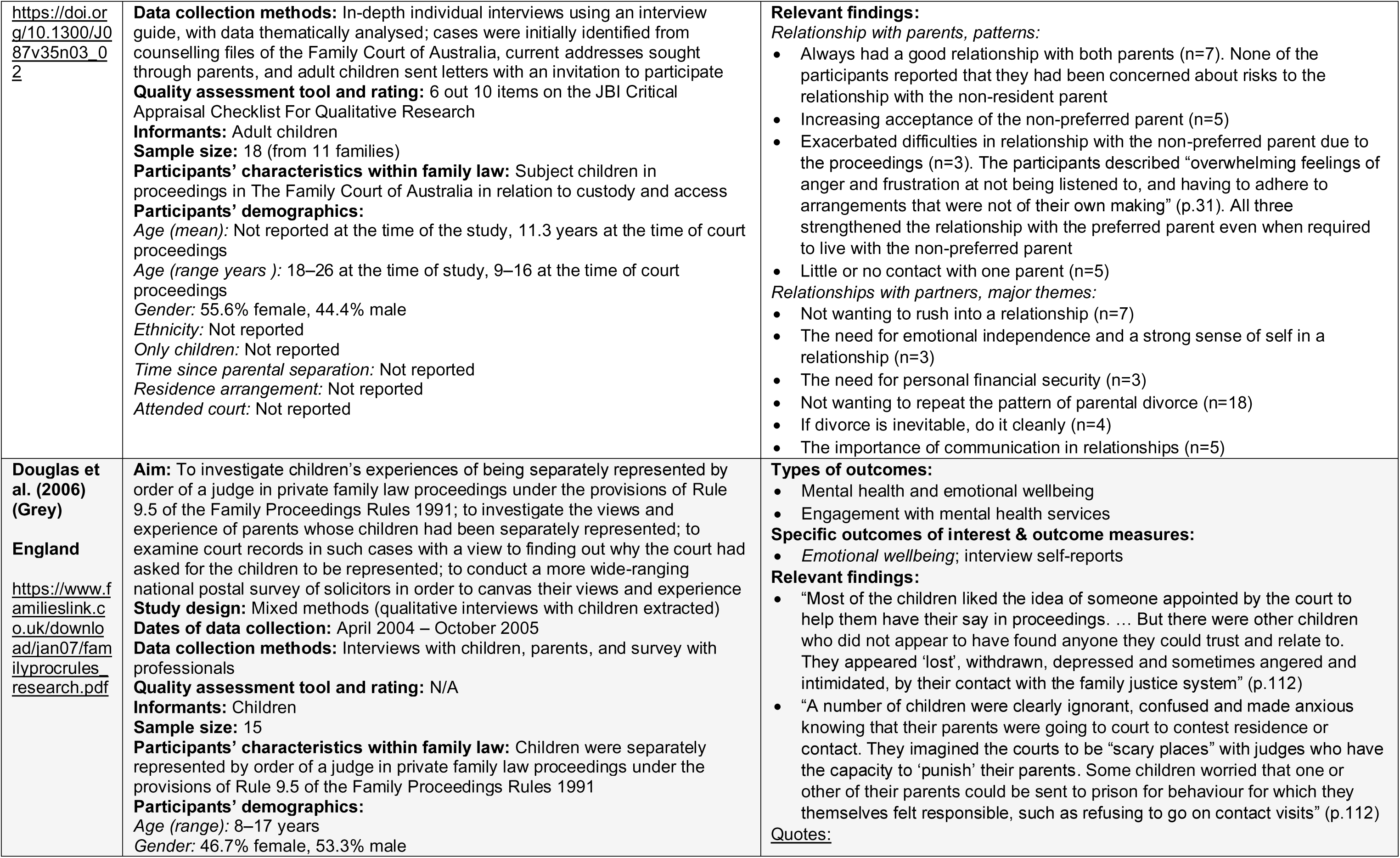
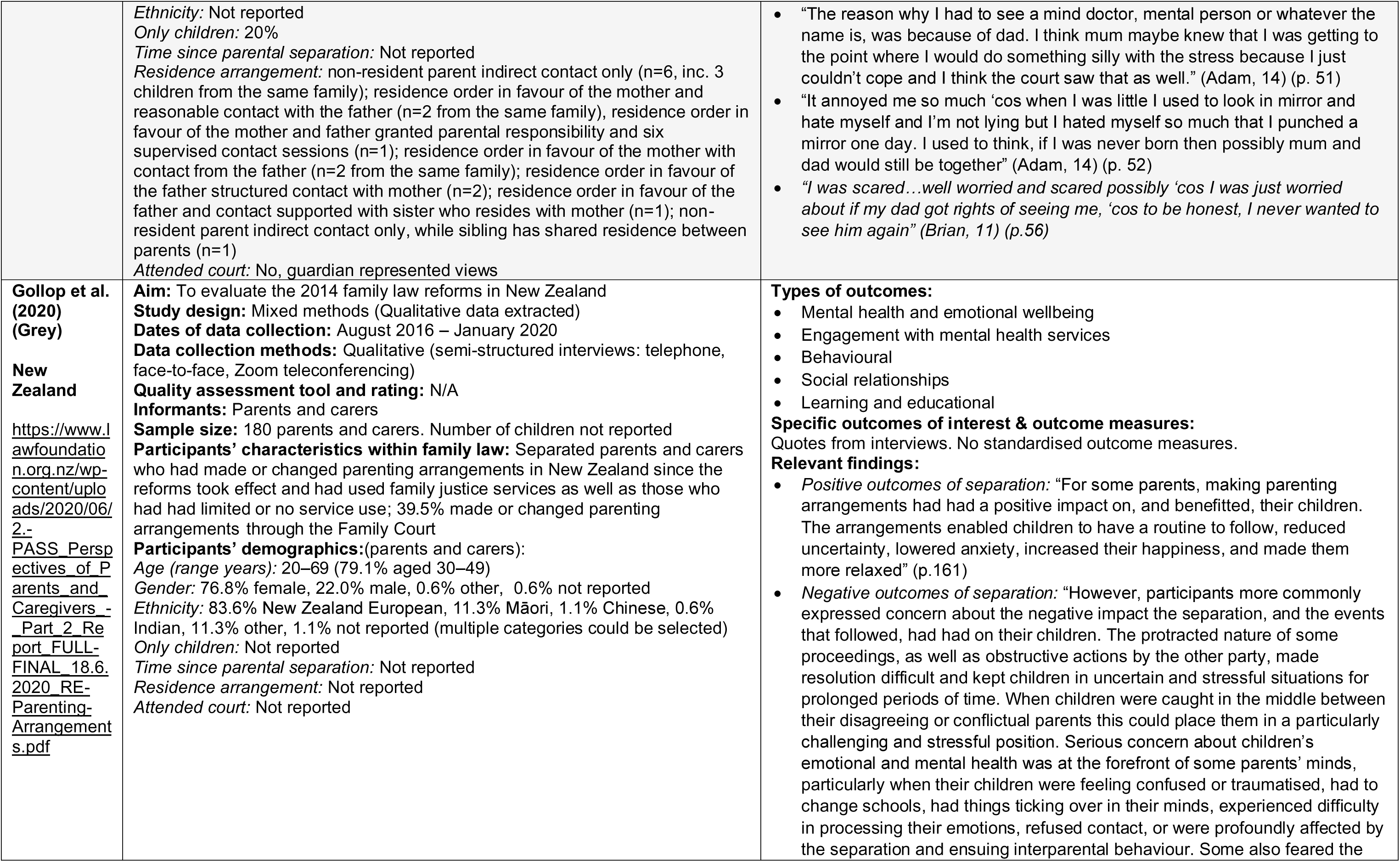
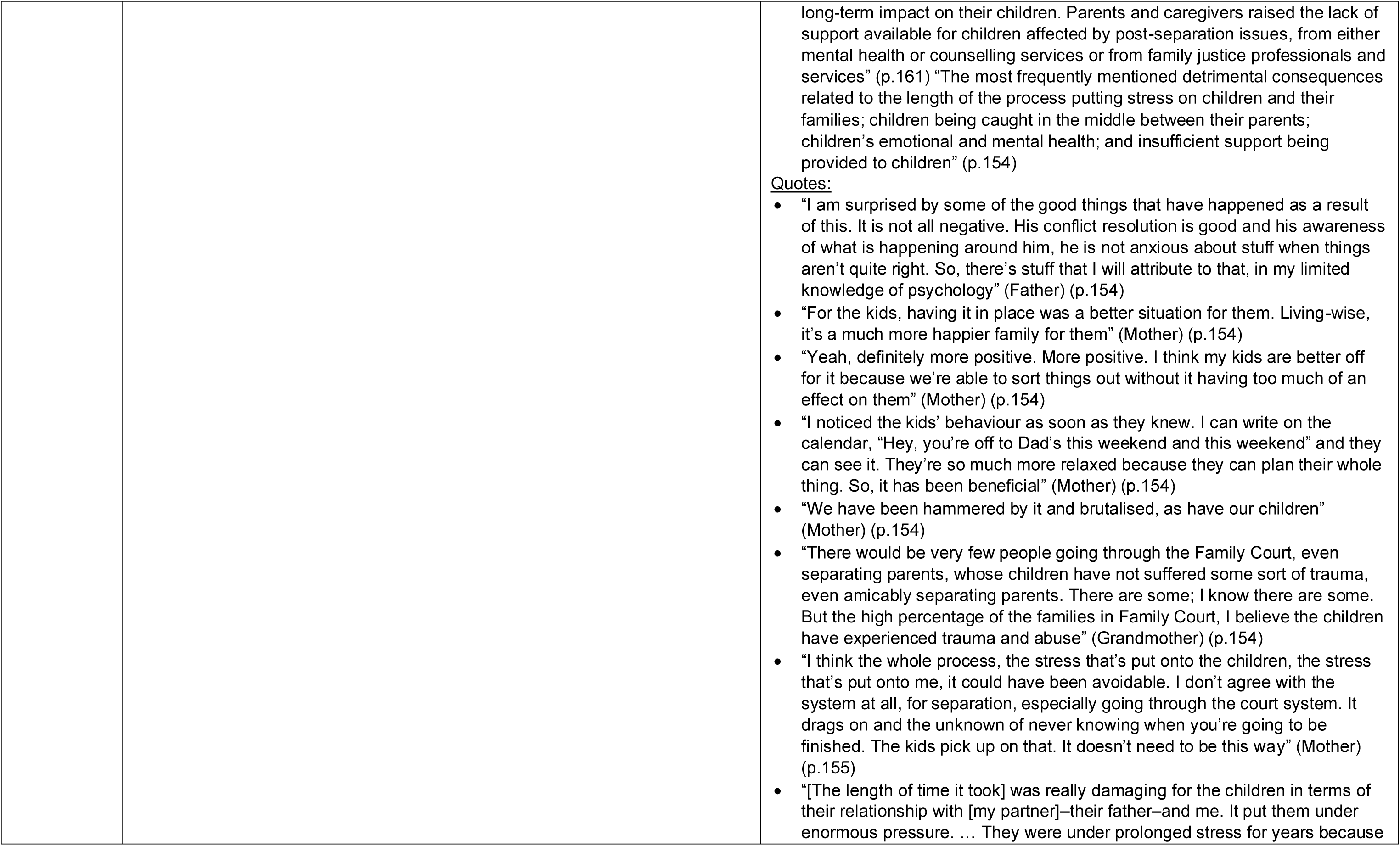
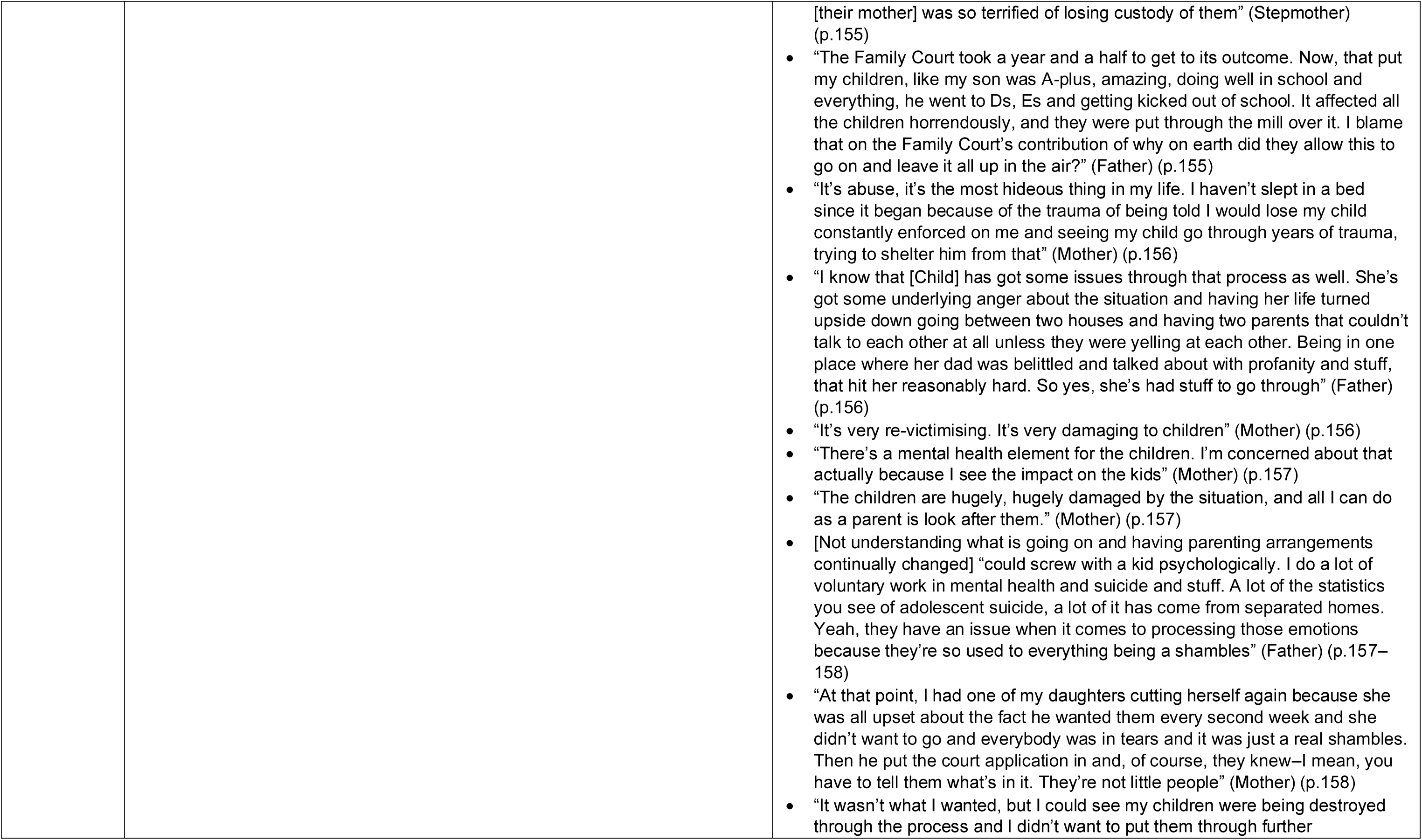
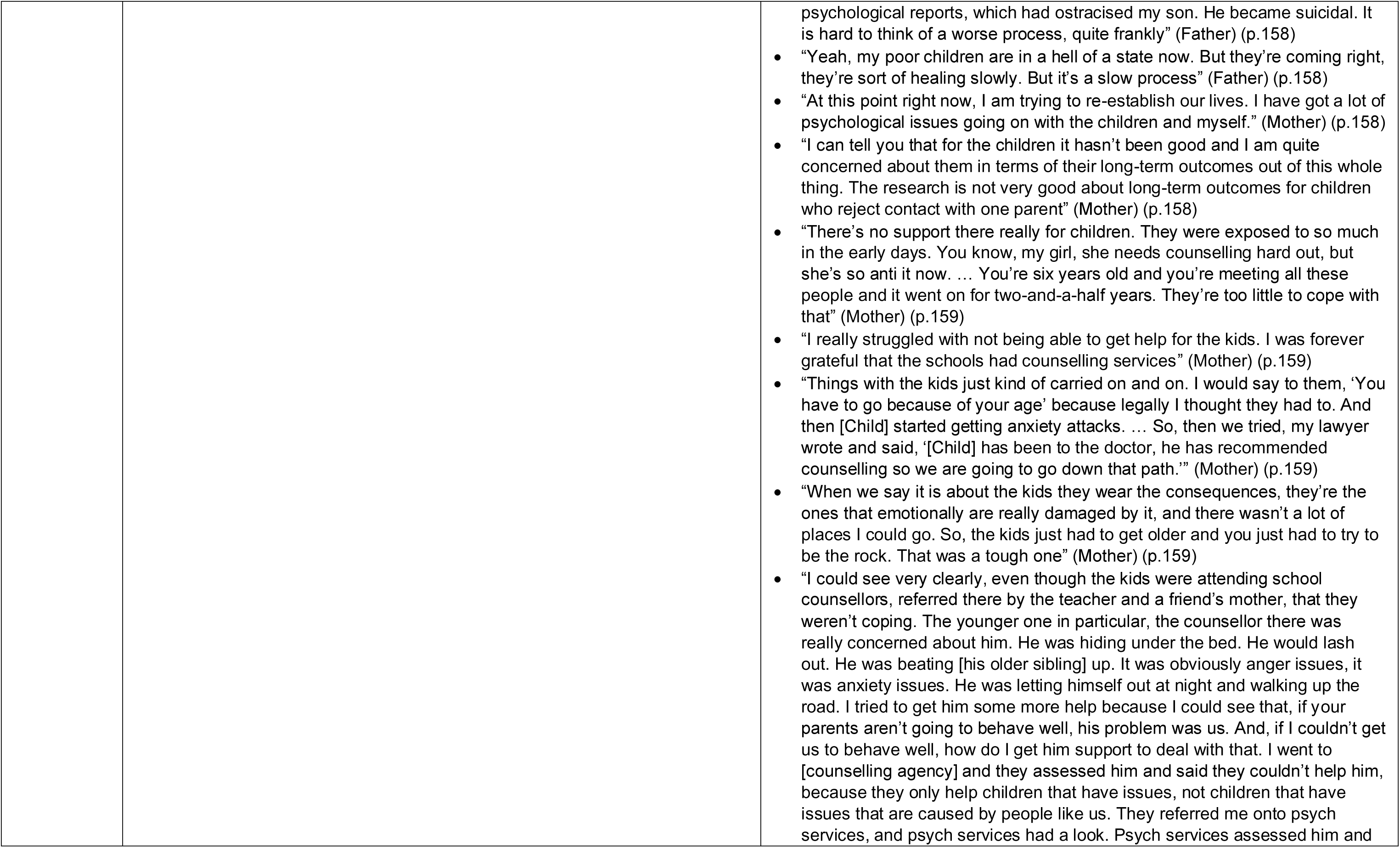
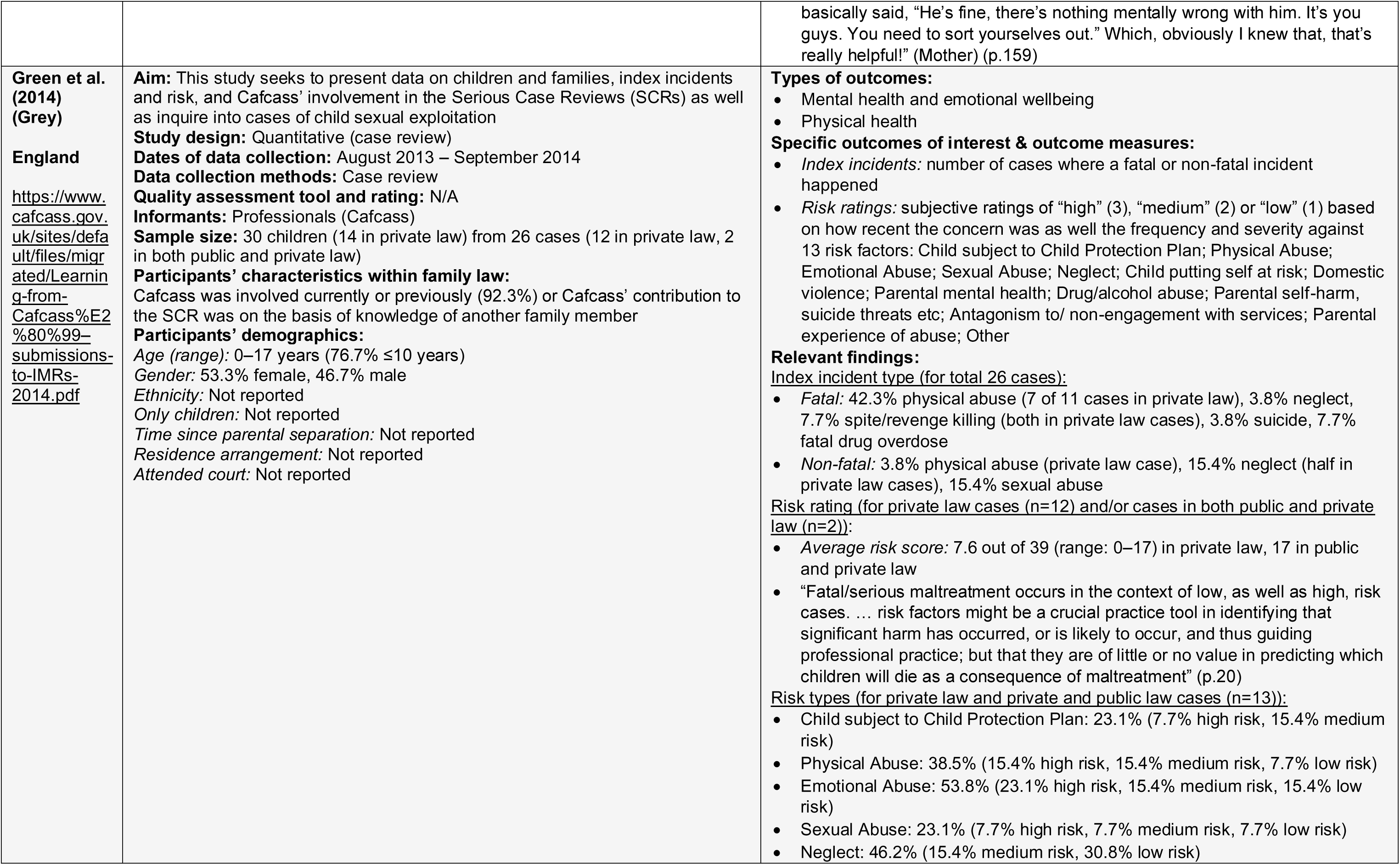
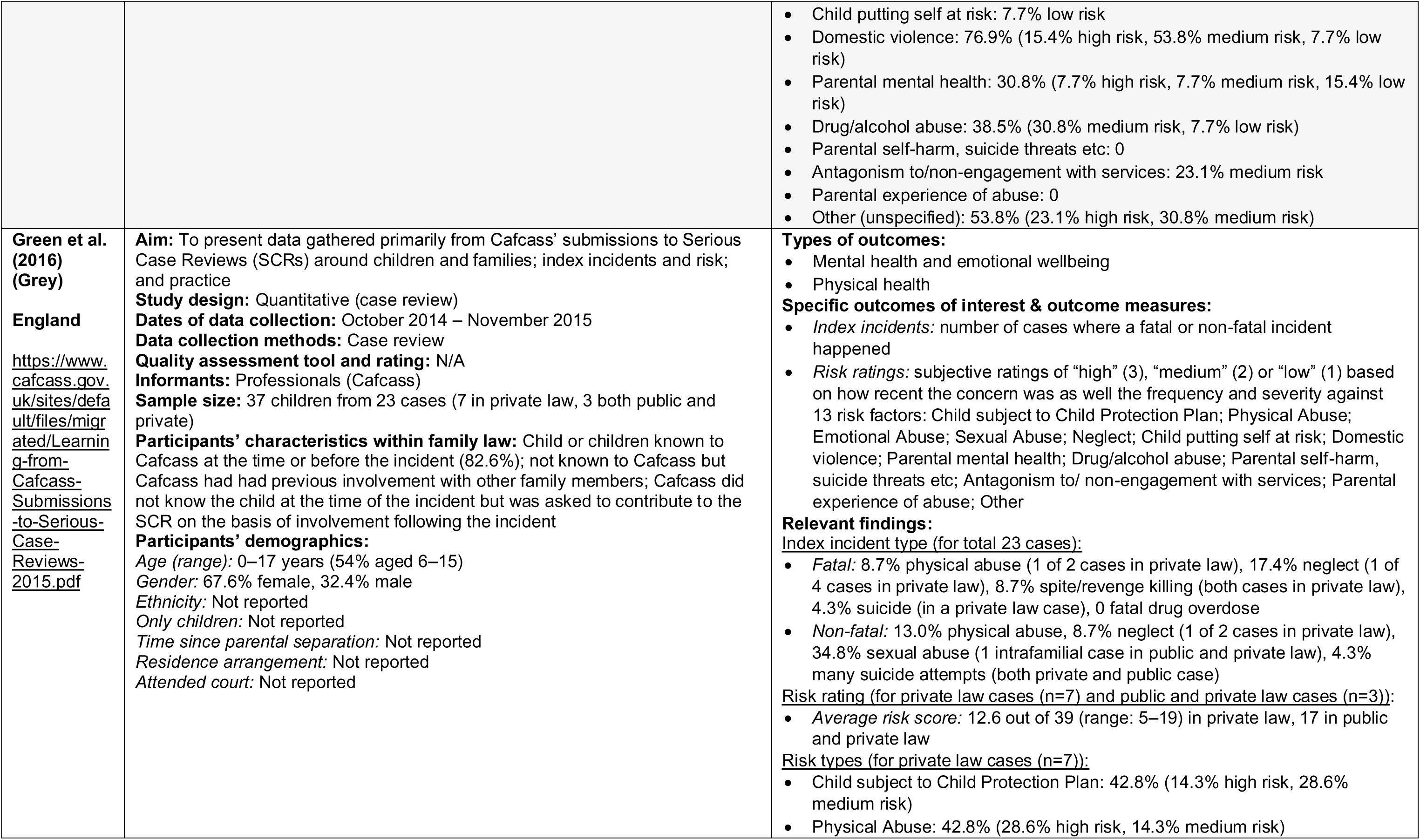
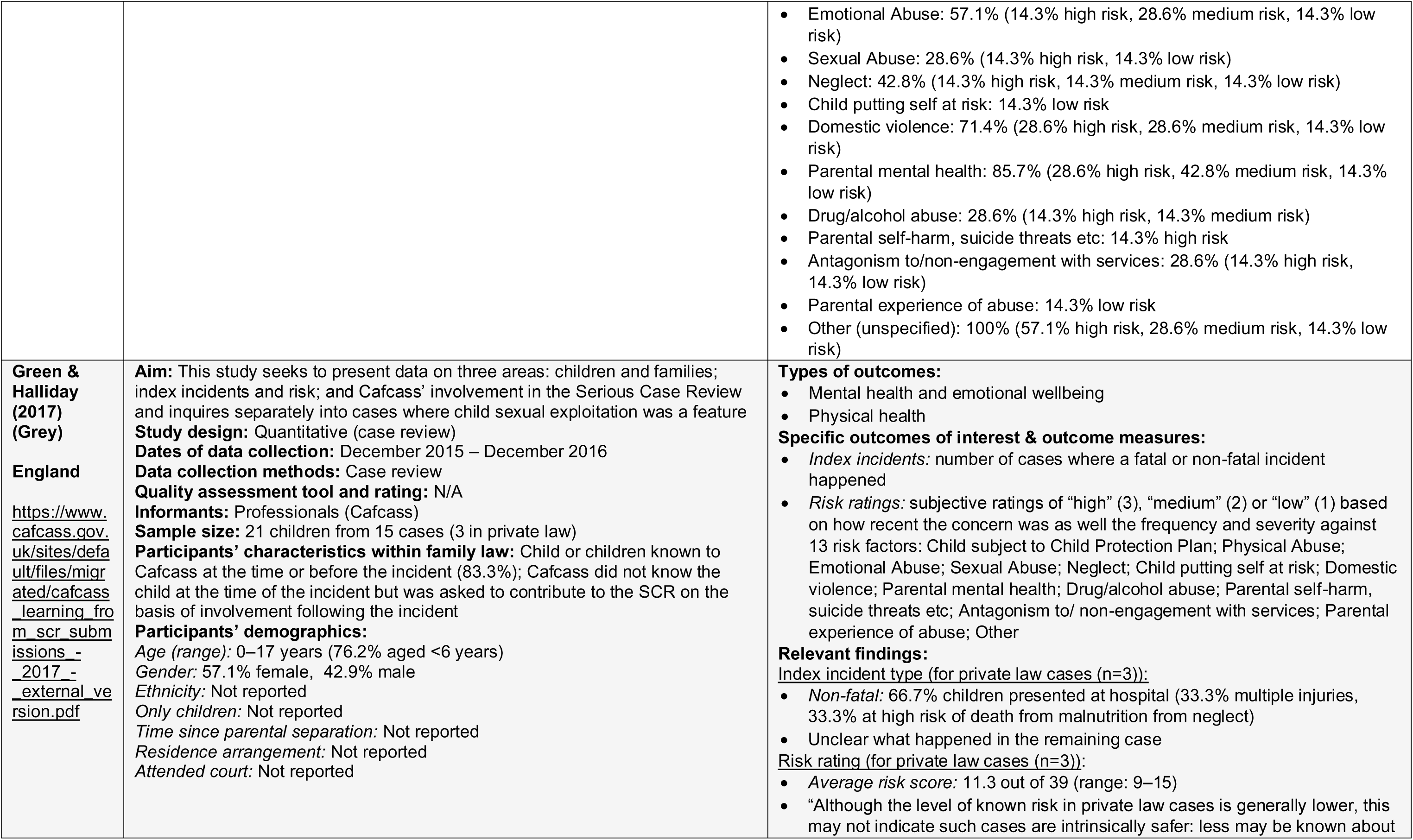
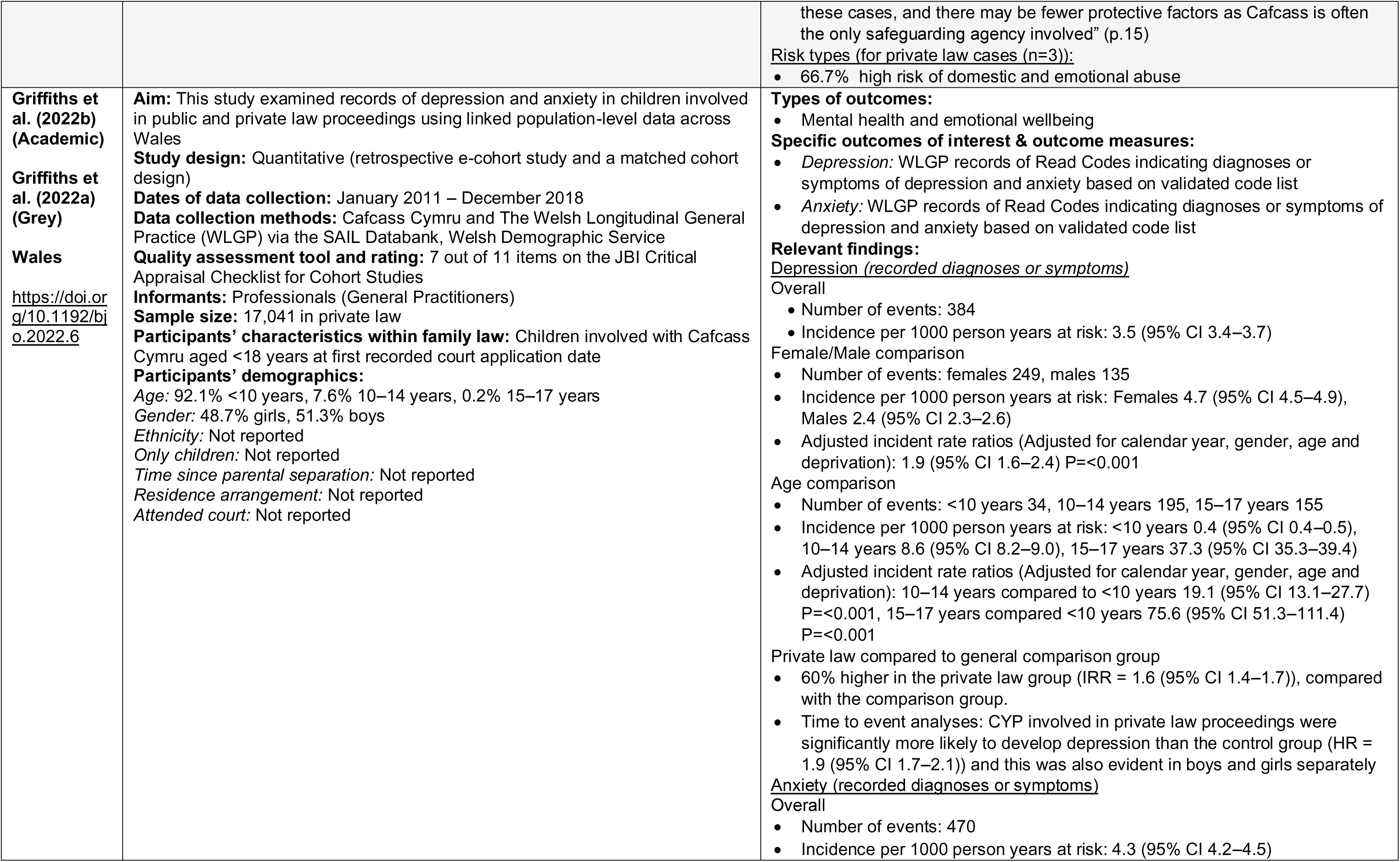
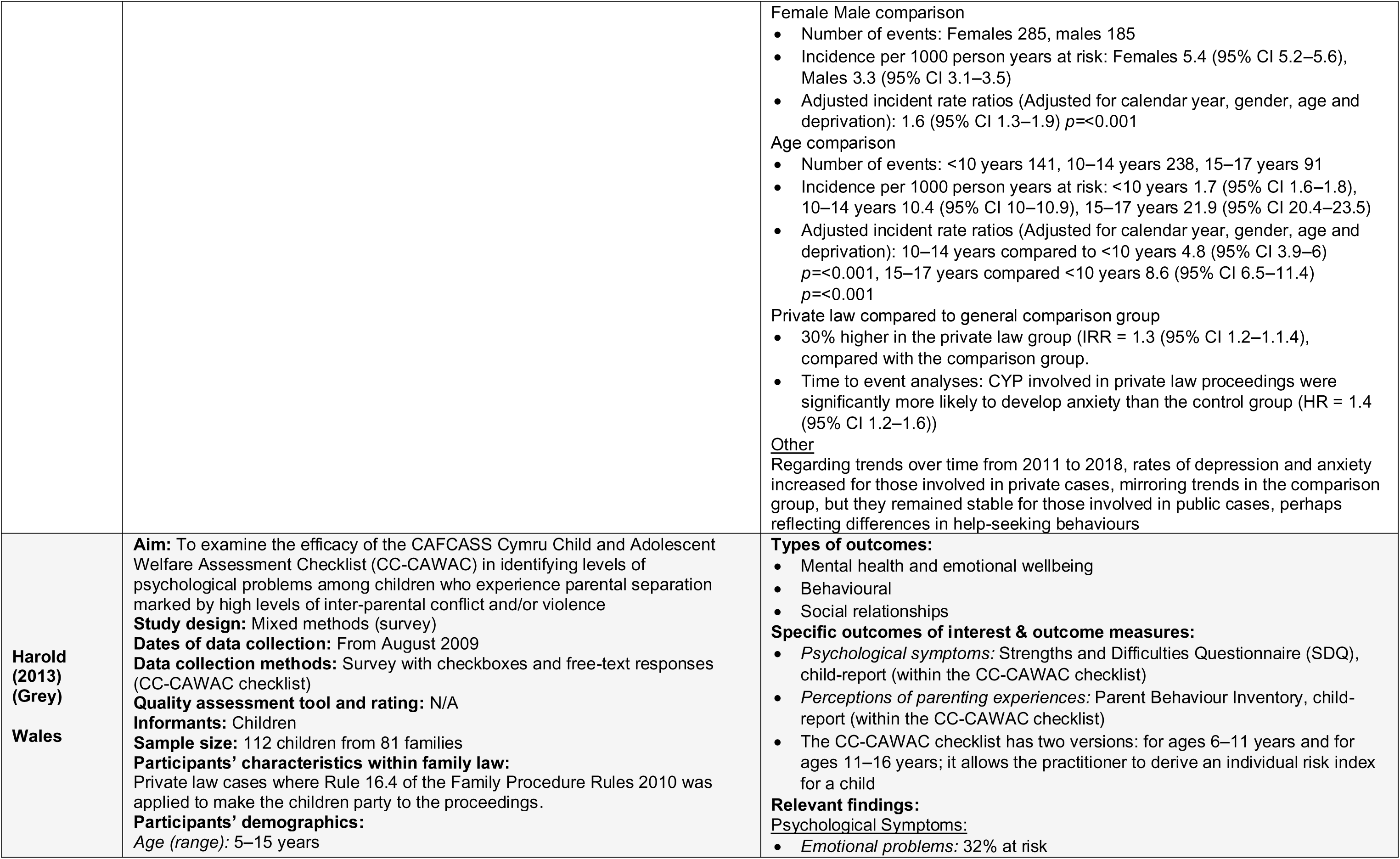
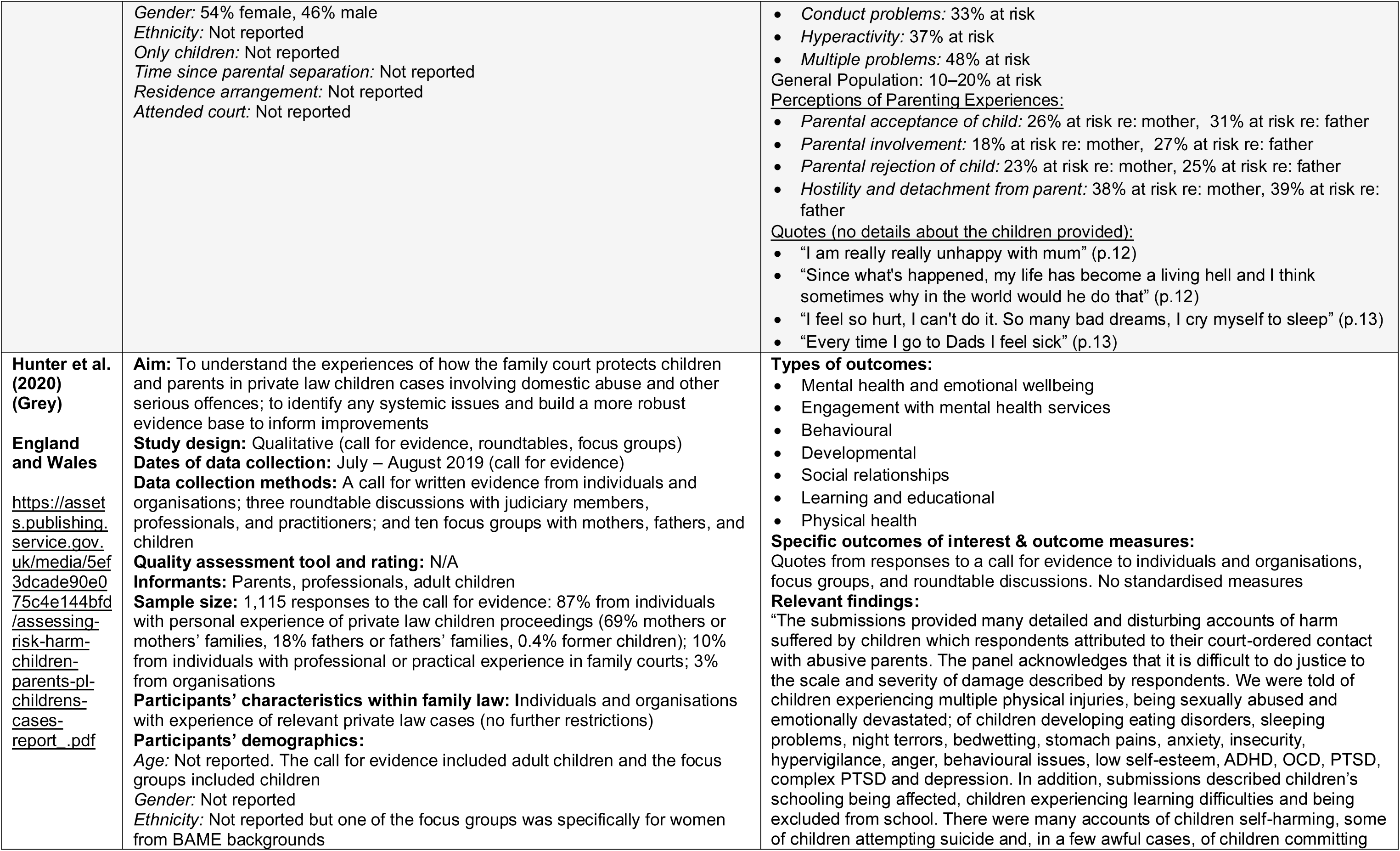
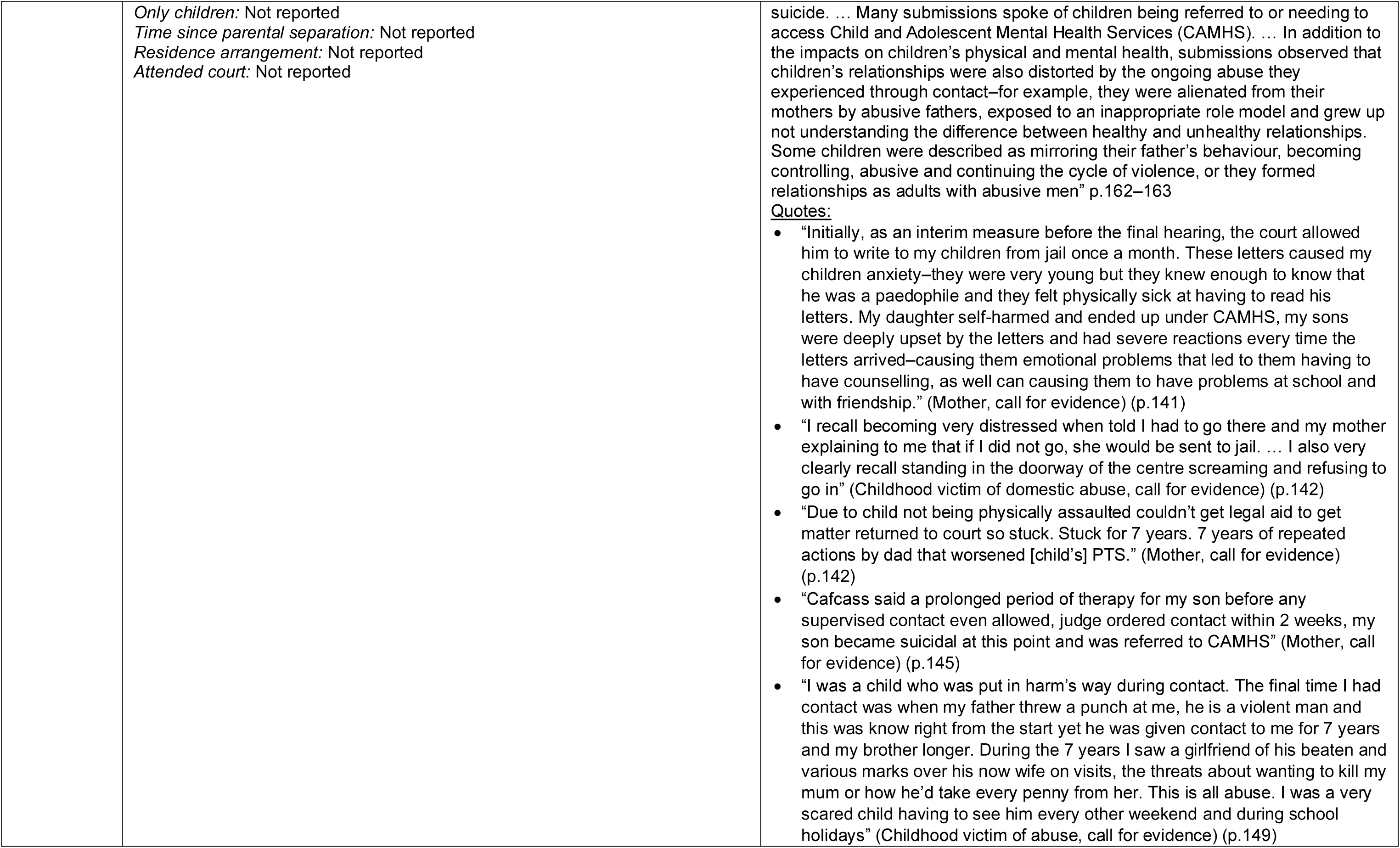
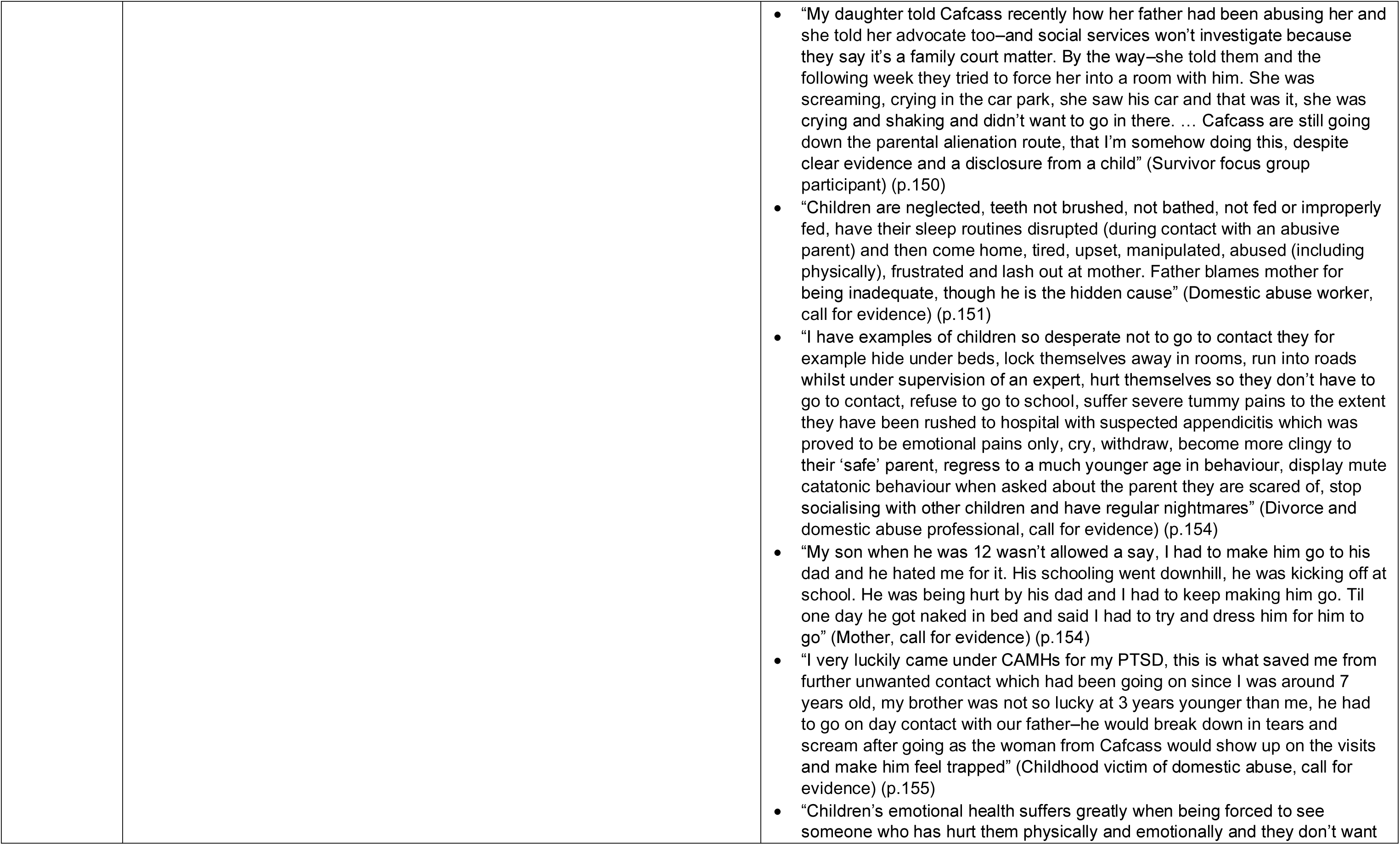
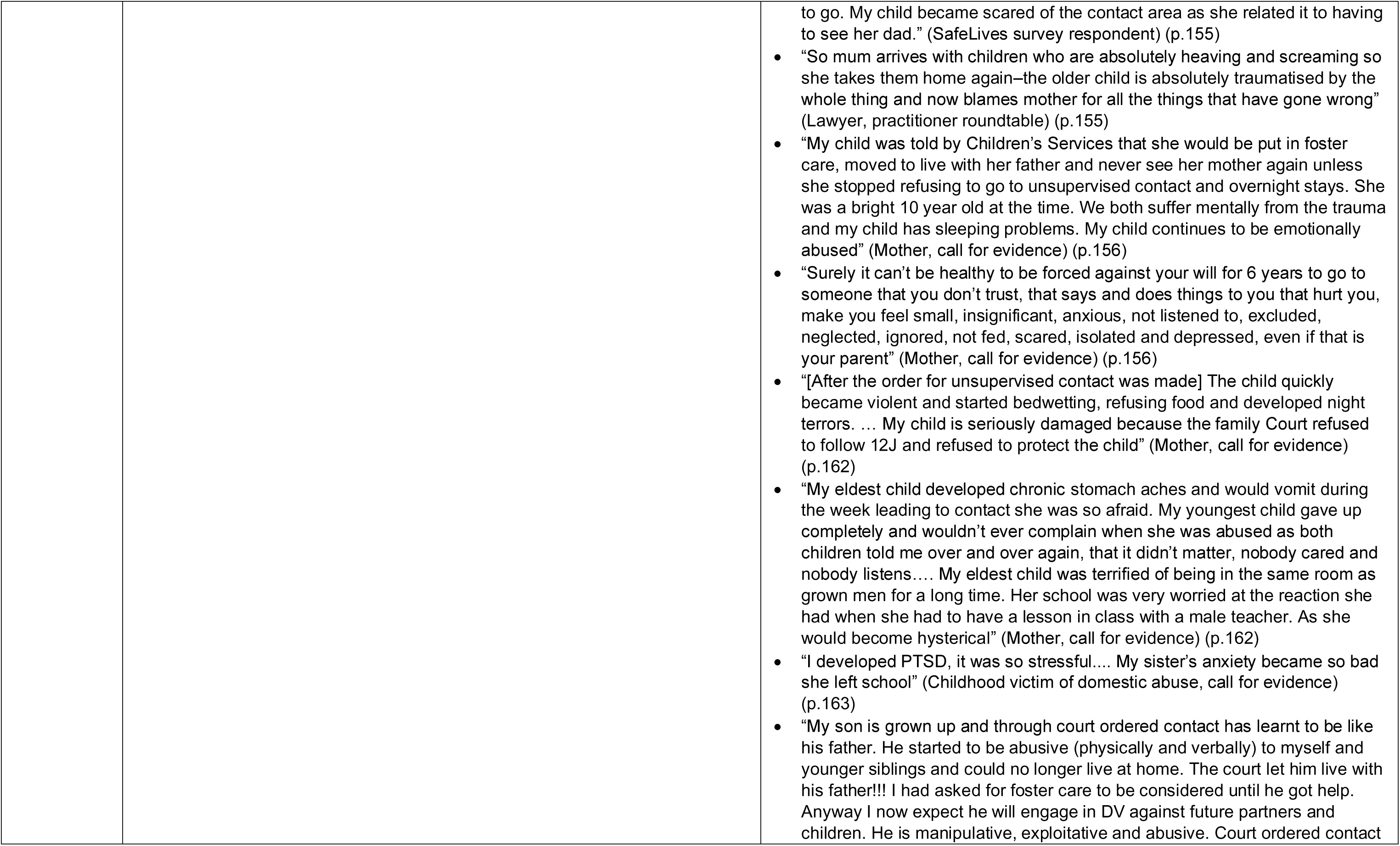
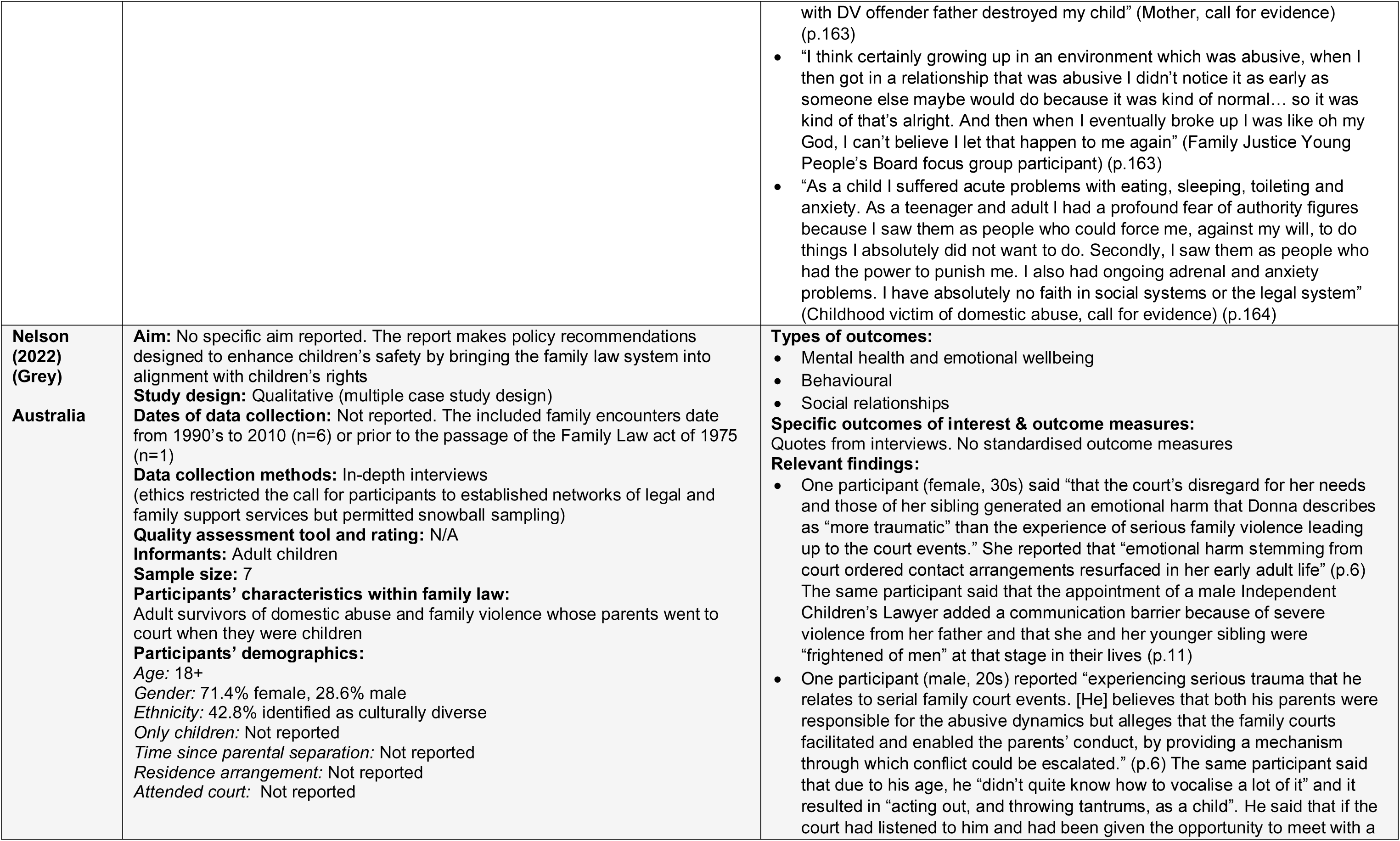
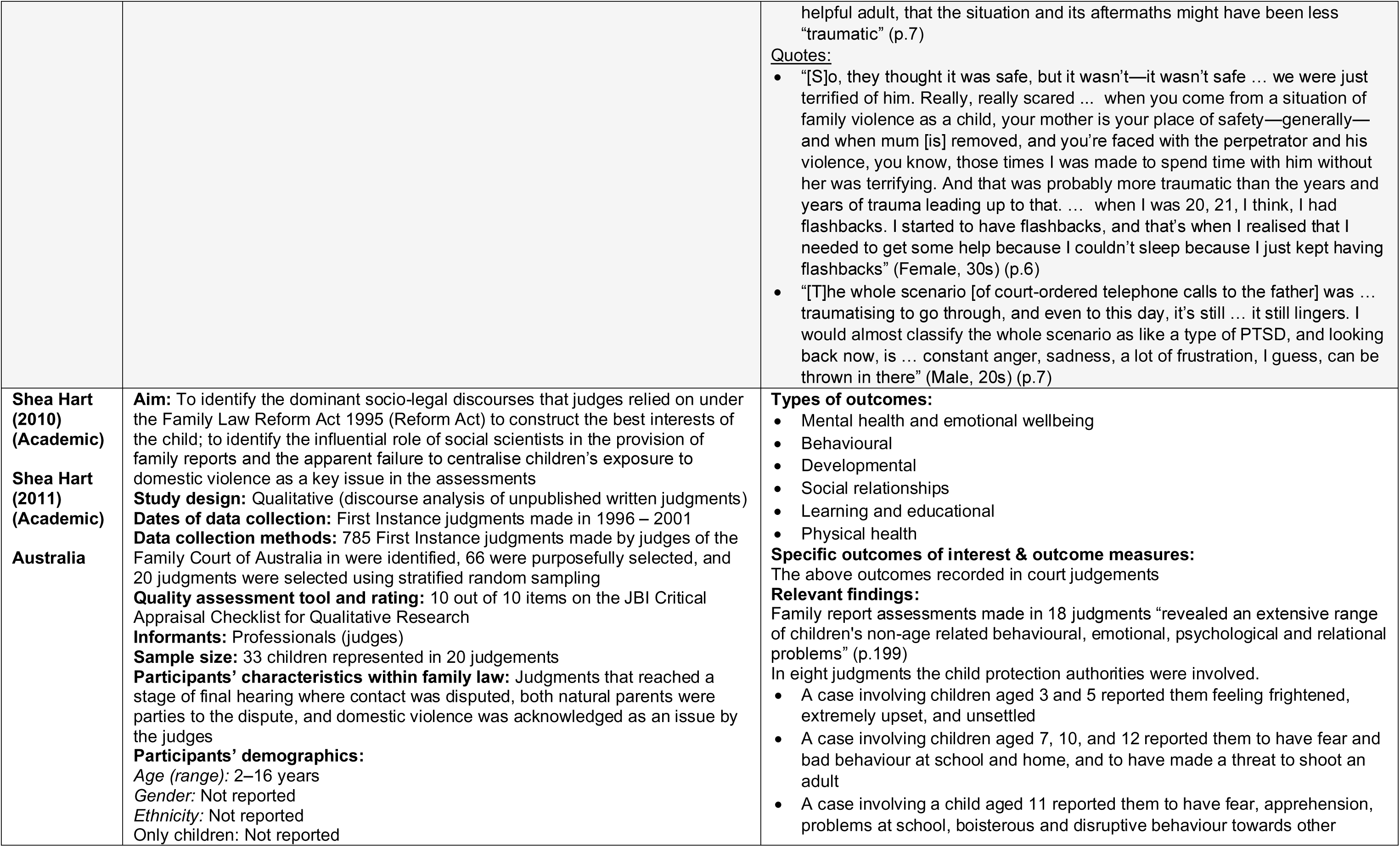
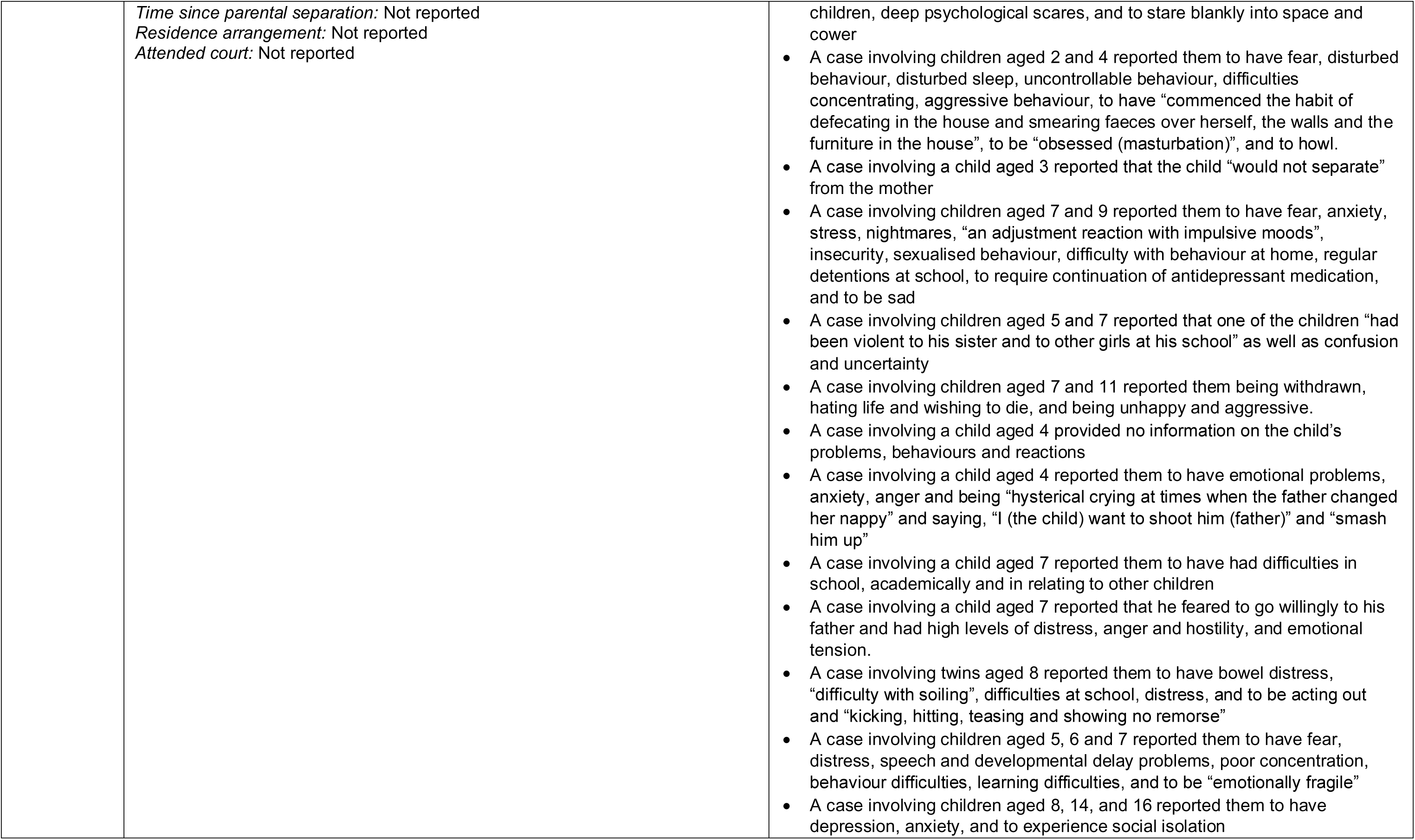
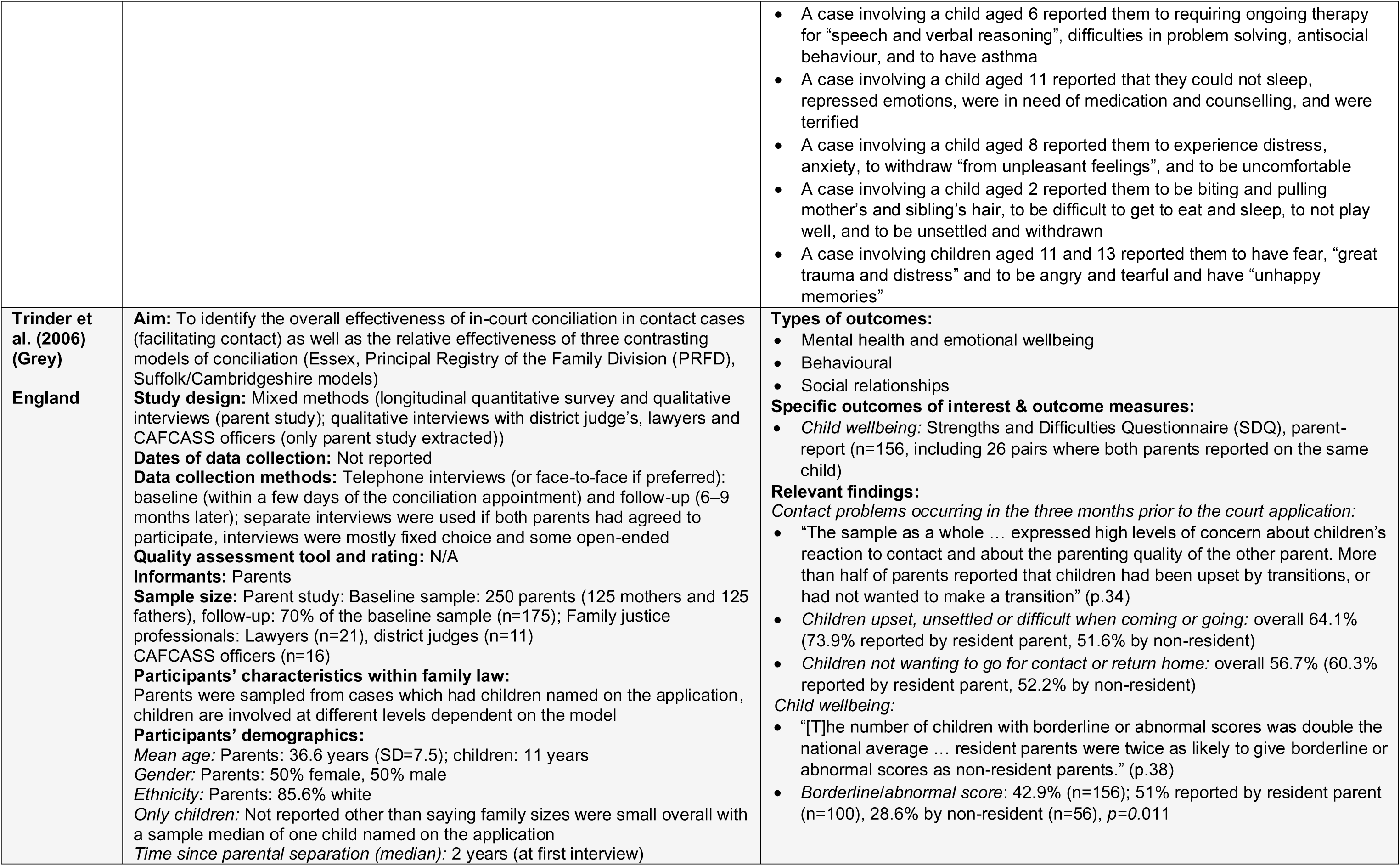
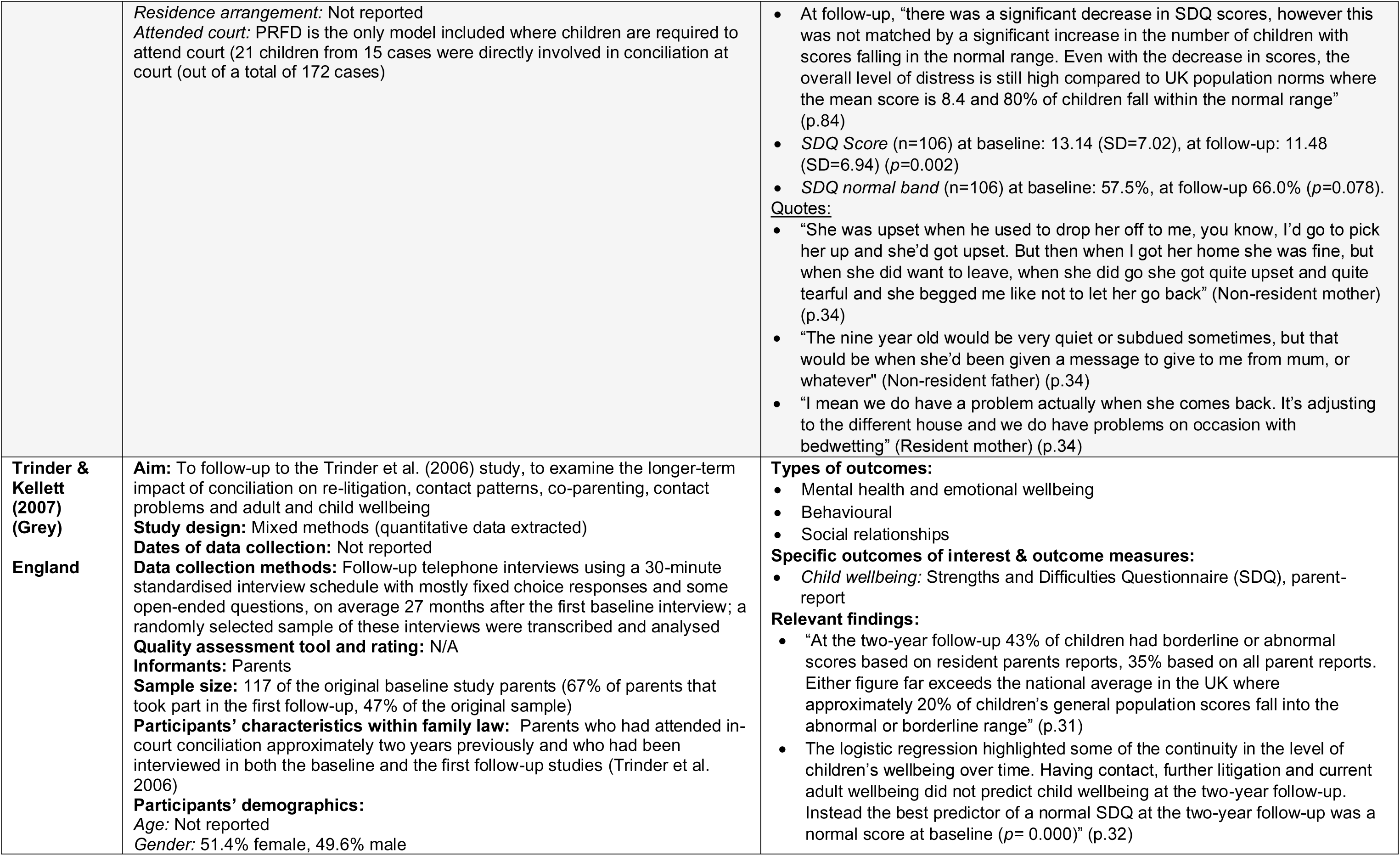
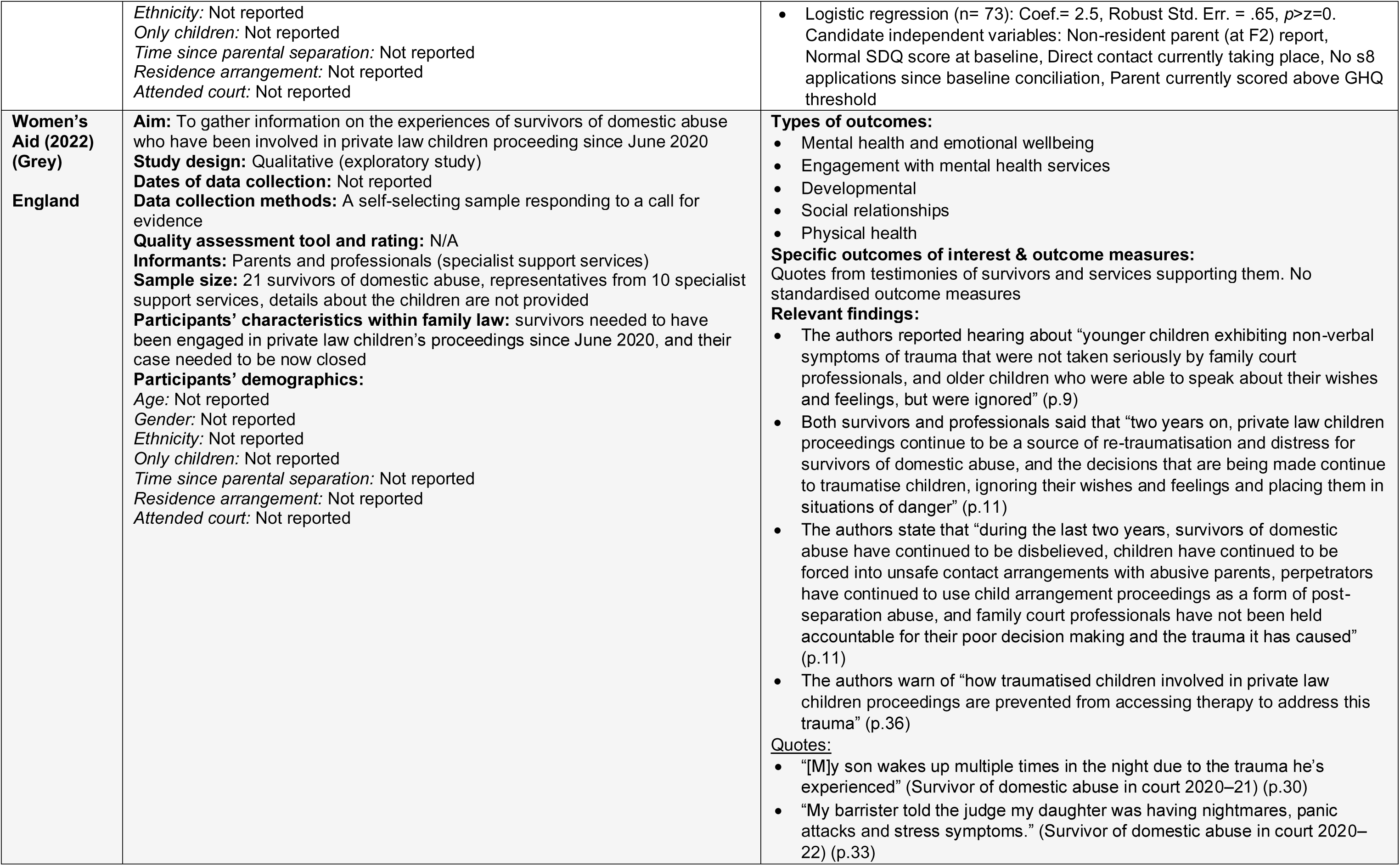
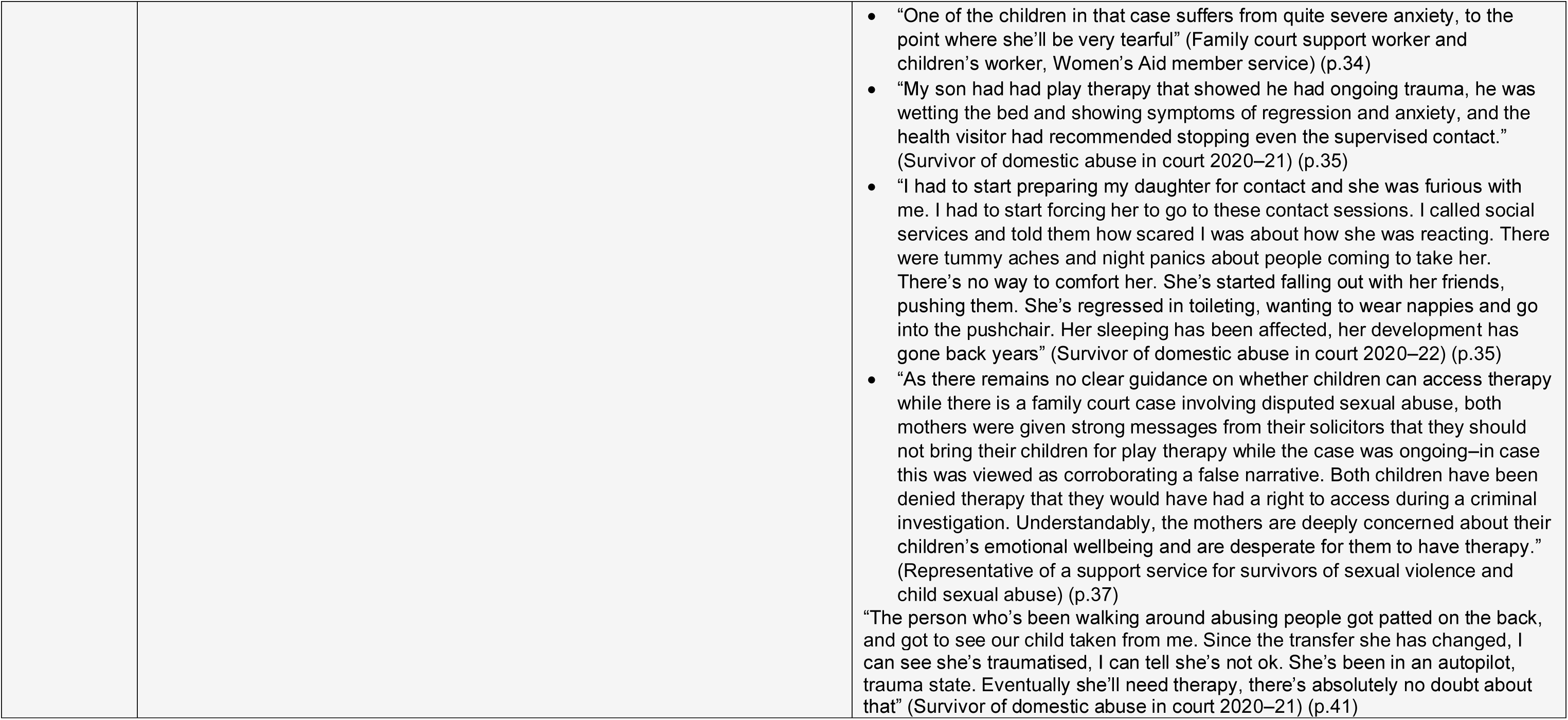

### 5.3 Quality appraisal

#### 5.3.1 Summary of the critical appraisal of the cross-sectional studies

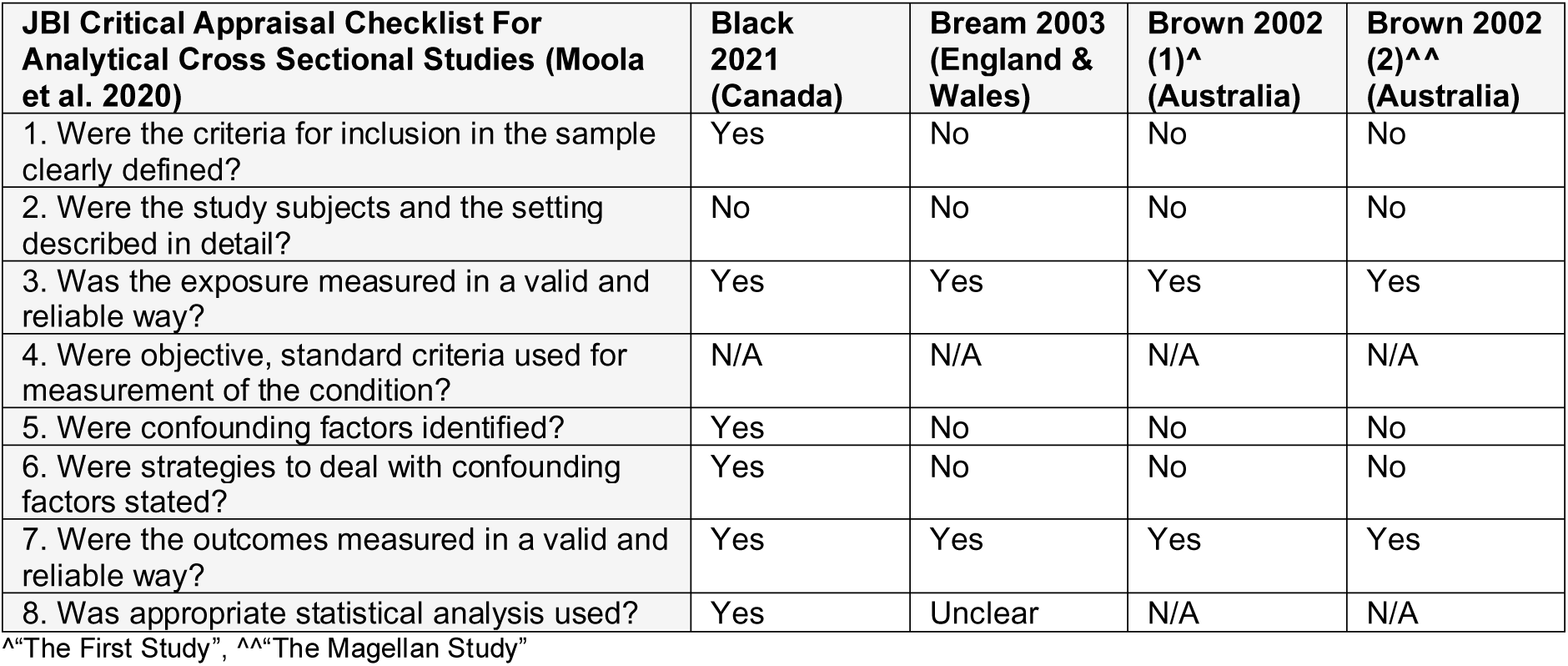

#### 5.3.2 Summary of the critical appraisal of the qualitative studies

6.3.2 Summary of the critical appraisal of the qualitative studies
| **JBICritical Appraisal Checklist For Qualitative Research (Lockwood et al. 2015)** | **Bailey 2011 (England)** | **Darlington 2001 (Australia)** | **Shea Hart 2010/2011 (Australia)** |
| --- | --- | --- | --- |
| 1. Is there congruity between the stated philosophical perspective and the research methodology? | Unclear | Unclear | Yes |
| 2. Is there congruity between the research methodology and the research question or objectives? | Yes | Yes | Yes |
| 3. Is there congruity between the research methodology and the methods used to collect data? | Yes | Yes | Yes |
| 4. Is there congruity between the research methodology and the representation and analysis of data? | Yes | Yes | Yes |
| 5. Is there congruity between the research methodology and the interpretation of results? | Yes | Yes | Yes |
| 6. Is there a statement locating the researcher culturally or theoretically? | No | No | Yes |
| 7. Is the influence of the researcher on the research, and vice-versa, addressed? | No | No | Yes |
| 8. Are participants, and their voices, adequately represented? | No | Yes | Yes |
| 9. Is the research ethical according to current criteria or, for recent studies, and is there evidence of ethical approval by an appropriate body? | Yes | Unclear | Yes |
| 10. Do the conclusions drawn in the research report flow from the analysis, or interpretation, of the data? | Yes | Yes | Yes |

#### 5.3.3 Summary of the critical appraisal of the cohort study

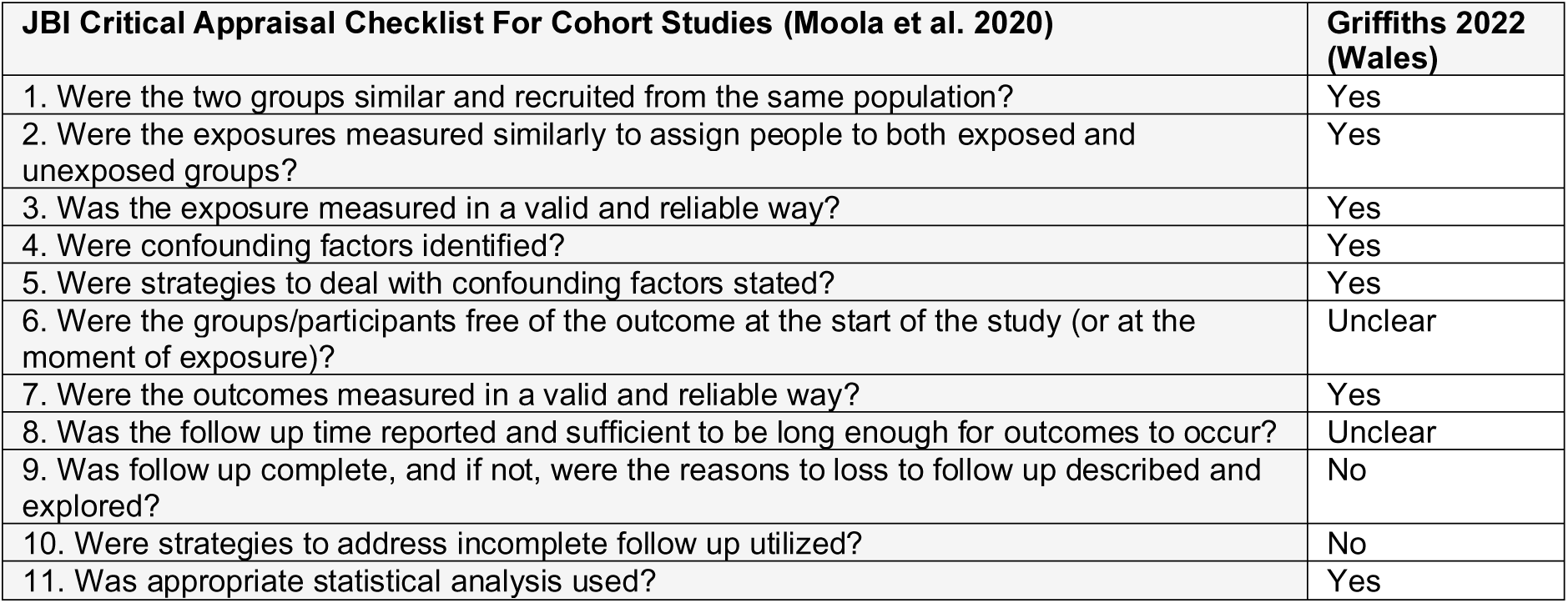

## APPENDIX 1 Database search strategies

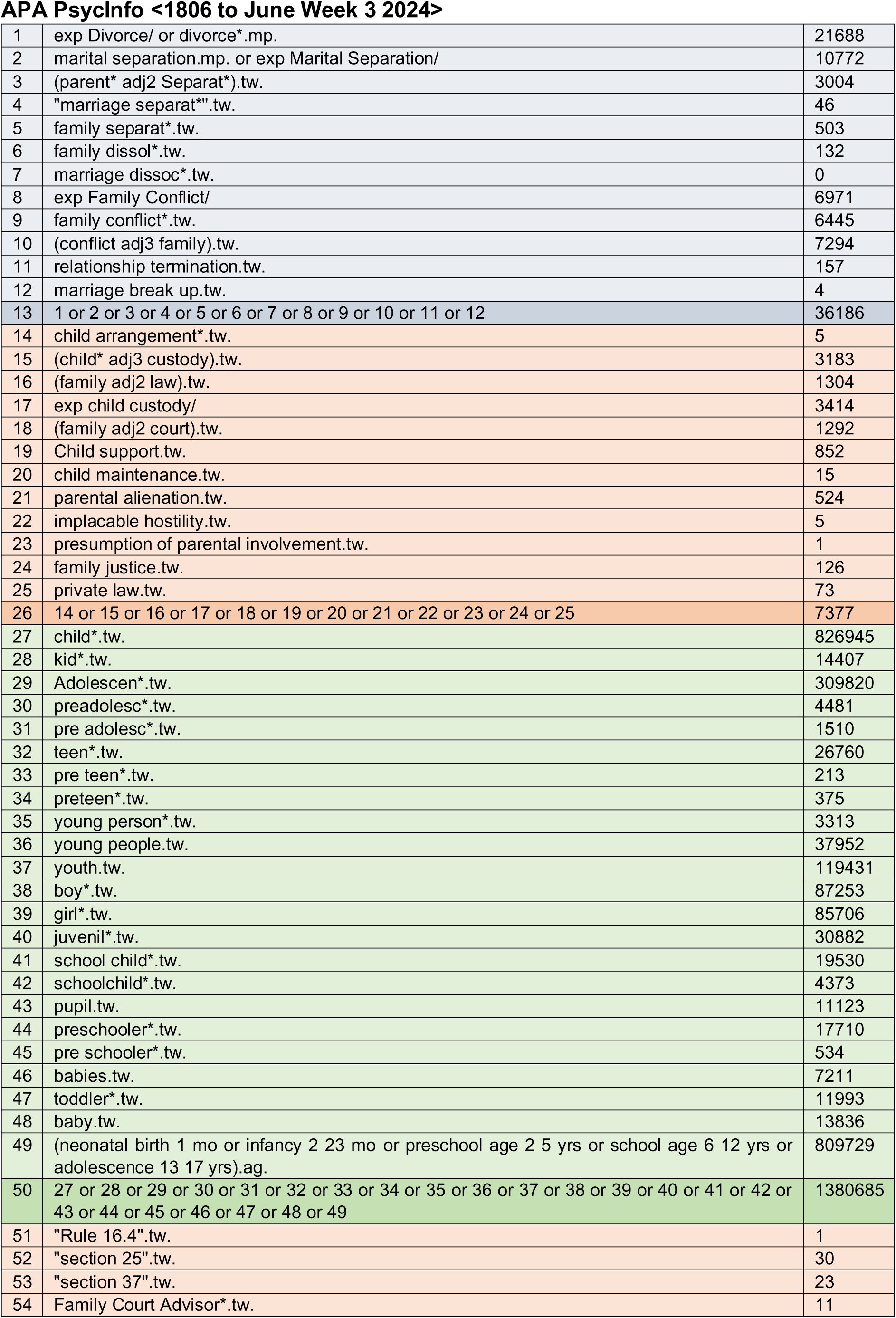
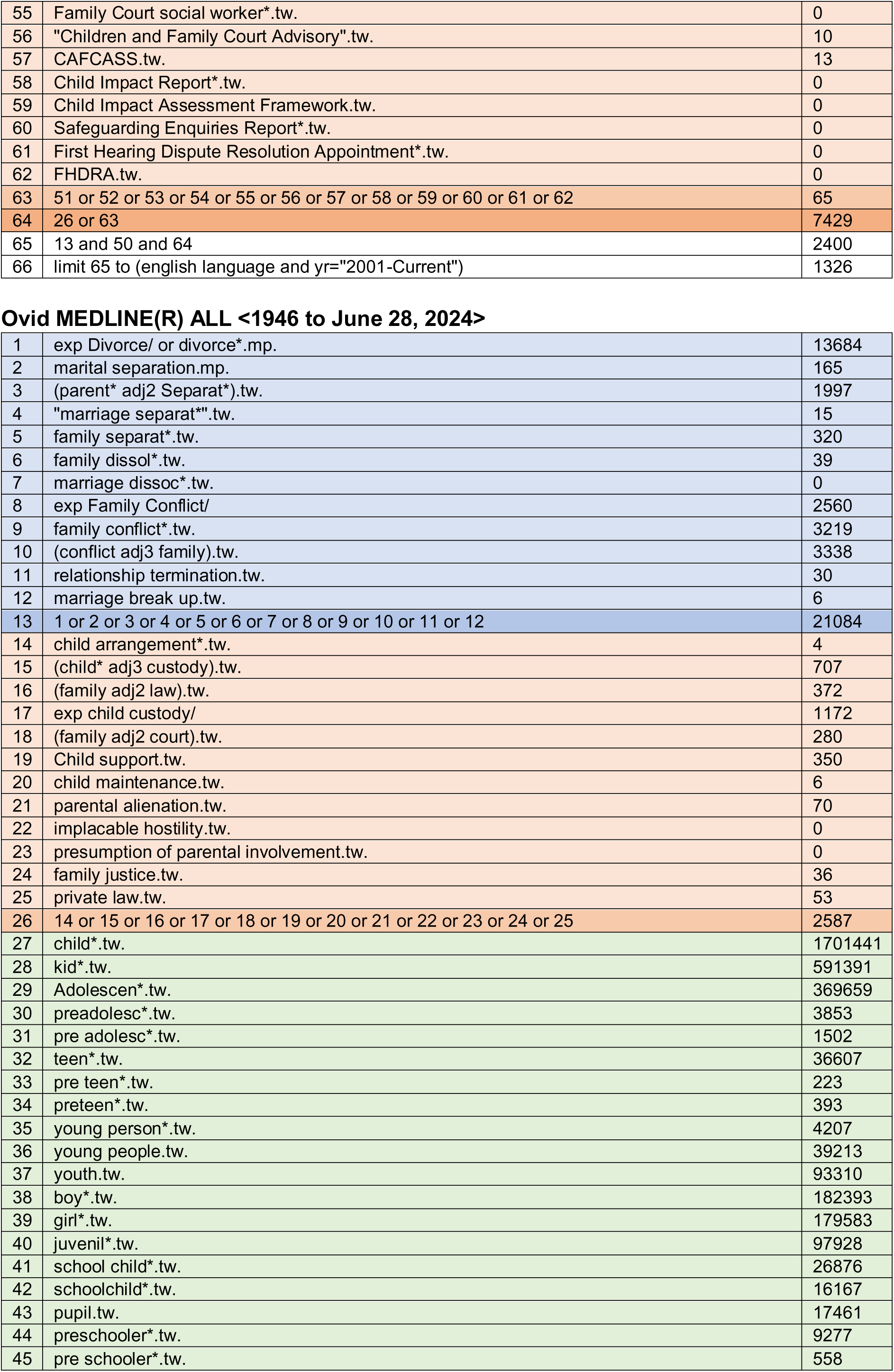
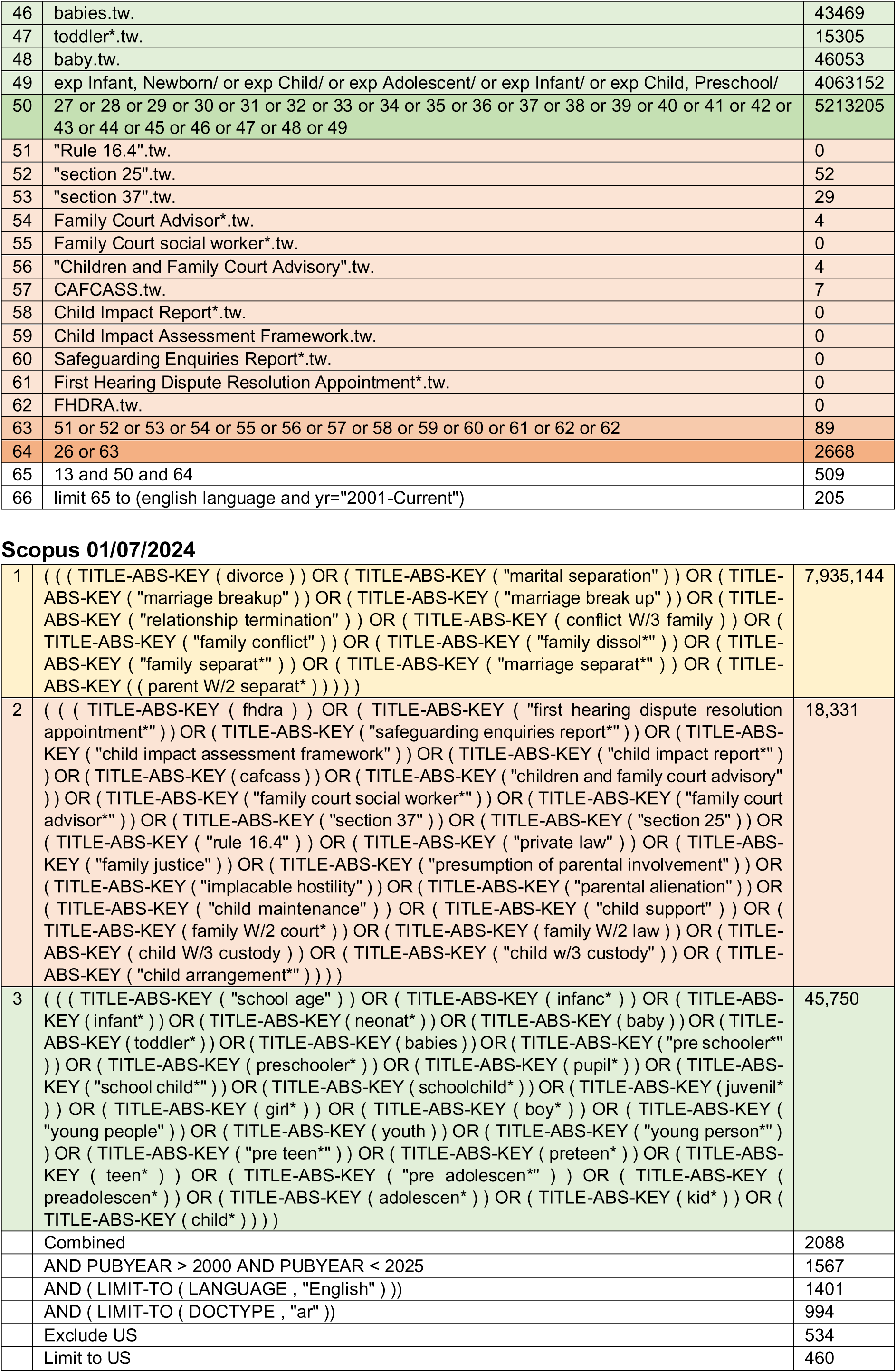
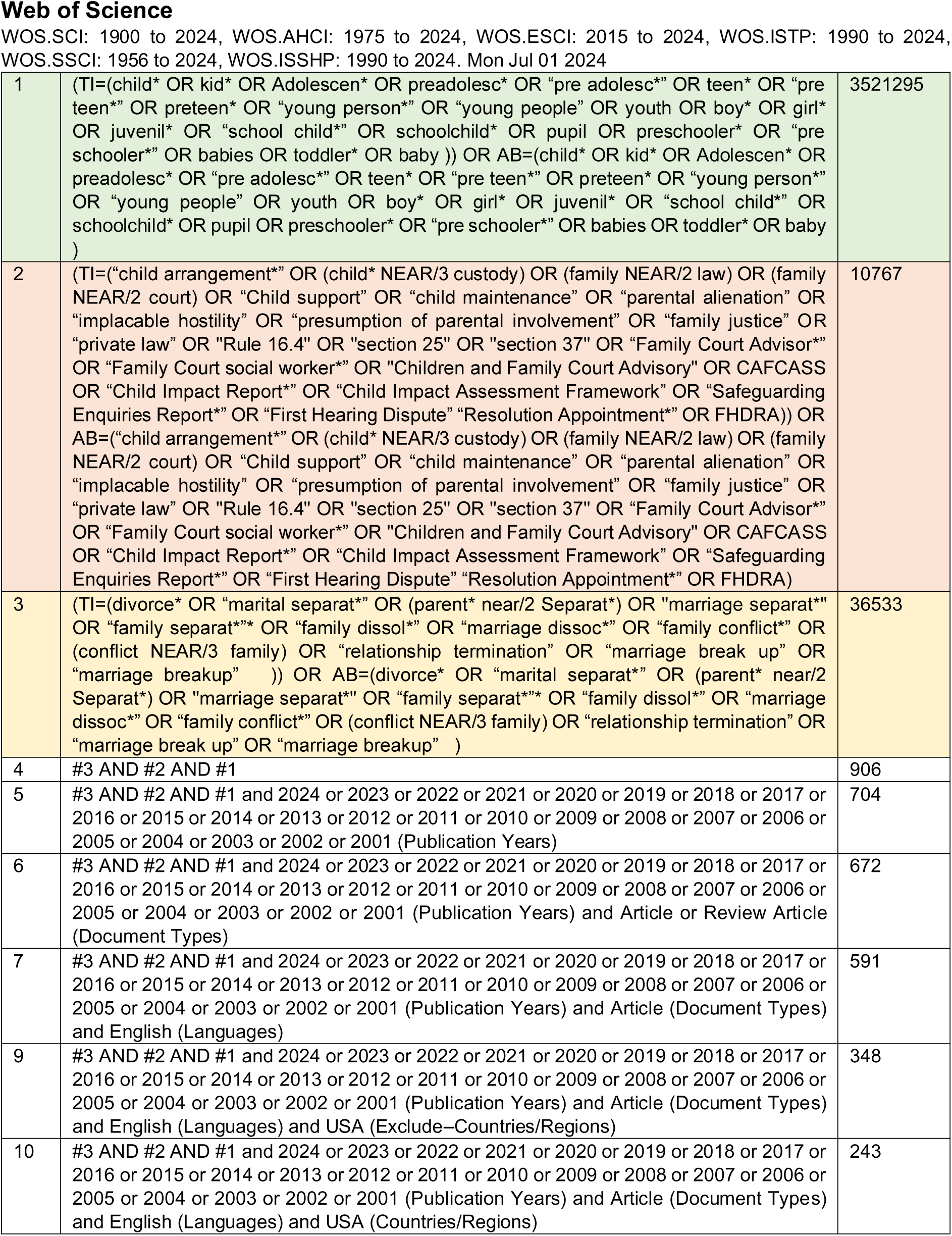

## Social Science Database via Proquest

### Search 1

Applied filters:

- 2001–01–01-2024–07–01
- INCLUDE-United Kingdom--UK OR Australia OR Canada OR England OR Wales title(child* OR kid* OR Adolescen* OR preadolesc* OR (“pre adolescence” OR “pre adolescent” OR “pre adolescents”) OR teen* OR (“pre teen” OR “pre teenage” OR “pre teenager” OR “pre teens”) OR preteen* OR (“young person” OR “young persons”) OR “young people” OR youth OR boy* OR girl* OR juvenil* OR (“school child” OR “school childcare” OR “school children”) OR schoolchild* OR pupil OR preschooler* OR (“pre schooler” OR “pre schoolers”) OR babies OR toddler* OR baby OR child* OR kid* OR Adolescen* OR preadolesc* OR (“pre adolescence” OR “pre adolescent” OR “pre adolescents”) OR teen* OR (“pre teen” OR “pre teenage” OR “pre teenager” OR “pre teens”) OR preteen* OR (“young person” OR “young persons”) OR “young people” OR youth OR boy* OR girl* OR juvenil* OR (“school child” OR “school childcare” OR “school children”) OR schoolchild* OR pupil OR preschooler* OR (“pre schooler” OR “pre schoolers”) OR babies OR toddler* OR baby) AND abstract(divorce OR “marital separation” OR “marriage breakup” OR “marriage break up” OR “relationship termination” OR “family conflict” OR “family dissol*” OR (“family separated” OR “family separation”) OR “marriage separat*” OR “parent separat*”) AND abstract(“first hearing dispute resolution appointment*” OR “safeguarding enquiries report*” OR “child impact assessment framework” OR “child impact report*” OR cafcass OR “children and family court advisory” OR “family court social worker*” OR “family court advisor*” OR “section 37” OR “section 25” OR “rule 16.4” OR “private law” OR “family justice” OR “presumption of parental involvement” OR “implacable hostility” OR “parental alienation” OR “child maintenance” OR “child support” OR “child arrangement*” OR “child custody” OR (“family court” OR “family courts”) OR “family law”). 10 hits

### Search 2

Applied filters:

- 2001-01-01 - 2024-07-01
- INCLUDE Canada OR United Kingdom--UK OR Australia OR England OR Wales
- English

Title(child* OR kid* OR Adolescen* OR preadolesc* OR (“pre adolescence” OR “pre adolescent” OR “pre adolescents”) OR teen* OR (“pre teen” OR “pre teenage” OR “pre teenager” OR “pre teens”) OR preteen* OR (“young person” OR “young persons”) OR “young people” OR youth OR boy* OR girl* OR juvenil* OR (“school child” OR “school childcare” OR “school children”) OR schoolchild* OR pupil OR preschooler* OR (“pre schooler” OR “pre schoolers”) OR babies OR toddler* OR baby OR child* OR kid* OR Adolescen* OR preadolesc* OR (“pre adolescence” OR “pre adolescent” OR “pre adolescents”) OR teen* OR (“pre teen” OR “pre teenage” OR “pre teenager” OR “pre teens”) OR preteen* OR (“young person” OR “young persons”) OR “young people” OR youth OR boy* OR girl* OR juvenil* OR (“school child” OR “school childcare” OR “school children”) OR schoolchild* OR pupil OR preschooler* OR (“pre schooler” OR “pre schoolers”) OR babies OR toddler* OR baby) AND title(divorce OR “marital separation” OR “marriage breakup” OR “marriage break up” OR “relationship termination” OR “family conflict” OR “family dissol*” OR (“family separated” OR “family separation”) OR “marriage separat*” OR “parent separat*”) AND title(“first hearing dispute resolution appointment*” OR “safeguarding enquiries report*” OR “child impact assessment framework” OR “child impact report*” OR cafcass OR “children and family court advisory” OR “family court social worker*” OR “family court advisor*” OR “section 37” OR “section 25” OR “rule 16.4” OR “private law” OR “family justice” OR “presumption of parental involvement” OR “implacable hostility” OR “parental alienation” OR “child maintenance” OR “child support” OR “child arrangement*” OR “child custody” OR (“family court” OR “family courts”) OR “family law”)

OR

Abstract(child* OR kid* OR Adolescen* OR preadolesc* OR (“pre adolescence” OR “pre adolescent” OR “pre adolescents”) OR teen* OR (“pre teen” OR “pre teenage” OR “pre teenager” OR “pre teens”) OR preteen* OR (“young person” OR “young persons”) OR “young people” OR youth OR boy* OR girl* OR juvenil* OR (“school child” OR “school childcare” OR “school children”) OR schoolchild* OR pupil OR preschooler* OR (“pre schooler” OR “pre schoolers”) OR babies OR toddler* OR baby OR child* OR kid* OR Adolescen* OR preadolesc* OR (“pre adolescence” OR “pre adolescent” OR “pre adolescents”) OR teen* OR (“pre teen” OR “pre teenage” OR “pre teenager” OR “pre teens”) OR preteen* OR (“young person” OR “young persons”) OR “young people” OR youth OR boy* OR girl* OR juvenil* OR (“school child” OR “school childcare” OR “school children”) OR schoolchild* OR pupil OR preschooler* OR (“pre schooler” OR “pre schoolers”) OR babies OR toddler* OR baby) AND abstract(divorce OR “marital separation” OR “marriage breakup” OR “marriage break up” OR “relationship termination” OR “family conflict” OR “family dissol*” OR (“family separated” OR “family separation”) OR “marriage separat*” OR “parent separat*”) AND abstract(“first hearing dispute resolution appointment*” OR “safeguarding enquiries report*” OR “child impact assessment framework” OR “child impact report*” OR cafcass OR “children and family court advisory” OR “family court social worker*” OR “family court advisor*” OR “section 37” OR “section 25” OR “rule 16.4” OR “private law” OR “family justice” OR “presumption of parental involvement” OR “implacable hostility” OR “parental alienation” OR “child maintenance” OR “child support” OR “child arrangement*” OR “child custody” OR (“family court” OR “family courts”) OR “family law”) 12 hits

## Sociology Collection via Proquest

### Search 1

Applied filters:

- 2001–01–01-2024–07–01
- INCLUDE-United Kingdom--UK OR Canada OR Australia OR England OR UK OR United Kingdom OR Wales
- English

Title(child* OR kid* OR Adolescen* OR preadolesc* OR (“pre adolescence” OR “pre adolescent” OR “pre adolescents”) OR teen* OR (“pre teen” OR “pre teenage” OR “pre teenager” OR “pre teens”) OR preteen* OR (“young person” OR “young persons”) OR “young people” OR youth OR boy* OR girl* OR juvenil* OR (“school child” OR “school childcare” OR “school children”) OR schoolchild* OR pupil OR preschooler* OR (“pre schooler” OR “pre schoolers”) OR babies OR toddler* OR baby OR child* OR kid* OR Adolescen* OR preadolesc* OR (“pre adolescence” OR “pre adolescent” OR “pre adolescents”) OR teen* OR (“pre teen” OR “pre teenage” OR “pre teenager” OR “pre teens”) OR preteen* OR (“young person” OR “young persons”) OR “young people” OR youth OR boy* OR girl* OR juvenil* OR (“school child” OR “school childcare” OR “school children”) OR schoolchild* OR pupil OR preschooler* OR (“pre schooler” OR “pre schoolers”) OR babies OR toddler* OR baby) AND abstract(divorce OR “marital separation” OR “marriage breakup” OR “marriage break up” OR “relationship termination” OR “family conflict” OR “family dissol*” OR (“family separated” OR “family separation”) OR “marriage separat*” OR “parent separat*”) AND abstract(“first hearing dispute resolution appointment*” OR “safeguarding enquiries report*” OR “child impact assessment framework” OR “child impact report*” OR cafcass OR “children and family court advisory” OR “family court social worker*” OR “family court advisor*” OR “section 37” OR “section 25” OR “rule 16.4” OR “private law” OR “family justice” OR “presumption of parental involvement” OR “implacable hostility” OR “parental alienation” OR “child maintenance” OR “child support” OR “child arrangement*” OR “child custody” OR (“family court” OR “family courts”) OR “family law”). 12 hits

### Search 2

Applied filters:

OR

Abstract(child* OR kid* OR Adolescen* OR preadolesc* OR (“pre adolescence” OR “pre adolescent” OR “pre adolescents”) OR teen* OR (“pre teen” OR “pre teenage” OR “pre teenager” OR “pre teens”) OR preteen* OR (“young person” OR “young persons”) OR “young people” OR youth OR boy* OR girl* OR juvenil* OR (“school child” OR “school childcare” OR “school children”) OR schoolchild* OR pupil OR preschooler* OR (“pre schooler” OR “pre schoolers”) OR babies OR toddler* OR baby OR child* OR kid* OR Adolescen* OR preadolesc* OR (“pre adolescence” OR “pre adolescent” OR “pre adolescents”) OR teen* OR (“pre teen” OR “pre teenage” OR “pre teenager” OR “pre teens”) OR preteen* OR (“young person” OR “young persons”) OR “young people” OR youth OR boy* OR girl* OR juvenil* OR (“school child” OR “school childcare” OR “school children”) OR schoolchild* OR pupil OR preschooler* OR (“pre schooler” OR “pre schoolers”) OR babies OR toddler* OR baby) AND abstract(divorce OR “marital separation” OR “marriage breakup” OR “marriage break up” OR “relationship termination” OR “family conflict” OR “family dissol*” OR (“family separated” OR “family separation”) OR “marriage separat*” OR “parent separat*”) AND abstract(“first hearing dispute resolution appointment*” OR “safeguarding enquiries report*” OR “child impact assessment framework” OR “child impact report*” OR cafcass OR “children and family court advisory” OR “family court social worker*” OR “family court advisor*” OR “section 37” OR “section 25” OR “rule 16.4” OR “private law” OR “family justice” OR “presumption of parental involvement” OR “implacable hostility” OR “parental alienation” OR “child maintenance” OR “child support” OR “child arrangement*” OR “child custody” OR (“family court” OR “family courts”) OR “family law”) 23 hits

## ERIC via Proquest

### Search 1

Applied filters:

- 2001–01–01-2024–07–01
- INCLUDE-Articles, Reports, Reviews
- English

Title(child* OR kid* OR Adolescen* OR preadolesc* OR “pre adolesc*” OR teen* OR “pre teen*” OR preteen* OR “young person*” OR “young people” OR youth OR boy* OR girl* OR juvenil* OR “school child*” OR schoolchild* OR pupil OR preschooler* OR “pre schooler*” OR babies OR toddler* OR baby OR child* OR kid* OR Adolescen* OR preadolesc* OR “pre adolesc*” OR teen* OR “pre teen*” OR preteen* OR “young person*” OR “young people” OR youth OR boy* OR girl* OR juvenil* OR “school child*” OR schoolchild* OR pupil OR preschooler* OR “pre schooler*” OR babies OR toddler* OR baby) AND subject(Divorce OR “marital separation” OR “marriage breakup” OR “marriage break up” OR “relationship termination” OR “family conflict” OR “family dissol*” OR “family separat*” OR “marriage separat*” OR parent separat*) AND abstract(“first hearing dispute resolution appointment*” OR “safeguarding enquiries report*” OR “child impact assessment framework” OR “child impact report*” OR cafcass OR “children and family court advisory” OR “family court social worker*” OR “family court advisor*” OR “section 37” OR “section 25” OR “rule 16.4” OR “private law” OR “family justice” OR “presumption of parental involvement” OR “implacable hostility” OR “parental alienation” OR “child maintenance” OR “child support” OR “child arrangement*” OR “child custody” OR “family court*” OR “family law”). 9 hits

### Search 2

Applied filters:

OR

Abstract(child* OR kid* OR Adolescen* OR preadolesc* OR (“pre adolescence” OR “pre adolescent” OR “pre adolescents”) OR teen* OR (“pre teen” OR “pre teenage” OR “pre teenager” OR “pre teens”) OR preteen* OR (“young person” OR “young persons”) OR “young people” OR youth OR boy* OR girl* OR juvenil* OR (“school child” OR “school childcare” OR “school children”) OR schoolchild* OR pupil OR preschooler* OR (“pre schooler” OR “pre schoolers”) OR babies OR toddler* OR baby OR child* OR kid* OR Adolescen* OR preadolesc* OR (“pre adolescence” OR “pre adolescent” OR “pre adolescents”) OR teen* OR (“pre teen” OR “pre teenage” OR “pre teenager” OR “pre teens”) OR preteen* OR (“young person” OR “young persons”) OR “young people” OR youth OR boy* OR girl* OR juvenil* OR (“school child” OR “school childcare” OR “school children”) OR schoolchild* OR pupil OR preschooler* OR (“pre schooler” OR “pre schoolers”) OR babies OR toddler* OR baby) AND abstract(divorce OR “marital separation” OR “marriage breakup” OR “marriage break up” OR “relationship termination” OR “family conflict” OR “family dissol*” OR (“family separated” OR “family separation”) OR “marriage separat*” OR “parent separat*”) AND abstract(“first hearing dispute resolution appointment*” OR “safeguarding enquiries report*” OR “child impact assessment framework” OR “child impact report*” OR cafcass OR “children and family court advisory” OR “family court social worker*” OR “family court advisor*” OR “section 37” OR “section 25” OR “rule 16.4” OR “private law” OR “family justice” OR “presumption of parental involvement” OR “implacable hostility” OR “parental alienation” OR “child maintenance” OR “child support” OR “child arrangement*” OR “child custody” OR (“family court” OR “family courts”) OR “family law”) 19 hits

## APPENDIX 2: Searched websites

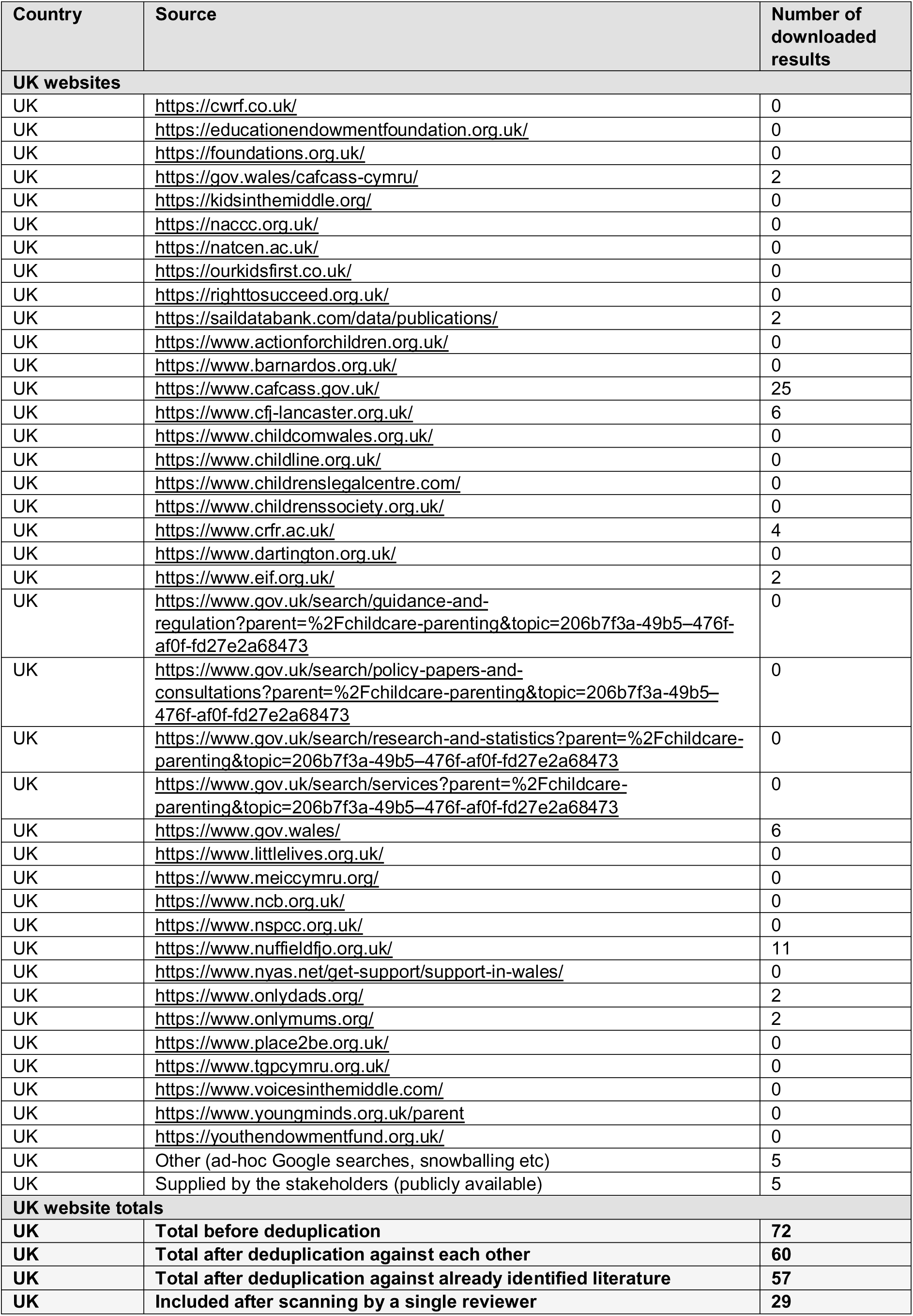
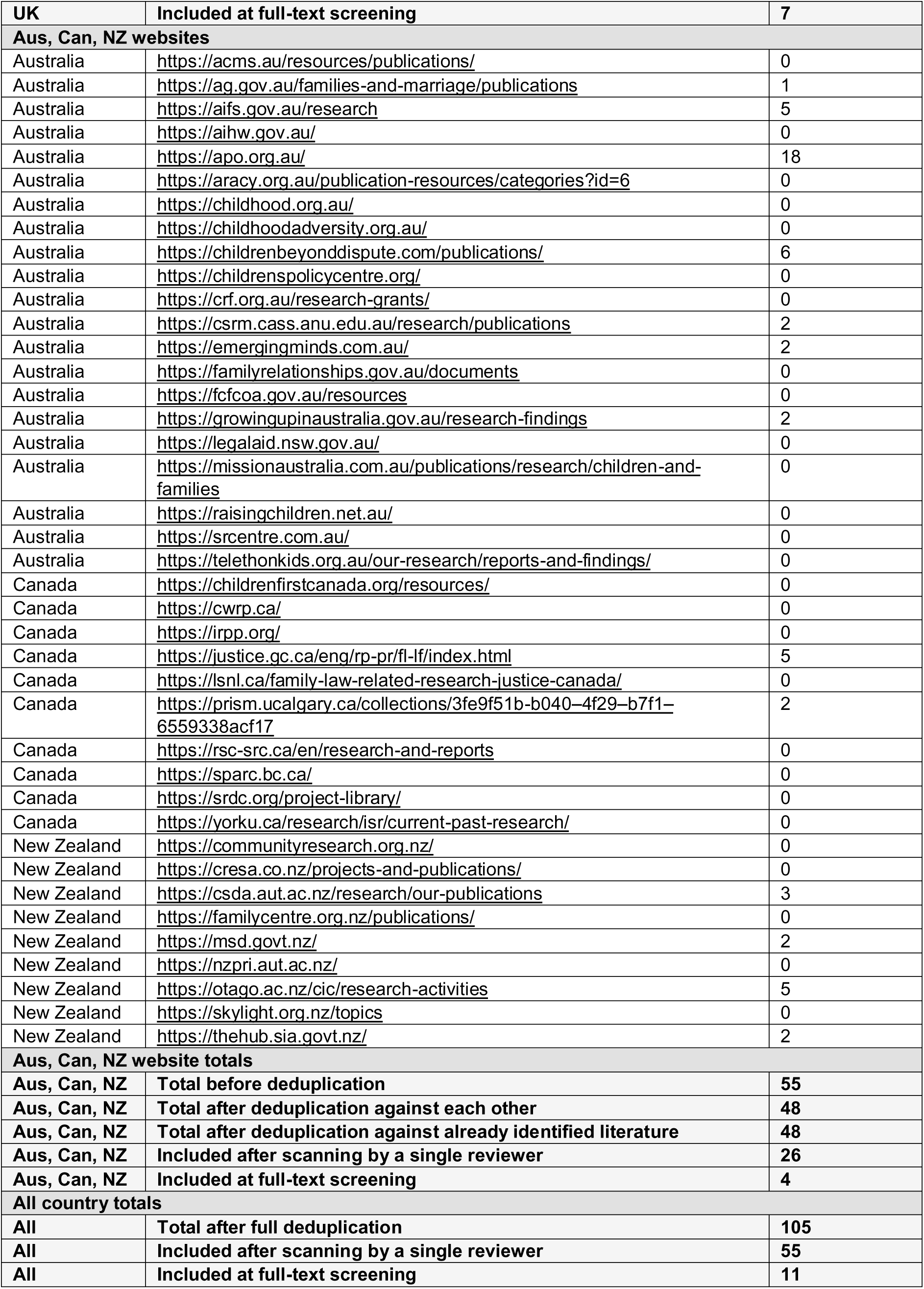

## APPENDIX 3: Studies excluded at full-text screening

| Full citation | Type | Source | Reason for exclusion |
| --- | --- | --- | --- |
| Afifi TD, McManus T, Hutchinson S, et al. (2007). Inappropriate parental divorce disclosures, the factors that prompt them, and their impact on parents' and adolescents' well-being. *Communication Monographs*. 74(1): 78-102. doi:https://dx.doi.org/10.1080/03637750701196870 | Academic | Database searches | Wrong country |
| Allen H. (2014). Unpacking court divorce decrees, children's outcomes, and three unconfounded determinants: An evidence-based look. *Journal of Divorce & Remarriage*. 55(3): 179-205. doi:https://dx.doi.org/10.1080/10502556.2014.887376 | Academic | Database searches | Wrong study design |
| Bagshaw D. (2007). Reshaping Responses to Children When Parents are Separating: Hearing Children's Voices in the Transition. *Australian Social Work*. 60(4): 450-65. doi: 10.1080/03124070701671164 | Academic | Review unpicking | No court involvement or insufficient information about court involvement |
| Baker AJL, Eichler A. (2016). The linkage between parental alienation behaviors and child alienation. *Journal of Divorce & Remarriage*. 57(7): 475-84. | Academic | Review unpicking | Wrong country |
| Bala N, Bertrans L, Wheeler A, et al. (2012). A Study of Post-Separation/Divorce Parental Relocation. https://www.justice.gc.ca/eng/rp-pr/fl-lf/divorce/spsdpr-edpads/index.html | Grey | Website searches | No court involvement or insufficient information about court involvement |
| Barlow A, Ewing J, Hunter R, et al. (2017). Creating paths to family justice: Briefing Paper & Report on Key Findings, University of Exeter. [https://www.exeter.ac.uk/v8media/universityofexeter/collegeofsocialsciencesandinternationalstudies/lawimages/familyregulationandsociety/Creating_Paths_briefing_paper_final_for_website_02.10.17_isbn_(003)_05-03-18.pdf](https://www.exeter.ac.uk/v8media/universityofexeter/collegeofsocialsciencesandinternationalstudies/lawimages/familyregulationandsociety/Creating_Paths_briefing_paper_final_for_website_02.10.17_isbn_%28003%29_05-03-18.pdf) | Grey | Website searches | Wrong outcome |
| Barnett A. (2015). 'Like Gold Dust These Days': Domestic Violence Fact-Finding Hearings in Child Contact Cases. *FEMINIST LEGAL STUDIES*. 23(1): 47-78. doi: 10.1007/s10691-015-9278-4 | Academic | Review unpicking | Wrong outcome |
| Barnett A. (2017). 'Greater than the mere sum of its parts': coercive control and the question of proof. | Grey | Review unpicking | Wrong outcome |
| Barnett A. (2020). A genealogy of hostility: parental alienation in England and Wales. *JOURNAL OF SOCIAL WELFARE AND FAMILY LAW*. 42(1): 18-29. doi: 10.1080/09649069.2019.1701921 | Academic | Review unpicking | Wrong outcome |
| Birchall J, Choudhry S. (2018). What about my right not to be abused? Domestic abuse, human rights and the family courts. *Women's Aid*. | Grey | Review unpicking | Wrong outcome |
| Birnbaum R, Saini M. (2013). A scoping review of qualitative studies about children experiencing parental separation. *Childhood: A Global Journal of Child Research*. 20(2): 260-82. doi:https://dx.doi.org/10.1177/0907568212454148 | Academic | Database searches | Wrong outcome |
| Birnbaum R. (2017). Views of the child reports: Hearing directly from children involved in post-separation disputes. *Social Inclusion*. 5(3): 148-54. doi:10.17645/si.v5i3.922 | Academic | Database searches | Wrong study design |
| Bow JN, Gould JW, Flens JR. (2009). Examining parental alienation in child custody cases: A survey of mental health and legal professionals. *The American Journal of Family Therapy*. 37(2): 127-45. | Academic | Review unpicking | Wrong country |
| Bush JE, Ehrenberg MF. (2003). Young persons' perspectives on the influence of family transitions on sibling relationships: A qualitative exploration. *Journal of Divorce & Remarriage*. 39(3): 1-35. doi:https://dx.doi.org/10.1300/J087v39n03_01 | Academic | Database searches | No court involvement or insufficient information about court involvement |
| Butler I, Scanlan L, Robinson M, et al. (2002). Children's involvement in their parents' divorce: implications for practice. *Children & Society*. 16(2): 89-102. doi: https://doi.org/10.1002/chi.702 | Academic | Review unpicking | No court involvement or insufficient information about court involvement |
| CAFCASS and Woman's Aid. (2017). Allegations of domestic abuse in child contact cases. https://www.cafcass.gov.uk/sites/default/files/migrated/Allegations-of-domestic-abuse-in-child-contact-cases-2017.pdf | Grey | Website searches | Data not extractable |
| CAFCASS Cymru. (2019a). Children's Resistance or Refusal To Spend Time With A Parent: Practice Guidance. https://www.gov.wales/sites/default/files/publications/2019-08/cafcass-cymru-childrens-resistance-or-refusal-to-spending-time-with-a-parent-practice-guidance.pdf | Grey | Website searches | Wrong publication type |
| CAFCASS Cymru. (2019b). Impact On Children of Experiencing Domestic Abuse. https://www.gov.wales/sites/default/files/publications/2019-08/cafcass-cymru-impact-on%20children-experiencing-domestic-abuse.pdf | Grey | Website searches | No court involvement or insufficient information about court involvement |
| CAFCASS. (2014). Report of an analysis of rule 16.4 appointments considered by the courts in September 2014. https://www.cafcass.gov.uk/sites/default/files/migrated/Report-of-an-analysis-of-rule-16.4-appointments-considered-by-the-courts-in-September-2014.pdf | Grey | Website searches | Wrong outcome |
| CAFCASS. (2016a). Study into Cafcass' role at First Hearing Dispute Resolution Appointments. https://www.cafcass.gov.uk/sites/default/files/migrated/Study-into-Cafcass%E2%80%99-role-at-First-Hearing-Dispute-Resolution-Appointments-2016.pdf | Grey | Website searches | Wrong outcome |
| CAFCASS. (2016b). Study of data held by Cafcass in cases featuring radicalisation concerns. https://www.cafcass.gov.uk/sites/default/files/migrated/Study-of-data-held-by-Cafcass-in-cases-featuring-radicalisation-concerns-2016.pdf | Grey | Website searches | Wrong outcome |
| CAFCASS. (2016c). What happened next? A study of interim outcomes following private law proceedings as reported by parents six to nine months on. https://www.cafcass.gov.uk/sites/default/files/migrated/What-happened-next-A-study-of-interim-outcomes-following-private-law-proceedings-2016.pdf | Grey | Website searches | Wrong outcome |
| CAFCASS. (2017). Cases featuring trafficking as a 'child need'. https://www.cafcass.gov.uk/sites/default/files/migrated/Study-of-data-held-by-Cafcass-in-cases-featuring-trafficking-as-a-child-need-2017.pdf | Grey | Website searches | Wrong outcome |
| Caffrey L. (2013). Hearing the Voice of the Child: The Role of Child Contact Centres in the Family Justice System. *Child & Fam. LQ*. 25: 357. | Academic | Review unpicking | Wrong outcome |
| Campbell A. (2008). The right to be heard: Australian children's views about their involvement in decision-making following parental separation. *Child Care in Practice*. 14(3): 237-55. doi: 10.1080/13575270802042496 | Academic | Review unpicking | No court involvement or insufficient information about court involvement |
| Carlson MJ. (2006). Family structure, father involvement, and adolescent behavioral outcomes. *Journal of marriage and family*. 68(1): 137-54. doi: 10.1111/j.1741-3737.2006.00239.x | Academic | Review unpicking | Wrong country |
| Carson R, Dunstan E, Dunstan J, et al. (2018). Children and young people in separated families: Family law system experiences and needs. Melbourne: Australian Institute of Family Studies. 1-111. | Grey | Review unpicking | Wrong outcome |
| Cashmore J, Parkinson P. (2009). What responsibility do courts have to hear children's voices? *Children's Rights*. Routledge. | Grey | Review unpicking | Wrong outcome |
| ChildLine. (2010). Casenote Family Relationship Problems. https://www.slideshare.net/slideshow/casenote-family-relationship-problems/5450558 | Grey | Website searches | No court involvement or insufficient information about court involvement |
| Coley RL, Medeiros BL. (2007). Reciprocal longitudinal relations between nonresident father involvement and adolescent delinquency. *Child development*. 78(1): 132-47. doi: DOI 10.1111/j.1467-8624.2007.00989.x | Academic | Review unpicking | No court involvement or insufficient information about court involvement |
| Connolly H. (2018). What children and young people think should happen when families separate. Commissioner for Children and Young People, South Australia. https://apo.org.au/sites/default/files/resource-files/2018-08/apo-nid323628.pdf | Grey | Website searches | Wrong population |
| Cusworth L, Hargreaves C, Alrouh B, et al. (2021). Uncovering private family law: Adult characteristics and vulnerabilities (Wales), London, Nuffield Family Justice Observatory. https://popdatasci.swan.ac.uk/wp-content/uploads/2021/11/Eng.-report.-nfjo_private-law_adults_final_20211110.pdf | Grey | Website searches | Wrong population |
| Cusworth L, Hargreaves C, Alrouh B, et al. (2022). Adults in private family law proceedings in Wales: characteristics and vulnerabilities. *International Journal of Population Data Science*. 7(3). doi: 10.23889/ijpds.v7i3.1910 | Academic | Website searches | Wrong population |
| Cyr F, Di Stefano G, Desjardins B. (2013). Family life, parental separation, and child custody in Canada: A focus on Quebec. *Special Issue: Global family law*. 51(4): 522-41. doi: doi:https://dx.doi.org/10.1111/fcre.12050 | Academic | Database searches | No court involvement or insufficient information about court involvement |
| Dare H, Goodman C, John R, et al. (2019). Toward better outcomes for families through innovative dispute resolution programs in England. *Family Court Review*. 57(3): 368-74. doi: doi:https://dx.doi.org/10.1111/fcre.12421 | Academic | Database searches | Wrong study design |
| Death J, Ferguson C, Burgess K. (2019). Parental alienation, coaching and the best interests of the child: Allegations of child sexual abuse in the Family Court of Australia. *Child Abuse & Neglect*. 94. doi: 10.1016/j.chiabu.2019.104045 | Academic | Citation searching | Wrong outcome |
| Dreman S, Shemi R. (2004). Perception of family structure, state-anger, and parent-child communication and adjustment of children of divorced parents. *Journal of Divorce & Remarriage*. 41(1): 47-68. doi: doi:https://dx.doi.org/10.1300/J087v41n01_04 | Academic | Database searches | Wrong country |
| Dunn J, Cheng H, O'Connor TG, et al. (2004). Children's perspectives on their relationships with their nonresident fathers: influences, outcomes and implications. *Journal of child psychology and psychiatry*. 45(3): 553-66. doi: DOI 10.1111/j.1469-7610.2004.00245.x | Academic | Review unpicking | No court involvement or insufficient information about court involvement |
| Fabricius WV, Luecken LJ. (2007). Postdivorce living arrangements, parent conflict, and long-term physical health correlates for children of divorce. *Journal of Family Psychology*. 21(2): 195-205. doi: doi:https://dx.doi.org/10.1037/0893-3200.21.2.195 | Academic | Database searches | Wrong country |
| Family Law Council. (2016). Report on Families with Complex Needs and the Intersection of Family Law and Child Protection, Commonwealth of Australia. https://www.ag.gov.au/sites/default/files/2020-03/Family-with-Complex-Needs-Intersection-of-Family-Law-and-Child-Protection-Systems-Final-Report-Terms-3-4-5.PDF | Grey | Website searches | Wrong publication type |
| Finley GE, Schwartz SJ. (2007). Father involvement and long-term young adult outcomes: The differential contributions of divorce and gender. *Family Court Review*. 45(4): 573-87. doi: doi:https://dx.doi.org/10.1111/j.1744-1617.2007.00172.x | Academic | Database searches | Wrong country |
| Finley GE, Schwartz SJ. (2010). The divided world of the child: Divorce and long-term psychosocial adjustment. *Family Court Review*. 48(3): 516-27. doi: doi:https://dx.doi.org/10.1111/j.1744-1617.2010.01326.x | Academic | Database searches | Wrong country |
| Fortin J, Hunt J, Scanlan L. (2012). Taking a longer view of contact: the perspectives of young adults who experience parental separation in their youth. | Grey | Review unpicking | Data not extractable |
| Fowler E, Stewart S. (2005). A review of cases involving 95 children in which the National Youth Advocacy Service represented children in family proceedings pursuant to rule 9.5 of the Family Proceedings Rules 1991. *Family Law*. January(35): 49-52. | Academic | Citation searching | Wrong outcome |
| Francia L, Millea P. (2015). Mastery or misery: Conflict between separated parents a psychological burden for children. *Journal of Divorce & Remarriage*. 56(7): 551-68. doi: https://dx.doi.org/10.1080/10502556.2015.1080090 | Academic | Database searches | Data not extractable |
| Friesen MD, Horwood LJ, Fergusson DM, Woodward LJ. (2017) Exposure to parental separation in childhood and later parenting quality as an adult: evidence from a 30-year longitudinal study. *Journal of Child Psychology and Psychiatry* 58(1), 30–37. doi: 10.1111/jcpp.12610 | Academic | Website searches | No court involvement or insufficient information about court involvement |
| Giancarlo C, Rottmann K. (2014). Kids come last: The effect of family law involvement in parental alienation. *International Journal of Interdisciplinary Social Sciences*. 9(1): 27-42. doi: doi:10.18848/1833-1882/CGP/v09/53552 | Academic | Database searches | Wrong outcome |
| Gollop M, Taylor N, Cameron C, et al. (2019). Parenting Arrangements after Separation Study: Evaluating the 2014 Family Law Reforms – Parents’ and caregivers’ perspectives – Part 1 Research Report for the New Zealand Law Foundation., Dunedin, New Zealand, Children’s Issues Centre, University of Otago. https://www.otago.ac.nz/_data/assets/pdf_file/0022/328090/read-the-phase-two-parents-and-caregivers-perspectives-part-1-report-739545.pdf | Grey | Website searches | Wrong population |
| Graham A, Fitzgerald R. (2006). Taking account of the ‘to and fro’ of children’s experiences in family law. *Children Australia*. 31(2): 30-6. doi: 10.1017/S103507720001110X | Academic | Review unpicking | Wrong outcome |
| Grey R. (2023). “Catastrophic”: A qualitative exploration of survivors experiences of expert instruction in private law child arrangements proceedings. *Journal of Social Welfare and Family Law*. 45(4): 344-62. doi: 10.1080/09649069.2023.2281832 | Academic | Website searches | Wrong population |
| Hale B. (2010). The children act 1989 in the highest courts. *Journal of Children's Services*. 5(2): 17-28. doi: 10.5042/jcs.2010.0298 | Academic | Citation searching | Wrong case type |
| Halliday E, Green R, Marsh B. (2017). Private law cases that return to court: a Cafcass study. https://www.cafcass.gov.uk/sites/default/files/migrated/private_law_cases_that_return_to_court_-_cafcass_research_november_2017.pdf | Grey | Website searches | Wrong outcome |
| Harding M. (2015). How do county courts share the care of children between parents? https://www.nuffieldfoundation.org/wp-content/uploads/2019/11/Full20report.pdf | Grey | Website searches | Wrong outcome |
| Hargreaves C, Cusworth L, Alrouh B, et al. (2022). Uncovering private family law: What can the data tell us about children’s participation?, Nuffield Family Justice Observatory. https://www.nuffieldfjo.org.uk/resource/uncovering-private-family-law-what-can-the-data-tell-us-about-childrens-participation | Grey | Website searches | Wrong outcome |
| Hargreaves C, Cusworth L, Alrouh B, et al. (2024). Uncovering private family law: how often do we hear the voice of the child?, Nuffield Family Justice Observatory. https://www.nuffieldfjo.org.uk/resource/uncovering-private-family-law-how-often-do-we-hear-the-voice-of-the-child | Grey | Website searches | Wrong outcome |
| Higgins D, J. (2007). Cooperation and Coordination: An evaluation of the Family Court of Australia’s Magellan case-management model, Canberra, Australia, Family Court of Australia. https://apo.org.au/sites/default/files/resource-files/2007-10/apo-nid8337.pdf | Grey | Website searches | Wrong outcome |
| Hight G, Jamieson L. (2007). Cool with change: Young people and family change. | Grey | Review unpicking | Wrong outcome |
| Hughes R, Rose W. (2010). Coming of age: has the Children Act 1989 lived up to its promise? *Journal of Children's Services*. 5(2): 2-6. doi: 10.5042/jcs.2010.0296 | Academic | Citation searching | Wrong publication type |
| Hunt J, Macleod A. (2008). Outcomes of applications to court for contact orders after parental separation or divorce, Family Law and Justice Division, Ministry of Justice. | Grey | Review unpicking | Wrong outcome |
| Jennings M, Howe N. (2001). Siblings' perceptions of their parents' divorce. *Journal of Divorce & Remarriage*. 35(1): 91-106. doi: https://dx.doi.org/10.1300/J087v35n01_05 | Academic | Database searches | No court involvement or insufficient information about court involvement |
| Johnson RD, Ford DV, Broadhurst K, et al. (2020). Data Resource: Population level family justice administrative data with opportunities for data linkage. *International Journal of Population Data Science*. 5(1). doi: 10.23889/ijpds.v5i1.1339 | Academic | Citation searching | Data not extractable |
| Johnston JR, Goldman JR. (2010). Outcomes of family counseling interventions with children who resist visitation: An addendum to Friedlander and Walters (2010). *Family Court Review*. 48(1): 112-5. | Academic | Review unpicking | Wrong country |
| Johnston JR, Walters MG, Olesen NW. (2005). Is it alienating parenting, role reversal or child abuse? A study of children's rejection of a parent in child custody disputes. *Journal of Emotional Abuse*. 5(4): 191-218. | Academic | Review unpicking | Wrong country |
| Jones R. (2023). Children and young people's experiences of participation in private proceedings in the family courts, Crown Copyright. https://www.gov.wales/sites/default/files/statistics-and-research/2023-12/children-and-young-peoples-experiences-of-participation-in-private-proceedings-in-the-family-courts-report.pdf | Grey | Website searches | Wrong outcome |
| Kaspiew R, Carson R, Dunstan J, et al. (2015). Experiences of Separated Parents Study (Evaluation of the 2012 Family Violence Amendments), Melbourne, Australian Institute of Family Studies. https://aifs.gov.au/sites/default/files/efva-esps_0_0.pdf | Grey | Website searches | Wrong outcome |
| Kaspiew R, Carson R, Moore S, et al. (2014). Independent Children's Lawyers Study: Final report (2nd ed.), Canberra, Attorney-General's Department. https://aifs.gov.au/sites/default/files/2022-06/IndependentChildrensLawyerStudyFinalReport.pdf | Grey | Website searches | Wrong outcome |
| Kaspiew R. (2007). Empirical insights into parental attitudes and children's interests in family court litigation. *Sydney Law Review*. 29(1): 131-61. | Academic | Citation searching | Wrong outcome |
| Kaye M, Stubbs J, Tolmie J. (2003). Negotiating Child Residence and Contact Arrangements Against a Background of Domestic Violence, Families, Law and Social Policy Research Unit, Socio-Legal Research, Centre School of Law. | Academic | Citation searching | Wrong outcome |
| Kuruppu J, Novy K, Fetter L, et al. (2023). 'Family court...sucks out your soul': Australian general practitioners' experiences supporting domestic violence survivors through family court. *BMC Primary Care*. 24(1). doi: 10.1186/s12875-023-02044-2 | Academic | Citation searching | Wrong population |
| Lehtme R. (2024). Child-centred approach to shared parenting: Children's experiences of bird's nest parenting arrangements. *Children and Youth Services Review*. 160. doi: [10.1016/j.childyouth.2024.107584](https://doi.org/10.1016/j.childyouth.2024.107584) | Academic | Database searches | Wrong country |
| Luecken LJ, Fabricius WV. (2003). Physical health vulnerability in adult children from divorced and intact families. *J Psychosom Res*. 55(3): 221-8. doi: 10.1016/s0022-3999(02)00552-4 | Academic | Review unpicking | Wrong country |
| Macdonald GS. (2017). Hearing children's voices? Including children's perspectives on their experiences of domestic violence in welfare reports prepared for the English courts in private family law proceedings. *CHILD ABUSE & NEGLECT*. 65: 1-13. doi: 10.1016/j.chiabu.2016.12.013 | Academic | Review unpicking | Wrong outcome |
| Mantle G, Leslie J, Parsons S, et al. (2006). Establishing children's wishes and feelings for family court reports: The significance attached to the age of the child. *Childhood*. 13(4): 499-518. doi: [10.1177/0907568206068560](https://doi.org/10.1177/0907568206068560) | Academic | Database searches | Wrong outcome |
| Mantle G, Moules T, Johnson K, et al. (2007). Whose wishes and feelings? Children's autonomy and parental influence in family court enquiries. *British Journal of Social Work*. 37(5): 785-805. doi: https://dx.doi.org/10.1093/bjsw/bcl035 | Academic | Database searches | Wrong outcome |
| Masson J. (2010). Judging the children act 1989: Courts and the administration of family justice. *Journal of Children's Services*. 5(2): 52-9. doi: 10.5042/jcs.2010.0301 | Academic | Citation searching | Wrong publication type |
| McIntosh J, Long C. (2005). Current Findings on Australian Children in Postseparation Disputes: Outer Conflict, Inner Discord. *Journal of Family Studies*. 11(1): 99-109. doi: doi:https://dx.doi.org/10.5172/jfs.327.11.1.99 | Academic | Database searches | No court involvement or insufficient information about court involvement |
| McIntosh J, Long C. (2006). Children Beyond Dispute: A Prospective Study of Outcomes from Child Focused and Child Inclusive Post-Separation Family Dispute Resolution, Family Transitions Pty Ltd / LaTrobe University 2006 | Grey | Website searches | Data not extractable |
| McIntosh J, Smyth B, Kelaher M, et al. (2011). Post-separation parenting arrangements: Patterns and developmental outcomes. *Studies of two risk groups. Family Matters*. 86: 40-8. | Grey | Review unpicking | No court involvement or insufficient information about court involvement |
| McIntosh JE, Bryant HD, Murray K. (2008). EVIDENCE OF A DIFFERENT NATURE: THE CHILD- RESPONSIVE AND LESS ADVERSARIAL INITIATIVES OF THE FAMILY COURT OF AUSTRALIA. *Family Court Review*. 46(1): 125-36. doi: 10.1111/j.1744-1617.2007.00187.x | Academic | Citation searching | Wrong outcome |
| McIntosh JE, Long CM, Wells YD. (2009). Children beyond dispute: A four year follow up study of outcomes from child focused and child inclusive post-separation family dispute resolution, Australian Government, Attorney-General's Department. | Academic | Citation searching | No court involvement or insufficient information about court involvement |
| McIntosh JE, Wells YD, Long CM. (2007). Child-focused and child-inclusive Family Law dispute resolution: One year findings from a prospective study of outcomes. *Journal of Family Studies*. 13(1): 8-25. doi: doi:https://dx.doi.org/10.5172/jfs.327.13.1.8 | Academic | Database searches | No court involvement or insufficient information about court involvement |
| McIntosh JE, Wells YD, Smyth BM, et al. (2008). Child-focused and child-inclusive divorce mediation: Comparative outcomes from a prospective study of postseparation adjustment. Special issue: Including children in family law proceedings--International perspectives. 46(1): 105-24. doi: doi:https://dx.doi.org/10.1111/j.1744-1617.2007.00186.x | Academic | Database searches | No court involvement or insufficient information about court involvement |
| McIntosh JE. (2009). Four young people speak about children's involvement in family court matters. *Journal of Family Studies*. 15(1): 98-103. doi: doi:https://dx.doi.org/10.5172/jfs.327.15.1.98 | Academic | Database searches | Wrong study design |
| Mitchell D, Chapman P. (2009). Pathways through parental separation: the experiences of a group of non-resident fathers, Wellington, New Zealand, Families Commission. https://thehub.sia.govt.nz/assets/documents/IP-pathways-parental-separation_FC_09.pdf | Grey | Website searches | Wrong Population |
| Moloney L, Smyth B, Richardson N, et al. (2016). Understanding parenting disputes after separation (Research Report No. 36), Melbourne, Australian Institute of Family Studies. https://apo.org.au/sites/default/files/resource-files/2016-08/apo-nid261951.pdf | Grey | Website searches | Wrong outcome |
| Moloney L, Smyth B, Weston R, et al. (2007). Allegations of Family Violence and Child Abuse in Family Law Children's Proceedings. A Pre-reform Exploratory Study. *Allegations of Family Violence and Child Abuse in Family Law Children's Proceedings: A Pre-reform Exploratory Study*. | Academic | Citation searching | Wrong outcome |
| Monk D, Macvarish J. (2018). Siblings, Contact and the Law: An Overlooked Relationship, London, Birbeck. https://www.nuffieldfoundation.org/wp-content/uploads/2019/11/Siblings20Full20Report202018.pdf | Grey | Website searches | Wrong case type |
| Morrison F. (2009). After domestic abuse: Children's Perspectives on contact with fathers. | Grey | Review unpicking | Wrong country |
| Morrison F. (2016). Children's views on contact with non-resident fathers in the context of domestic abuse. | Grey | Review unpicking | Wrong country |
| North L, Akbari A, Cowley L, et al. (2022). What do we know about ethnic diversity in the family justice system in Wales?, Nuffield Family Justice Observatory | Grey | Website searches | Wrong outcome |
| Parkinson P, Cashmore J, Single J. (2005). ADOLESCENTS' VIEWS ON THE FAIRNESS OF PARENTING AND FINANCIAL ARRANGEMENTS AFTER SEPARATION. *Family Court Review*. 43(3): 429-44. doi: 10.1111/j.1744-1617.2005.00044.x | Academic | Review unpicking | Wrong outcome |
| Parkinson P, Cashmore J, Single J. (2007). Parents' and children's views on talking to judges in parenting disputes in Australia. *International Journal of Law, Policy and the Family*. 21(1): 84-107. doi: 10.1093/lawfam/ebi022 | Academic | Review unpicking | Wrong outcome |
| Pauls M, Bertrand L. (2007). A study of perceptions and experiences of Albertans involved with child custody and access, Canadian Research Institute for Law and the Family. Alberta Law Foundation. https://prism.ucalgary.ca/bitstreams/04eb63f7-a2dd-4095-8d13-825d9845ab9b/download | Grey | Website searches | Wrong outcome |
| Peacey V, Hunt J. (2008). Problematic contact after separation and divorce. A national survey of parents. 1-146. | Grey | Review unpicking | Wrong outcome |
| Peacey V, Hunt J. (2009). I'm not saying it was easy... Contact problems in separated families. Contact problems in separated families, London, Gingerbread. | Grey | Review unpicking | No court involvement or insufficient information about court involvement |
| Private Law Working Group. (2020). Private law: family disputes the time for change the need for change the case for change. https://www.judiciary.uk/wp-content/uploads/2020/04/PRIVATE-LAW-WORKING-GROUP-REPORT.pdf | Grey | Website searches | Wrong publication type |
| Pruett MK, Williams TY, Insabella G, et al. (2003). Family and legal indicators of child adjustment to divorce among families with young children. *Journal of Family Psychology*. 17(2): 169. doi: 10.1037/0893-3200.17.2.169 | Academic | Review unpicking | Wrong country |
| Reeves C. (2008). Youth included!, SPARC: Social Planning and Research Council of British Columbia. | Grey | Review unpicking | Wrong outcome |
| Richardson S, McCabe MP. (2001). Parental divorce during adolescence and adjustment in early adulthood. *Adolescence*. 36(143): 467-89. | Academic | Review unpicking | No court involvement or insufficient information about court involvement |
| Robertson J, Pryor J, Moss J. (2008). Putting the Kids First: Caring for Children After Separation, Families Commission. https://thehub.sia.govt.nz/assets/documents/putting-kids-first_FC_07.pdf | Grey | Website searches | Wrong outcome |
| Rodgers B, Gray P, Davidscon T, et al. (2011). Parental Divorce and Adult Family, Social and Psychological Outcomes: The Contribution of Childhood Family Adversity. https://apo.org.au/node/24969 | Grey | Website searches | No court involvement or insufficient information about court involvement |
| Shea Hart A. (2004). Children exposed to domestic violence: Undifferentiated needs in Australian family law. *Australian Journal of Family Law*. 18(2): 170-92. | Academic | Citation searching | Wrong outcome |
| Smart C, May V, Wade A, et al. (2003). Residence and Contact Disputes in Court, volume 1. | Grey | Review unpicking | Wrong outcome |
| Smart C, May V, Wade A, et al. (2005). Residence and Contact Disputes in Court Volume 2. Citeseer. | Grey | Review unpicking | Wrong outcome |
| Symonds J, Dermott E, Staples E, et al. (2022). Separating families: Experiences of separation and support, Nuffield Family Justice Observatory. https://www.nuffieldfjo.org.uk/resource/separating-families-experiences-of-separation-and-support | Grey | Website searches | Wrong outcome |
| Taylor N, Gollop M, Heneghan M. (2010). Relocation Following Parental Separation: The Welfare and Best Interests of Children, Dunedin New Zealand, Centre for Research on Children and Families and Faculty of Law University of Otago. https://www.otago.ac.nz/__data/assets/pdf_file/0028/291943/read-the-research-report-630000.pdf | Grey | Website searches | No court involvement or insufficient information about court involvement |
| Taylor N, Gollop M, Liebergreen N. (2019). Parenting arrangements after Separation study: evaluating the 2014 family law reforms Family justice Professionals' perspectives, Dunedin, New Zealand, Children's Issues Centre Faculty of Law University of Otago. https://www.otago.ac.nz/__data/assets/pdf_file/0029/327692/read-the-phase-two-family-justice-professionals-perspectives-report-739547.pdf | Grey | Website searches | Wrong outcome |
| Taylor N. (2005). Care of Children: Families, Dispute Resolution and the Family Court. PhD, University of Otago. | Thesis | Website searches | Wrong publication type |
| Taylor RJ. (2001). Listening to the Children: Children of Divorce Speak Out About Their Parents. Journal of Divorce & Remarriage. 35(1-2): 147-54. doi: 10.1300/J087v35n01_09 | Academic | Review unpicking | Wrong country |
| Timms JE, Bailey S, Thoburn J. (2008). Children's views of decisions made by the court: Policy and practice issues arising from the Your Shout Too! survey. Special Issue: Improving services for children when parents are separating. 14(3): 257-74. doi: https://dx.doi.org/10.1080/13575270802061926 | Academic | Database searches | Wrong outcome |
| Trinder L, Hunt J, Macleod A, et al. (2013). Enforcing contact orders: problem-solving or punishment. | Grey | Review unpicking | Wrong outcome |
| Walsh MC, Joyce S, Maloney T, et al. (2019). Adverse childhood experiences and school readiness outcomes: Results from the Growing up in New Zealand study, Wellington, New Zealand, Ministry of Social Development. https://msd.govt.nz/documents/about-msd-and-our-work/publications-resources/research/children-and-families-research-fund/children-and-families-research-fund-report-adverse-childhood-experiences-and-school-readiness-outcomes-april-2019-final.pdf | Grey | Website searches | No court involvement or insufficient information about court involvement |
| Walsh MC, Joyce S, Maloney T, et al. (2019). Exploring the protective factors of children and families identified at highest risk of adverse childhood experiences by a predictive risk model: An analysis of the growing up in New Zealand cohort. Children and Youth Services Review. 108: 104556. doi: https://doi.org/10.1016/j.chldyouth.2019.104556 | Academic | Website searches | No court involvement or insufficient information about court involvement |
| Walsh MC, Joyce S, Maloney T, et al. (2019). Protective factors of children and families at highest risk of adverse childhood experiences: An analysis of children and families in the Growing up in New Zealand data who "beat the odds, Wellington, New Zealand, Ministry of Social Development. https://msd.govt.nz/documents/about-msd-and-our-work/publications-resources/research/children-and-families-research-fund/children-and-families-research-fund-report-protective-factors-aces-april-2019-final.pdf | Grey | Website searches | No court involvement or insufficient information about court involvement |
| Walsh MC, Macvarish J, Vaithianathan R. (2021). School Readiness, Adversities in Childhood Experience and Access to Government Services: A Scoping Study on Potential Protective Factors, Wellington, New Zealand, Ministry of Social Development https://csda.aut.ac.nz/__data/assets/pdf_file/0015/503313/ACES_FIN_1.pdf | Grey | Website searches | No court involvement or insufficient information about court involvement |
| Walsh MC, Maloney T, Vaithianathan R. (2021). Adversities of Childhood Experience and School Readiness - Focus on children born to teen and non-teen mothers in the Growing Up in New Zealand data, Wellington, New Zealand, Ministry of Social Development https://csda.aut.ac.nz/__data/assets/pdf_file/0016/503314/ACES2_Teens.pdf | Grey | Website searches | No court involvement or insufficient information about court involvement |

## ABBREVIATIONS

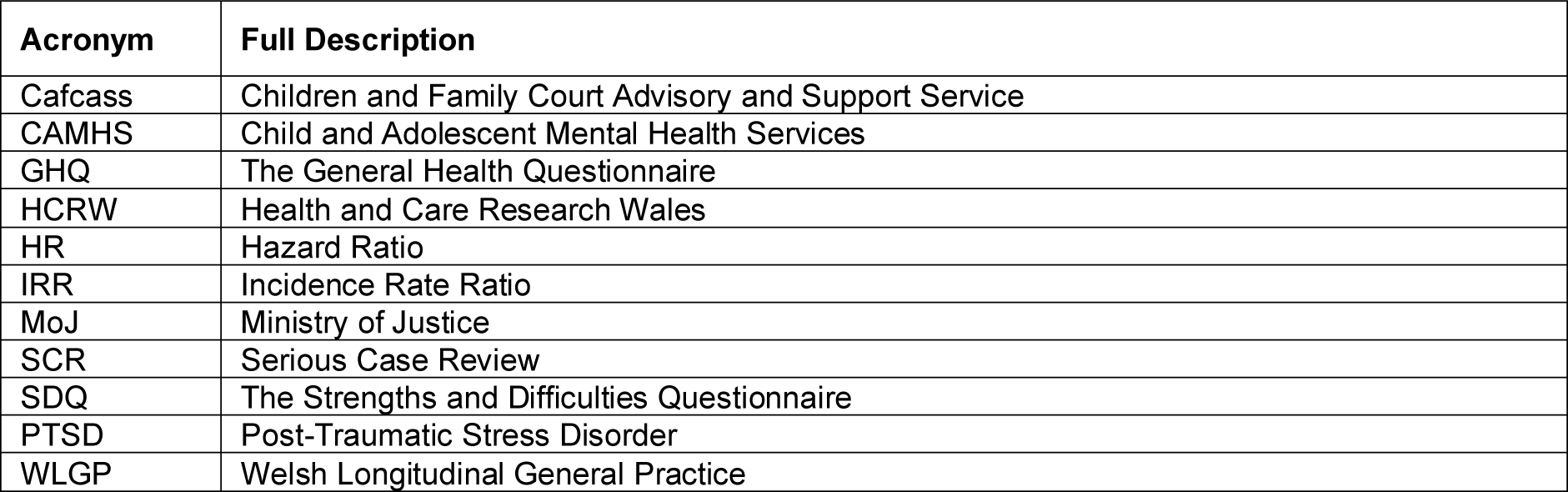

| Acronym | Full Description |
| --- | --- |
| Cafcass | Children and Family Court Advisory and Support Service |
| CAMHS | Child and Adolescent Mental Health Services |
| GHQ | The General Health Questionnaire |
| HCRW | Health and Care Research Wales |
| HR | Hazard Ratio |
| IRR | Incidence Rate Ratio |
| MoJ | Ministry of Justice |
| SCR | Serious Case Review |
| SDQ | The Strengths and Difficulties Questionnaire |
| PTSD | Post-Traumatic Stress Disorder |
| WLGP | Welsh Longitudinal General Practice |

## Footnotes

* This section has been completed by the Centre for Health Economics & Medicines Evaluation (CHEME), Bangor University

